# Infections in biological and targeted synthetic drug use in rheumatoid arthritis: where do we stand? A scoping review and meta-analysis

**DOI:** 10.1101/2023.03.09.23287022

**Authors:** BJM Bergmans, BY Gebeyehu, EP van Puijenbroek, K Van Deun, B Kleinberg, JL Murk, E de Vries

## Abstract

**Objectives:** The advent of biological and targeted synthetic therapies has revolutionised rheumatoid arthritis (RA) treatment. However, this has come at the price of an increased risk of infections. The aim of this study was to present an integrated overview of both serious and non-serious infections, and to identify potential predictors of infection risk in RA patients using biological or targeted synthetic drugs.

**Design:** We systematically reviewed available literature from PubMed and Cochrane and performed multivariate meta-analysis with meta-regression on the reported infections. Randomised controlled trials and prospective and retrospective observational studies including patient registry studies were analysed, combined as well as separately. We excluded studies focusing on viral infections only.

**Results:** Infections were not reported in a standardised manner. Meta-analysis showed significant heterogeneity that persisted after forming subgroups by study design and follow-up duration. Overall, the pooled proportions of patients experiencing an infection during a study were 0.30 (95%CI, 0.28-0.33) and 0.03 (95%CI, 0.028-0.035) for any kind of infections or serious infections only, respectively. We found no potential predictors that were consistent across all study subgroups.

**Conclusions:** The high heterogeneity and the inconsistency of potential predictors between studies show that we do not yet have a complete picture of infection risk in RA patients using biological or targeted synthetic drugs. Besides, we found non-serious infections outnumbered serious infections by a factor 10:1, but only few studies have focused on their occurrence. Future studies should apply a uniform method of infectious adverse event reporting and also focus on non-serious infections and their impact on treatment decisions and quality of life.

## Introduction

Patients with rheumatoid arthritis (RA) have an increased risk to contract serious infections which are defined as infections that are life-threatening or fatal, result in significant disability or require hospitalization[1, 2]. Aside from RA itself, which impairs an effective immune response to pathogens [3], the immunomodulating therapies used to treat RA also contribute to this risk.

Biologicals are monoclonal antibodies that target essential components in immune pathways related to RA pathogenesis. TNF-alpha inhibitors were introduced first, in the late 90’s (infliximab, etanercept, adalimumab, certolizumab pegol, golimumab). Other biologicals soon followed: anti-CD20 (B-cell) agents (rituximab), T-cell co-stimulation inhibitors (abatacept) and interleukin-(IL)-antagonists (most notably tocilizumab (anti-IL6) and anakinra (anti-IL1)). Existing categories are still expanding, and many other biological classes are investigated as potential RA treatments. Targeted synthetic drugs are the most recent addition to the RA treatment options. These are small molecules that inhibit proinflammatory cytokine production through interference with intracellular signalling pathways, for example Janus Kinase-Signal Transducer and Activator of Transcription (JAK-STAT) inhibitors (tofacitinib, filgotinib). Most RA patients need a combination of different immunomodulators to adequately control their RA.

The current knowledge on infectious adverse events following treatment of RA with biologicals originates from several study designs, each with their own strengths and weaknesses[4]. Initially, infectious adverse events were mainly reported in randomised controlled trials (RCTs), which are limited by a relatively short follow-up and the employment of exclusion criteria, such as serious comorbidities or recurrent infections. Post-marketing observational studies may have a longer follow-up and include patients that would be considered ineligible in RCTs, but generally focus on serious infections only. Case reports mostly describe (very) rare, serious infections which are likely to be missed in trials due to the shorter follow-up.

A number of factors have been identified that increase the risk of serious infections in RA, most notably the presence of comorbidities such as diabetes mellitus and cardiovascular disease [5] [6], longer RA disease duration[7], older age, extra-articular RA manifestations, a previous history of recurrent infections, higher disease activity[8–10] and corticosteroid use[3, 11]. Biological use is another risk factor. An increased rate of serious infections as compared to placebo has been reported for several biologicals[12]. Tuberculosis reactivation, for example, is a well-documented adverse effect of TNF-alpha inhibitor use[13–15]. Little attention has been paid to the specific types of infections or the occurrence of non-serious infections during biological therapy [16]. More detailed knowledge would enable clinicians to monitor patients more closely or even prevent infections.

To perform a thorough analysis of infections in biological and targeted synthetic therapy use, we included as many study designs and as many articles as possible to be able to generate a complete overview. To this end, we performed a scoping review and meta-analysis with meta-regression taking disease characteristics, patient characteristics and comorbidities into account. We determined the pooled proportion of patients that experienced serious and/or non-serious infections during treatment with biologicals or targeted synthetic therapies included in trials, observational studies and registries. We compared this information with an overview of infections published in case reports of RA patients using biological or targeted synthetic therapies. In addition, we aimed to identify risk factors for developing these infections.

## Materials and methods

We systematically collected available literature and performed a descriptive as well as meta-analysis with meta-regression following the PRISMA guidelines[17]. See S1 Appendix for the search terms and strategy. We aimed to include a wide variety of article types, as a complete overview of infections can only be obtained when analysing all available literature. Narrative reviews, meta-analyses, studies on other antirheumatic drugs, studies not reporting infectious complications, not reporting data on RA patients separately, focusing on postoperative complications, or aggregating results of different drug classes were excluded. We also excluded studies focusing on the coronavirus disease that emerged in 2019 (COVID-19) because the lockdowns created a unique situation of decreased exposure, and studies focusing on reactivating viral infections only, as they originate in a different epidemiological setting altogether. Only published, peer-reviewed articles published until January 10, 2021, written in English and collected from PubMed and Cochrane were included. This review was registered at the Open Science Framework (Registration DOI 10.17605/OSF.IO/FWTQE).

Blinded study selection was performed using Rayyan software[18]. EdV, JLM and BB performed the title and abstract selection, JLM and EdV each evaluating half and BB all hits, in order to review each article twice. Discrepancies were resolved by discussion until consensus was reached. BB screened the remaining full-text articles (flowchart in S2 Appendix). We discussed beforehand which variables would be extracted (see S3 Appendix and S4 Appendix). Two databases were constructed: one for RCTs, RCTs with open label extension studies (RCT+OLE), prospective observational studies, registry studies, open label studies and retrospective observational studies, and one for the case reports and series.

### Randomised controlled, prospective and retrospective observational studies

Studies with multiple study arms examining multiple drugs, multiple different dosages of the same drug, or studies that reported the use of more than one observational study separately were divided into multiple entries in the database. Studies using the same registry or database, and RCTs and associated extension studies were critically reviewed and in- or excluded as described in S5 Appendix and S6 Appendix to prevent duplicate data. The following information was extracted per study (arm): year of publication, country of origin, biological or targeted synthetic agent name, class and dosage, follow-up duration, number of included patients, sex, age, and other patient characteristics, laboratory parameters, being: C-Reactive Protein (CRP) or Erythrocyte Sedimentation Rate (ESR), disease activity inventories (Disease Activity Score using CRP or ESR (DAS28-CRP/ESR), Clinical Disease Activity Index for RA (CDAI), Health Assessment Questionnaire – Disability Index (HAQ-DI), Swollen Joint Count in 66 joints (SJC66), Tender Joint Count in 68 Joints (TJC68), Patient’s Global Assessment (Visual Analogue Scale 0-100), Physicians, Global Assessment (Visual Analogue Scale 0-100), Pain score (Visual Analogue Scale 0-100), Functional Assessment of Chronic Illness Therapy – Fatigue (FACIT-fatigue)) concomitant immunosuppressant use, concomitant Non-Steroidal Anti-Inflammatory Drug (NSAID) use, use of any prior conventional synthetic disease-modifying anti-rheumatic drugs (csDMARDs) or biological/targeted synthetic therapies, presence of comorbidities, number and type of infectious events during follow-up, number of serious infectious events and rate of infectious events during follow-up (details in S3 Appendix).

Infection count was extracted either as the total number of infected *patients* in a population of *n* patients, the number of infectious *events* in a population of *n* patients, an incidence rate (*n* infected patients per 100 patients) or an event rate (*n* events per 100 patient-years of exposure to study drug), as reported in the original article. Infections were extracted as either the number and/or rate of the *total* number of infections, as stated by the authors, or the number and/or rate of *serious* infections, as stated by the authors. See table 1 for terms and definitions used throughout this paper. Specific infections were grouped into composite variables by the corresponding organ-system (see S7 Appendix). Information on the occurrence of *specific* infections was also extracted, if available, in the manner stated previously.

**Table 1:** terms and definitions used throughout this paper.

| Term | Definition |
| --- | --- |
| Serious infection | A serious adverse event[2] of infectious nature, as defined by the authors |
| Total number of infected patients | Total number of infected patients in the study as stated by the authors (a patient is counted only once even though she/he could have had multiple infections) including both serious and non-serious infections |
| Total number of seriously infected patients | Total number of patients in the study that experienced an infection considered serious, as stated by the authors (a patient is counted only once even though she/he could have had multiple serious infections) |
| Total number of infectious events | Total number of infectious events in the study as |
|  | stated by the authors (a patient can be counted multiple times as she/he can experience multiple events), including both serious and non-serious infections |
| Total number of serious infectious events | Total number of serious infectious events in the study as stated by the authors (each event a patient experiences is counted, even if the patient experiences multiple serious events) |
| Incidence rate | The number of infected patients per 100 patient-years of follow-up |
| Event rate | The number of infectious events per 100 patient-years of follow-up |

### Case reports and series

Single case reports and case series from the literature search and from secondary citations were included. We analysed the case series as separate case reports (they contained 2-15 cases per article). Per case, the following information was registered in the database: country of origin, year of publication, patient characteristics (age, sex, comorbidities), time since RA diagnosis, prescribed biological/targeted therapy and duration of use, comedication, infectious diagnosis using Medical Dictionary for Regulatory Activities (MedDRA)[19] terminology, causative micro-organism, affected organ system, diagnostic method and outcome (details in S4 Appendix).

### Statistical analysis

Data analyses and visualizations were carried out using R (R Core Team, version 4.2.1, 2022-06-23) and RStudio (version 2021.9.1.372) with the packages metafor[20], dmetar[21] and ggplot2[22].

#### Randomised controlled, prospective and retrospective observational studies

We estimated the pooled proportions of the total number of infected patients and the number of seriously infected patients with their 95% confidence intervals (CI) across all studies as well as grouped by study design and biological target. The majority of the studies included in this review contributed two or more study arms and therefore multiple proportions of infections. A multivariate meta-regression model was used to account for this potential dependence between proportions belonging to the same study. Raw proportions were transformed into logit transformed proportions (i.e., the log of the proportion divided by one minus the proportion). Logit transformation was selected for its straightforward back-transformation and to ensure the confidence interval estimates fell between 0 and 1[23]. Event rates per patient-year were also estimated for total and serious infection events using multivariate meta-regression.

We used the *rma.mv* function of the *metafor package*[20] with the input of the observed effect sizes and the corresponding sampling variances; see S8 Appendix for details. The heterogeneity between the reported proportions and event rates was assessed by computing *I*^2^ [24] and Cochran’s generalized Q test [25, 26]. The *I*^2^ statistic describes the percentage of variation in the estimated effects that is explained by differences between the included studies rather than by chance. Similar to *Higgins et al.*[24], we categorised the level of heterogeneity based on *I*^2^ as low (25% to 50%), moderate (50% to 75%), and high (≥75%). The Cochran’s Q test statistic is the weighted sum of squared differences between the observed effects and the overall effects and the pooled effect across studies. The null hypothesis of the test is that all studies have the same effect size, and the alternative hypothesis is that the effect size for at least one study is different. For the Cochran’s generalized Q test, *p* ≤ 0.05 was regarded as significant heterogeneity.

We followed a two-stage procedure to deal with heterogeneity, if present. In the first stage, we attempted to create mutually exclusive sets of studies that were relatively homogeneous. We grouped studies based on study design (RCT, retrospective study, RCT+OLE, prospective cohort study, registry, open-label trial) and follow-up length (δ18 weeks; 19-38 weeks; 39 – 60 weeks) (see S8 Appendix for details) and assessed the level of heterogeneity separately for each subgroup. In the second stage, a multivariate meta-regression was used to further identify factors that significantly moderated the observed heterogeneity.

A common pitfall of meta-regression is the use of a small number of studies per examined covariate[27]. The Cochrane handbook suggests a minimum of 10 studies per covariate[28]. For our multivariate meta-regression model, we first identified potential moderators from the literature (see S9 Appendix for details). Then, all of these features that were reported by at least 10 studies were included in the model. Moderator analysis using multivariate meta-regression was performed only for RCT studies as the number of studies per subgroup was below 10 for other study designs. In comparison, the multivariate meta-regression model was also fitted for alternative grouping of studies, separately for each study design (results see S10 appendix).

We also created two composite features ourselves. The included studies used a selection of ten different disease activity indices (see above). Each study reported at least two of these indices, however, the specific indices that were used varied greatly across studies. A composite disease activity was computed by scaling each disease activity index using each index’s minimum and maximum values and then taking the mean of the scaled values (details in S11 Appendix). Out of all utilised exclusion criteria, we identified 14 unique ones that all studies used in varying combinations. To overcome this, we created a second composite variable: the summed number of exclusion criteria used in each study (details in S12 Appendix).

#### Case reports and series

In the case reports database, patient characteristics, infection location and micro-organisms were analysed using descriptive statistics.

## Results

### Trials, registries and cohort studies

The final selection for this category yielded 242 studies containing 512 study arms and a total of 293,431 patients (descriptive statistics see S13 Appendix). The largest proportion of studies were RCTs (150, 61.9%), but the highest proportion of patients were supplied by registry studies (172,892 patients, 59%). A high proportion of studies was carried out in Western countries, with 182 (79%) studies taking place in North America and/or Europe (see S14 Appendix). Out of all studies reporting on race or ethnicity, 89% of participants was Caucasian. TNF-alpha inhibitors were the most often administered biological class, with 117 studies (44%), 205 study arms (42%) and 170,826 patients (58%). Most patients were female (79.0%). The weighted mean age was 57 years. No causative micro-organisms were specified in trials, registries, or cohort studies. There was a high variability in the methods of infectious adverse event reporting. These were not mutually exclusive, with 129 studies reporting a total number of infected patients, 157 studies reporting a total number of seriously infected patients, 25 studies reporting the total number of infectious events and 38 studies reporting the total number of serious infectious events. Specific infections within the number of infected patients and number of infectious events were reported by 166 and 29 studies, respectively. Incidence and event rates were reported by a minority of studies, see S13 Appendix. Fig 1 shows the number of studies by study design and by biological target over the past three decades.

### Between-study heterogeneity and subgroups

The between-study heterogeneity was assessed first for all studies together, then for each study design separately, and finally by further grouping studies based on follow-up duration (see Fig 2). There was substantial between-study heterogeneity when all studies are taken together (both total as well as seriously infected patients). All subgroups based on study design – except the registry subgroup (which comprised only three studies) – showed significant heterogeneity. Following the pattern observed in the follow-up duration of studies (see S8 Figs 1 and 2), we defined three subgroups: ≤ 18 weeks, 19-38 weeks, and 39-60 weeks. Although the level of heterogeneity was reduced after subgroups were formed, a moderate to higher level of heterogeneity remained, especially among subgroups for the total number of infected patients. For that, we employed a multivariate meta-regression model, see S15 Appendix and S16 Appendix for the results.

**Fig 1.**
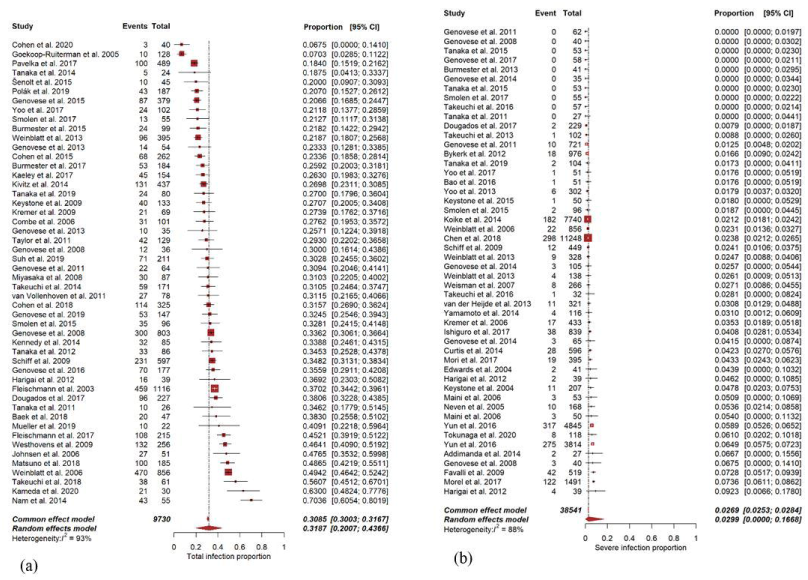
Distribution of included studies by year of publication and study design (a) and by year of publication and biological target (b) JAK= Janus kinase, OLE= open label extension, RCT= randomised controlled trial, TNF= tumour necrosis factor.

**Fig 2.**
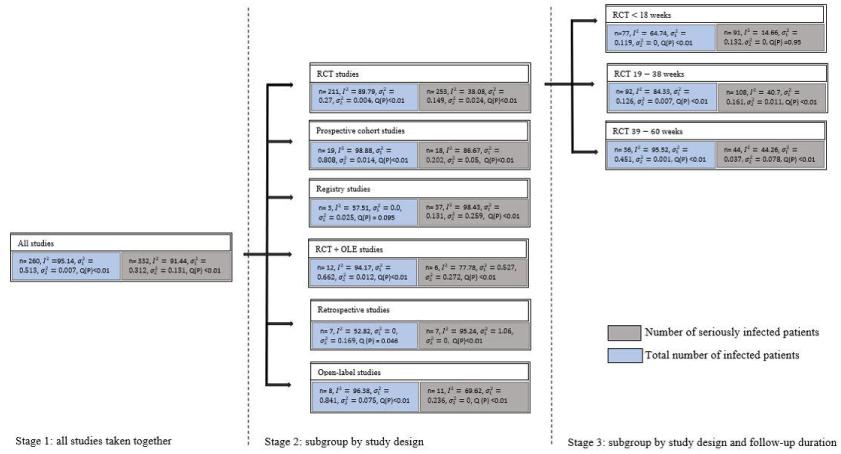
Between-study heterogeneity metrics and subgroup formation. I^2^ =variation (%) explained by differences in studies, n= number of studies/study arms, 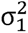 = variance (between-study), 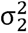 = variance (within study), Q(P)= Cochran’s Q test p-value. Overall, heterogeneity reduced sequentially as studies were grouped first based on study design (stage 2) then based on follow-up duration (stage 3). This was especially true for the number of seriously infected patients.

### Pooled proportion of infected patients and event rate estimates

The estimated pooled proportions of the total number of infected patients and of the number of seriously infected patients were determined consecutively 1) across all study arms, 2) grouped by study design, and 3) grouped by biological/synthetic therapy target. Across all study arms, this comprised 58,789 (totally infected) and 168,042 (seriously infected) participants, as there were more studies reporting on serious infections only. There was considerable between-study heterogeneity, more so in the pooled proportion of the total number of infected patients than in the pooled proportion of seriously infected patients (Fig 3). The pooled estimates and heterogeneity metrics for both the proportion of infected patients and event rates are presented in table 2. In table 2, we presented results for the categories of biological target and study design that are reported by at least two studies (n≥2) as it is the minimum number (n) required to compute the pooled estimates using multivariate meta-regression.

**Fig 3.**
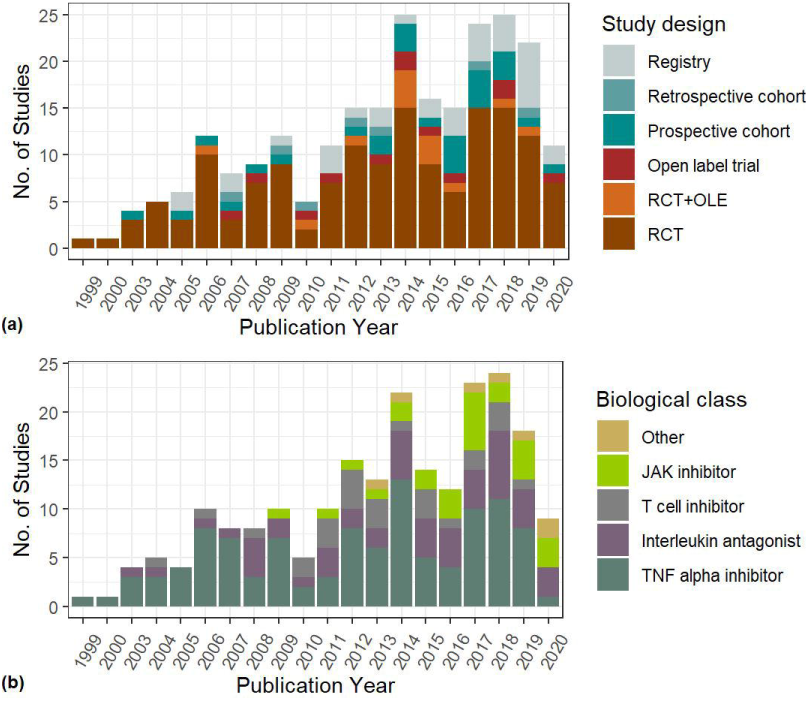
Forest plot for 50 randomly selected RCT studies, (a) for the total number of infected patients, (b) for seriously infected patients. 95% CI = 95% confidence interval. Events = number of (seriously) infected patients. Total = total number of patients included in the study. A forest plot visually illustrates the relationship between included studies and gives an overview of heterogeneity of individual results. For a complete overview of forest plots, see S17 Appendix.

**Table 2.** a. Pooled proportion estimates 2 b. Pooled event rate estimates.

| Table 2a. proportions | Number of infected patients |  |  |  |  | Number of seriously infected patients |  |  |  |  |
| --- | --- | --- | --- | --- | --- | --- | --- | --- | --- | --- |
| | Pooled estimate | n | 95% CI | $I^2$ | Cochran's Q test(P) | Pooled estimate | n | 95% CI | $I^2$ | Cochran's Q test(P) |
| All studies | 0.30 | 260 | [0.28, 0.33] | 95.14 | < 0.001 | 0.03 | 332 | [0.03, 0.03] | 91.44 | < 0.001 |
| Study design |  |  |  |  |  |  |  |  |  |  |
| RCT | 0.35 | 211 | [0.32, 0.37] | 89.79 | < 0.001 | 0.03 | 253 | [0.02, 0.03] | 38.08 | 0.001 |
| Prospective cohort | 0.16 | 19 | [0.10, 0.23] | 98.89 | < 0.001 | 0.05 | 18 | [0.03, 0.06] | 86.67 | < 0.001 |
| Registry | 0.23 | 3 | [0.19, 0.27] | 57.51 | 0.1 | 0.07 | 37 | [0.05, 0.09] | 98.43 | < 0.001 |
| RCT+OLE | 0.36 | 12 | [0.21, 0.53] | 94.17 | < 0.001 | 0.04 | 6 | [0.01, 0.10] | 77.78 | < 0.001 |
| Retrospective c. | 0.10 | 7 | [0.07, 0.15] | 52.82 | 0.04 | 0.06 | 7 | [0.02, 0.19] | 95.24 | < 0.001 |
| Open label trial | 0.18 | 8 | [0.10, 0.32] | 96.38 | < 0.001 | 0.03 | 11 | [0.02, 0.05] | 69.62 | < 0.001 |
| Biological target |  |  |  |  |  |  |  |  |  |  |
| TNF alpha inhibitor | 0.30 | 95 | [0.26, 0.35] | 96.12 | < 0.001 | 0.03 | 113 | [0.03, 0.04] | 94.63 | < 0.001 |
| B- cell inhibitor | 0.37 | 24 | [0.30, 0.45] | 86.5 | < 0.001 | 0.03 | 18 | [0.02, 0.05] | 61.27 | 0.01 |
| Rituximab | 0.29 | 11 | [0.20, 0.40] | 96.16 | < 0.001 | 0.06 | 21 | [0.04, 0.09] | 96.77 | < 0.001 |
| T-cell inhibitor | 0.34 | 12 | [0.21, 0.50] | 99.17 | < 0.001 | 0.03 | 20 | [0.02, 0.05] | 95.33 | < 0.001 |
| JAK1/3 inhibitor | 0.25 | 10 | [0.20, 0.30] | 36.30 | 0.06 | 0.02 | 20 | [0.01, 0.03] | 31.6 | 0.48 |
| JAK1/2 inhibitor | 0.40 | 12 | [0.33, 0.47] | 81.61 | < 0.001 | 0.02 | 15 | [0.02, 0.03] | 0 | 0.96 |
| JAK1 inhibitor | 0.32 | 15 | [0.27, 0.37] | 65.75 | 0.03 | 0.02 | 15 | [0.012, 0.03] | 0 | 0.76 |
| Pan-JAK inhibitor | 0.25 | 4 | [0.17, 0.34] | 53.25 | 0.09 | 0.02 | 14 | [0.01, 0.03] | 25.85 | 0.88 |
| IL6 inhibitor | 0.30 | 48 | [0.24, 0.36] | 95.33 | < 0.001 | 0.04 | 59 | [0.03, 0.04] | 77.29 | < 0.001 |
| IL17 inhibitor | 0.26 | 17 | [0.21, 0.31] | 36.12 | 0.29 | 0.02 | 11 | [0.01, 0.05] | 9.70 | 0.94 |
| IL1 inhibitor | 0.23 | 2 | [0.04, 0.67] | 85.40 | 0.01 | 0.03 | 7 | [0.02, 0.06] | 74.33 | 0.001 |
| IL23 inhibitor | 0.23 | 2 | [0.16, 0.32] | 0 | 0.78 | 0.03 | 2 | [0.01, 0.09] | 0 | 0.36 |
| IL 12/IL 23 inhibitor | 0.31 | 2 | [0.18, 0.47] | 62.76 | 0.10 | 0.01 | 2 | [0, 0.07] | 0 | 0.68 |
| BTK inhibitor | 0.11 | 4 | [0.08, 0.16] | 3.69 | 0.39 | 0.01 | 4 | [0.01, 0.04] | 0 | 0.94 |
| SYK inhibitor | - | - | - | - | - | 0.02 | 2 | [0.01, 0.06] | 0 | 0.63 |
| GM-CSF inhibitor | - | - | - | - | - | 0.01 | 7 | [0.01, 0.04] | 0 | 1 |
| Table 2b. event rates | Total number of infected patients |  |  |  |  | Number of seriously infected patients |  |  |  |  |
| | ER | n | 95% CI | $I^2$ | Cochran's Q test(P) | ER | n | 95% CI | $I^2$ | Cochran's Q test(P) |
| **All studies** | 0.92 | 8 | [0.34, 1.50] | 99.82 | <0.001 | 0.05 | 44 | [0.03, 0.08] | 99.44 | <0.001 |
| **Study design** |  |  |  |  |  |  |  |  |  |  |
| **RCT** | - | - | - | - | - | 0.03 | 7 | [0.01, 0.086] | 87.97 | <0.001 |
| **Prospective cohort** | - | - | - | - | - | - | - | - | - | - |
| **Registry** | 0.76 | 3 | [0.67, 0.83] | 79.03 | 0.01 | 0.07 | 32 | [0.03, 0.11] | 99.71 | <0.001 |
| **RCT+OLE** | 1.32 | 3 | [0.03, 2.60] | 99.75 | <0.001 | 0.04 | 4 | [0.03, 0.05] | 43.15 | 0.38 |
| **Retrospective cohort** | - | - | - | - | - | - | - | - | - | - |
| **Open label trial** | - | - | - | - | - | - | - | - | - | - |
| **Biological target** |  |  |  |  |  |  |  |  |  |  |
| **TNF alpha inhibitor** | - | - | - | - | - | 0.05 | 19 | [0.00, 0.10] | 99.80 | <0.001 |
| **IL 6 inhibitor** | 0.53 | 5 | [0.30, 0.77] | 99.24 | <0.001 | 0.04 | 9 | [0.03, 0.06] | 94.63 | <0.001 |
| **Rituximab** | - | - | - | - | - | 0.09 | 4 | [0.01, 0.18] | 99 | <0.001 |
| **T cell inhibitor** | - | - | - | - | - | 0.07 | 6 | [0.0, 0.17] | 99.71 | <0.001 |
| **B cell inhibitor** | - | - | - | - | - | 0.04 | 5 | [0.02, 0.07] | 73.98 | 0.01 |

*BTK = Bruton tyrosine kinase, CI = 95% confidence interval, ER = pooled event rate, GM-CSF = granulocyte-macrophage colony stimulating factor, IL = interleukin, JAK*
*= Janus kinase, n = number of study arms, RCT = randomised controlled trial, RCT+OLE = randomised controlled trial and open label extension, SYK = spleen tyrosine*
*kinase, TNF = tumour necrosis factor*

The estimated pooled proportion of the total number of infected patients was highest in RCT ± OLE and RCT studies. However, the differences between the estimated pooled proportions must be interpreted with caution as the heterogeneity as well as the number of studies per design subgroup varied substantially. The estimated pooled proportion of seriously infected patients was highest in registry and retrospective studies; however, the heterogeneity was also highest in these two study designs.

Fewer studies reported the infection event rates; 37 study arms (25 studies) and 79 study arms (38 studies) reported 7,821 infectious events in 13,937 patients and 6,100 serious infectious events in 90,204 patients, respectively.

### Multivariate meta-regression

A multivariate meta-regression model was fitted using one moderator at a time to explore the association between demographic, biological/targeted therapy and methodological characteristics as potential predictors of the logit of infection prevalence in RCT studies with a follow-up duration of ≤ 18 weeks, 19-38 weeks, and 39 - 60 weeks. For the purpose of readability, we included the results of the meta-regression analyses in S15 Appendix (follow-up duration of ≤ 8 weeks and 19-38 weeks) and S16 Appendix (follow-up duration 39-60 weeks). A separate analysis by follow-up duration was carried out with anticipation that different sets of moderators are responsible for the heterogeneity in the three subgroups. No single moderator was found to be significant across all three groups. In both the total number of infected patients as well as in the number of seriously infected patients, the subgroup of 39-60 weeks of follow-up was the smallest and most heterogenous, with results not significantly different from other subgroups. More information on how studies are grouped based on study design and follow-up duration is available in S8 Appendix.

### Type of infections

Across all biological/targeted synthetic therapy classes, upper respiratory tract (URT) infections were the most prevalent (12.7%, 95%CI 10.6-15.0), followed by genitourinary tract infections (3.5%, 95%CI 2.9-4.2) and lower respiratory tract (LRT) infections (2.2%, 95%CI 1.9-2.7). See table 3. Individual infection proportions were comparable across biological classes. TNF-alpha inhibitors were associated with an increased proportion of mycobacterial infections (0.9%, 95%CI 0.6-1.0) compared to other drug classes. Overall, the proportion of seriously infected patients was highest in rituximab users (6.7%, 95%CI 4.5-9.7). The proportion of LRT infections (3.3%, 95%CI 1.6-6.5), and skin and soft tissue infections (2.3%, 95%CI 0.4-13.4) was also relatively high in this group, as was the proportion of Herpes Zoster (11.8%, 95%CI 3.1-35.4), however, only few studies reported on these adverse events. Compared to other drug classes, tofacitinib use was associated with an increased proportion of Herpes Zoster (2.8%, 95%CI 1.3-6.0) and pneumocystis jirovecii pneumonia (PJP) (1.5%, 95%CI 0.4-4.5).

**Table 3:** estimated pooled proportions of the various infections across different biological classes and targeted synthetic therapies.

|  | Total |  |  |  |  |  | TNF-alpha inhibitors |  |  |  |  |  | IL-6 inhibitors |  |  |  |  |  |
| --- | --- | --- | --- | --- | --- | --- | --- | --- | --- | --- | --- | --- | --- | --- | --- | --- | --- | --- |
|  | Prop | 95%CI | I2 (%) | k | events | observations | Prop | 95%CI | I2 (%) | k | events | observations | Prop | 95%CI | I2 (%) | k | events | observations |
| **Total infected patients** | 0.3029 | [0.2765-0.3306] | 95.14 | 260 | 15515 | 58789 | 0.3031 | [0.2604-0.3494] | 96.12 | 95 | 5725 | 24088 | 0.2971 | [0.2402-0.3610] | 95.34 | 48 | 3111 | 11225 |
| **Seriously infected patients** | 0.0308 | [0.0275-0.0345] | 91.45 | 332 | 8969 | 168042 | 0.0323 | [0.0271-0.0385] | 94.63 | 113 | 4829 | 83393 | 0.0359 | [0.0295-0.0437] | 77.29 | 59 | 1137 | 24568 |
| **URT** | 0.1270 | [0.1067-0.1504] | 95.79 | 252 | 6887 | 57167 | 0.1332 | [0.0997-0.1757] | 97.31 | 82 | 3193 | 170826 | 0.1515 | [0.0958-0.2314] | 95.68 | 42 | 1067 | 45379 |
| **LRT** | 0.0229 | [0.0192-0.0274] | 86.12 | 159 | 1607 | 293431 | 0.0233 | [0.0181-0.0301] | 89.17 | 67 | 959 | 43271 | 0.0158 | [0.0107-0.0233] | 89.17 | 22 | 246 | 12429 |
| **Genitourinary** | 0.0354 | [0.0294-0.0425] | 75.66 | 121 | 859 | 25599 | 0.0356 | [0.0255-0.0495] | 83.69 | 41 | 341 | 10020 | 0.0388 | [0.0278-0.0540] | 50.72 | 19 | 104 | 3472 |
| **Gastrointestinal** | 0.0127 | [0.0094-0.0172] | 71.08 | 67 | 228 | 19437 | 0.0098 | [0.0064-0.0151] | 60.49 | 23 | 77 | 8784 | 0.0154 | [0.0085-0.0278] | 61.72 | 13 | 39 | 3388 |
| **Skin-soft tissue** | 0.0122 | [0.0092-0.0161] | 78.83 | 73 | 377 | 28054 | 0.0117 | [0.0070-0.0196] | 82.96 | 27 | 150 | 8943 | 0.0150 | [0.0093-0.0241] | 82.24 | 18 | 168 | 11514 |
| **Bone and joint** | 0.0070 | [0.0050-0.0100] | 40.9 | 73 | 81 | 15212 | 0.0086 | [0.0065-0.0114] | 0 | 16 | 49 | 6205 | 0.0059 | [0.0028-0.0125] | 49.27 | 8 | 23 | 7192 |
| **Sepsis** | 0.0057 | [0.0040-0.0082] | 68.2 | 36 | 153 | 39927 | 0.0055 | [0.0033-0.0094] | 75.88 | 20 | 104 | 28321 | 0.0079 | [0.0034-0.0179] | 59.51 | 5 | 32 | 6494 |
| **Mycobacterial** | 0.0072 | [0.0050-0.0105] | 79.98 | 47 | 254 | 40844 | 0.0095 | [0.0063-0.0143] | 77.42 | 35 | 206 | 25426 | 0.0055 | [0.0039-0.0076] | 0 | 2 | 35 | 6412 |
| **Neurological infections** | 0.0147 | [0.0117-0.0184] | 65.15 | 78 | 371 | 34681 | 0.0106 | [0.0082-0.0136] | 20.25 | 25 | 121 | 14436 | 0.0109 | [0.0067-0.0177] | 61.52 | 11 | 64 | 8941 |
| **Fungal** | 0.0088 | [0.0043-0.0182] | 77.66 | 17 | 39 | 12760 | 0.0078 | [0.0025-0.0238] | 83.48 | 9 | 23 | 10081 | NA | NA | NA | 0 | 2 | 32 |
| **Viral** | 0.0180 | [0.0138-0.0235] | 7.7 | 24 | 13 | 4693 | 0.0188 | [0.0140-0.0252] | 0 | 11 | 12 | 2554 | 0.0124 | [0.0040-0.0377] | 0 | 3 |  |  |
| **Other** | 0.0119 | [0.0072-0.0198] | 74.69 | 31 | 101 | 11869 | 0.0081 | [0.0053-0.0124] | 0% | 8 | 21 | 2901 | 0.0060 | [0.0030-0.0119] | 0 | 4 | 8 | 1401 |
| **Herpes Zoster** | 0.0139 | [0.0111-0.0174] | 60.19 | 97 | 366 | 36058 | 0.0101 | [0.0078-0.0131] | 18.48 | 29 | 116 | 14776 | 0.0106 | [0.0066-0.0172] | 59.22 | 12 | 64 | 9122 |
| **Salmonellosis** | 0.0024 | [0.0009-0.0065] | 0 | 3 | 4 | 1664 | 0.0025 | [0.0006-0.0100] | 0 | 2 | 2 | 814 | NA | NA | NA | 0 | 2 | 850 |
| **PJP** | 0.0048 | [0.0029-0.0079] | 63.65 | 31 | 91 | 35134 | 0.0038 | [0.0024-0.0062] | 50.99 | 16 | 78 | 28678 | 0.0032 | [0.0002-0.0587] | 67.47 | 2 | 1 | 920 |
| **Tuberculosis** | 0.0042 | [0.0031-0.0056] | 49.51 | 95 | 171 | 53277 | 0.0061 | [0.0044-0.0084] | 49.48 | 53 | 157 | 29592 | 0.0012 | [0.0006-0.0024] | 0 | 8 | 5 | 8046 |

|  | Rituximab |  |  |  |  |  | Abatacept |  |  |  |  |  | Tofacitinib |  |  |  |  |  |
| --- | --- | --- | --- | --- | --- | --- | --- | --- | --- | --- | --- | --- | --- | --- | --- | --- | --- | --- |
|  | prop | 95%CI | I2 (%) | k | event<br>s | observatio<br>ns | Prop | 95%CI | I2<br>(%) | k | event<br>s | observatio<br>ns | Prop | 95%CI | I2<br>(%) | k | event<br>s | observatio<br>ns |
| Total infected patients | 0.2875 | [0.1988-0.3962] | 96.72 | 8 | 772 | 4108 | 0.3321 | [0.2723-0.3978] | 99.17 | 12 | 2117 | 8423 | 0.2459 | [0.2022-0.2956] | 36.25 | 10 | 141 | 586 |
| Seriously infected patients | 0.0665 | [0.0450-0.0973] | 96.59 | 19 | 1323 | 14285 | 0.0335 | [0.0238-0.0469] | 95.34 | 20 | 1382 | 30169 | 0.0208 | [0.0144-0.0299] | 31.62 | 20 | 68 | 3576 |
| URT | 0.1601 | [0.1123-0.2232] | 75.69 | 8 | 180 | 1022 | 0.0887 | [0.0314-0.2264] | 99.34 | 13 | 908 | 40098 | 0.0840 | [0.0342-0.1919] | 96.71 | 21 | 450 | 3296 |
| LRT | 0.0327 | [0.0162-0.0652] | 80.21 | 11 | 51 | 2398 | 0.0165 | [0.0076-0.0352] | 92.5 | 13 | 141 | 8606 | 0.0244 | [0.0140-0.0421] | 59.78 | 12 | 48 | 2290 |
| Genitourinary | 0.0452 | [0.0350-0.0581] | 0 | 6 | 57 | 1306 | 0.0140 | [0.0053-0.0361] | 55.7 | 7 | 72 | 3659 | 0.0325 | [0.0185-0.0566] | 65.48 | 6 | 45 | 1550 |
| Gastrointestinal | 0.0080 | [0.0014-0.0454] | 38.57 | 2 | 2 | 359 | 0.0037 | [0.0016-0.0083] | 34.56 | 6 | 11 | 3390 | 0.0192 | [0.0061-0.0585] | 58.95 | 7 | 11 | 999 |
| Skin-soft tissue | 0.0231 | [0.0036-0.1349] | 82.01 | 4 | 7 | 569 | 0.0058 | [0.0034-0.0099] | 8 | 6 | 13 | 2484 | 0.0097 | [0.0036-0.0264] | 38.4 | 3 | 7 | 778 |
| Bone and joint | NA | NA | NA | <1 | NA | NA | 0.0031 | [0.0010-0.0095] | 0 | 2 | 3 | 996 | NA | NA | NA | <1 | 1 | 201 |
| Sepsis | NA | NA | NA | <1 | 6 | 2484 | 0.0034 | [0.0013-0.0090] | 0 | 2 | 4 | 1372 | 0.0057 | [0.0018-0.0176] | 0 | 3 | 3 | 647 |
| Mycobacterial | NA | NA | NA | <1 | 1 | 2484 | 0.0037 | [0.0005-0.0261] | 82.23 | 4 | 7 | 4595 | 0.0060 | [0.0016-0.0218] | 20.98 | 2 | 3 | 598 |
| Neurological infections | NA | NA | NA | <1 | 2 | 12 | 0.0297 | [0.0035-0.2087] | 81.76 | 6 | 43 | 4200 | 0.0293 | [0.0135-0.0624] | 80.58 | 11 | 67 | 2174 |
| Fungal | 0.0035 | [0.0015-0.0085] | 0 | 2 | 5 | 1469 | 0.0047 | [0.0018-0.0125] | 0 | 2 | 4 | 881 | NA | NA | NA | <1 | NA | NA |
| Viral | 0.0109 | [0.0015-0.0761] | 92.03 | 2 | NA | NA | NA | NA | NA | <1 | NA | NA | NA | NA | NA | <1 | NA | NA |
| Other | 0.0038 | [0.0010-0.0145] | 46.01 | 4 | 4 | 1966 | 0.0087 | [0.0063-0.0119] | 0 | 3 | 38 | 4477 | 0.0257 | [0.0172-0.0384] | 0 | 8 | 23 | 928 |
| Herpes Zoster | 0.1176 | [0.0314-0.3538] | 5.97 | 2 | 2 | 25 | 0.0087 | [0.0063-0.0119] | 0 | 3 | 43 | 4356 | 0.0282 | [0.0131-0.0597] | 77.67 | 13 | 67 | 2280 |
| Salmonellosis | NA | NA | NA | <1 | NA | NA | 0.0101 | [0.0075-0.0136] | 0 | 3 | 0 | 0 | NA | NA | NA | <1 | NA | NA |
| PJP | NA | NA | NA | <1 | NA | NA | NA | NA | NA | <1 | 6 | 4719 | 0.0145 | [0.0042-0.0488] | 0 | 5 | 2 | 209 |
| Tuberculosis | NA | NA | NA | <1 | 1 | 2484 | 0.0019 | [0.0007-0.0054] | 29.26 | 4 | 3 | 7973 | 0.0032 | [0.0009-0.0110] | 27.13 |  | 3 | 1869 |

Events= number of infected patients, K= selected number of included studies, LRT= lower respiratory tract, NA = not available, Observations = total number of patients in included studies, PJP= Pneumocystis jirovecii pneumonia, Prop= pooled proportion of (seriously) infected patients, URT= upper respiratory tract, 95%CI = 95% confidence interval. Herpes Zoster infections are included in neurological infections, salmonellosis in of gastrointestinal infections, PJP in lower respiratory tract infections and tuberculosis in mycobacterial infections.

### Case reports and series

The final selection yielded 372 articles in this category, comprising 503 cases and 509 identified micro-organisms. Most articles originated in the US (83, 22.3%) and Japan (77, 20.7%). Described patients were mostly female (66.8%), with a mean age of 61 years. Most described patients (398, 82%) used TNF-alpha inhibitors (details in S18 Appendix). The median onset of an infectious adverse event was 9 months after start of biological/targeted therapy (interquartile range (IQR) 3-24). Of all reported pathogens, 326 (64%) were bacterial (of which 143 mycobacterial infections), 19.4% (99) fungal, 7.9% (40) parasitic and 8.6% (44) viral (see S19 Appendix and S20 Appendix). Overall, the most frequently reported infections (187, 32%) occurred in the lower respiratory tract (LRT). The most frequently reported pathogens were *Mycobacterium tuberculosis* (85, 16.7% of pathogens), nontuberculous mycobacteria (50, 9.8%), *Pneumocystis jirovecii* (28, 5.5%), *Salmonella spp* (28, 5.5%), *Listeria monocytogenes* (24, 4.7%), *Histoplasma capsulatum* (22, 4.3%%) and *Leishmania spp* (21, 4.1%). The largest proportion of these infections were reported in TNF-alpha inhibitor users, who are known to have difficulty clearing mycobacterial and intracellular bacterial infections.

A relatively high number of viral (38%) and fungal (31%) infections were reported in abatacept users, however, absolute numbers were low. Relatively few fungal infections were reported in interleukin antagonist users (2, 3.6%), conversely, a relatively high number of other than intracellular bacterial infections was reported. This was also true for rituximab. Only three infections were reported in JAK-inhibitors, all fungal; see S20 Appendix.

## Discussion

We performed a scoping review and meta-analysis with meta-regression to create an overview of and identify risk factors for infectious complications of biological and targeted synthetic therapies in RA patients. We report upon the combined results of 242 studies (trials/registries) with in total 293,431 included patients. To the best of our knowledge, this is the largest meta-analysis on infections in biological and targeted synthetic drug use in RA patients to date.

We found a very high between-study heterogeneity across all analyses. We presume this was not only caused by differences in methodology, but also by differences in infectious adverse event reporting due to a lack of standardised definitions for non-serious infections and therefore variable interpretations. Furthermore, the registration of infectious adverse events during a clinical trial is in itself subject to some degree of bias and inconsistency [29] [30] [31]. Standardised definitions would reduce heterogeneity, as illustrated by a lower value in the pooled proportion of seriously as opposed to total infected patients (see Fig 2): only a definition of serious infection is currently available. We tried to mitigate this effect by grouping individual infections by the affected organ system. These shortcomings are important to consider when interpreting the results of this review, and of future studies as well.

Our data show non-serious infections to be the most prevalent infectious adverse events during biological or targeted synthetic drug use in RA, with 30% of patients being affected at some point in their treatment (vs. 2-3% being affected by a serious infection). Exotic and opportunistic infections, often the subject of the 372 identified case reports/series (containing 503 individual cases), were (very) rare in trials and cohort studies, indicating a poor reflection of the real-world prevalence/incidence. The upper respiratory tract, skin and soft tissues and urinary tract were most frequently affected in non-serious infections in the studies [16, 32]. Infections frequently led to treatment discontinuation, for which their recurrent nature may be more important than their severity [33]. Furthermore, recurrent infections are associated with a high patient-experienced burden and socioeconomic costs[34–38]. However, only few studies have made the occurrence of non-serious infections and their potential implications for treatment and quality of life in patients a priority[16, 32]. In all studies, TNF-alpha inhibitors were the most frequently administered drugs. We found only a few differences in pooled proportions of individual infections across various biological classes. JAK-inhibitor users had an increased proportion of Herpes Zoster and PJP, which is consistent with existing literature[39]. The proportion of mycobacterial infections was higher in TNF-alpha inhibitor users at 0.6% (95%CI 0.4-0.8) (see Table 3), however, these results were pooled from studies that in many cases excluded patients with a history of TB. Furthermore, 79% of studies took place in North America and/or Europe, where TB is not prevalent, and 89% of participants were Caucasian. As for the case reports, the majority originated in the US (83, 22%), Japan (21%) or France (39, 10%). This geographical spread of studies is most likely a reflection of the biological/targeted drug consumption, which is correlated with accessibility, that is not equally distributed around the world[40]. Therefore, little is known about the risks for infectious complications were these drugs to be used more in populations where other, ‘exotic’, infections are prevalent. As for predictive risk factors, we found no moderator that was significant across all three RCT subgroups for the total number of infected patients. However, several moderators were significant in one subgroup: corticosteroids below 18 weeks of follow-up, and RA duration, mean number of prior csDMARDs, leflunomide use, biological target and the use of one prior biological between 18-38 weeks of follow-up. In the above-38-week-follow-up group, no moderator was identified as significant; this was the most heterogenous group. Corticosteroids, leflunomide use, a longer RA duration and a high number of prior csDMARDs were previously associated with an increased risk of serious infection, and prior biological use seemed to decrease this risk [10, 41–45]. Previous studies also show differences in risks for serious infections between different biologicals [12]. Interestingly, though prior literature shows a dose-dependent effect of corticosteroids on the occurrence of serious infections, we could not confirm this. This may be explained by the exclusion of patients using high-dose corticosteroids from most RCTs and by the relatively short duration of corticosteroid therapy. When looking at seriously infected patients only (table 3), mean ESR at baseline seemed to significantly correlate with an increased proportion of seriously infected patients. This also has been found in one previous study[43].

As most included studies were RCTs (of which a significant number excluded patients with comorbidities or recurrent or chronic infections, see S12 Appendix), it is unclear to what extent our findings can be extrapolated to real-world settings in which patients have multiple comorbidities and use many different immunomodulators. This is a limitation of our study. The broad scope is without doubt this study’s greatest strength. This enabled us to analyse information on all aspects of infectious complications of biological/synthetic targeted treatment in RA, using different study designs that complement each other’s limitations. This review includes not only pooled proportions of the total number of (seriously) infected patients, but also proportions of patients contracting specific infections, and an analysis on potential predictors of infectious adverse events. The addition of case reports provides information on various rare serious infections and their causative pathogens, data that are generally not available from RCT studies.

In conclusion, *non*-serious infections outnumbered serious infections by 10:1 in RA patients using biological/targeted synthetic therapies, however, only little attention has been paid to them in existing literature. None of the risk factors for developing infections that were previously identified in separate studies were consistently confirmed in our meta-analysis. Further prospective research is needed using uniform infectious adverse event registration.

## Supporting information

PRISMA checklist

## Data Availability

All data produced in the present work are contained in the manuscript and the supplementary material

## Author contributions

BB – conception and design, data acquisition, article selection, data analysis, writing of manuscript

BG – data analysis, writing of manuscript

EP – critical revision of manuscript

KD - critical revision of manuscript

BK - critical revision of manuscript

JM – conception and design, article selection, critical revision of manuscript

EV – conception and design, article selection, critical revision of manuscript

## Ethics approval

This article is based on previously conducted studies and does not contain any new studies with human participants or animals performed by any of the authors.

## Data availability

All data generated or analysed during this study are included in this published article/as supplementary information files. All included articles are publicly available.

## Prior presentation

An abstract of this research was presented in poster form at the European Society for Immunodeificiencies (ESID), 12-15 October 2022, Gothenburg, Sweden.

## Supporting information

**S1 Appendix. Search strategy and terms**

**S2 Appendix. Flowchart of study in- and exclusion**

**S3 Appendix. Empty database format for RCTs, prospective observational studies, registries, open label studies, open label extension studies and retrospective observational studies**

**S4 Appendix. Empty database format for case reports**

**S5 Appendix. Studies making use of the same databases: inclusion, exclusion and reasoning**

**S6 Appendix. RCTs and extension studies: inclusion, exclusion and reasoning**

**S7 Appendix. Composition of infection composite variables**

**S8 Appendix. Subgroup construction**

**S9 Appendix. Variable selection**

**S10 Appendix. Multivariate meta-regression of the total number of infected patients by study design**

**S11 Appendix. Creation of RA severity composite variable**

**S12 Appendix. Creation of exclusion criteria composite variable**

**S13 Appendix. Characteristics of included trials**

**S14 Appendix. Geographical location of trials and case reports**

**S15 Appendix. Multivariate meta-regression for the total number of (seriously) infected patients in RCT studies with a follow-up duration of 18 weeks or less and between 19-38 weeks**

**S16 Appendix. Multivariate meta-regression for the total number of (seriously) infected patients in RCTs with a follow-up duration of 38-60 weeks**

**S17 Appendix. Additional forest plots**

**S18 Appendix. Characteristics of case reports**

**S19 Appendix. List of identified micro-organisms in case reports S20 Appendix. Figures related to the analysis of the case reports**

