## Supplementary appendix for "Infections in biological and targeted synthetic drug use in rheumatoid arthritis: where do we stand? A scoping review and meta-analysis"

**TABLE OF CONTENTS**

| **Appendix** | **Content** | **page** |
| --- | --- | --- |
| Appendix 1 | Search strategy and terms | 3 |
| Appendix 2 | Flowchart of study in- and exclusion | 8 |
| Appendix 3 | Empty database format for RCTs, prospective observational studies, registries, open label studies, open label extension studies and retrospective observational studies | 9 |
| Appendix 4 | Empty database format for case reports | 10 |
| Appendix 5 | Studies making use of the same databases: inclusion, exclusion and reasoning | 11 |
| Appendix 6 | RCTs and extension studies: inclusion, exclusion and reasoning | 13 |
| Appendix 7 | Composition of infection composite variables | 16 |
| Appendix 8 | Subgroup construction | 18 |
| Appendix 9 | Variable selection | 23 |
| Appendix 10 | Multivariate meta-regression of the total number of infected patients by study design | 32 |
| Appendix 11 | Creation of RA severity composite variable | 39 |
| Appendix 12 | Creation of exclusion criteria composite variable | 40 |
| Appendix 13 | Characteristics of included trials | 41 |
| Appendix 14 | Geographical location of trials and case reports | 44 |
| Appendix 15 | Multivariate meta-regression for the total number of (seriously) infected patients in RCT studies with a follow-up duration of 18 weeks or less and between 19-38 weeks | 45 |
| Appendix 16 | Multivariate meta-regression for the total number of (seriously) infected patients in RCTs with a follow-up duration of 38-60 weeks | 48 |
| Appendix 17 | Additional forest plots | 50 |
| Appendix 18 | Characteristics of case reports | 54 |
| Appendix 19 | List of identified micro-organisms in case reports | 60 |
| Appendix 20 | Figures related to the analysis of the case reports | 64 |

**Appendix 1: search strategy and terms**

We performed the literature search in two episodes. At first, a search was performed in December 2019 in PubMed with search terms for biological drugs. Immediately thereafter, the COVID-19 pandemic hit and the study was put on hold. In January 2021, an additional search was performed in both PubMed and Cochrane and aside from biological drugs, targeted synthetic drugs were included as well.

Pubmed:

((TNF-alpha inhibitor* [tiab] OR TNF-alpha antagonist* [tiab] OR tumor necrosis factor alpha inhibitor* [tiab] OR tumor necrosis factor alpha antagonist* [tiab] OR tumour necrosis factor alpha inhibitor* [tiab] OR tumour necrosis factor alpha antagonist* [tiab] OR "Infliximab"[Mesh] OR infliximab [tiab] OR Renflexis [tiab] OR Remicade [tiab] OR Inflectra [tiab] OR adalimumab [tiab] OR Humira [tiab] OR amjevita [tiab] OR Cyltezo [tiab] OR "Adalimumab"[Mesh] OR "Etanercept"[Mesh] OR Etanercept [tiab] OR Enbrel [tiab] OR Erelzi [tiab] OR "Certolizumab Pegol"[Mesh] OR certolizumab [tiab] OR certolizumab pegol [tiab] OR Cimzia [tiab] OR "golimumab"[Supplementary Concept] OR golimumab [tiab] OR Simponi [tiab] OR interleukin antagonist* [tiab] "Interleukin 1 Receptor Antagonist Protein"[Mesh] OR anakinra [tiab] OR Kineret [tiab] OR Antril [tiab] OR interleukin 1 receptor antagonist [tiab] OR "canakinumab" [Supplementary Concept] OR canakinumab [tiab] OR ilaris [tiab] OR dupilumab [tiab] OR "dupilumab" [Supplementary Concept] OR Dupixent [tiab] OR secukinumab [tiab] OR "secukinumab" [Supplementary Concept] OR cosentyx [tiab] OR "tocilizumab" [Supplementary Concept] OR tocilizumab [tiab] OR actemra [tiab] OR ustekinumab [tiab] OR "Ustekinumab"[Mesh] OR stelara [tiab] OR ixekizumab [tiab] OR taltz [tiab] OR sarilumab [tiab] OR kevzara [tiab] OR abatacept [tiab] OR "Abatacept"[Mesh] OR belatacept [tiab] OR nulojix [tiab] OR orencia [tiab] OR B-cell inhibitor* [tiab] OR rituximab [tiab] OR "Rituximab"[Mesh] OR Rituxan [tiab] OR Mabthera [tiab] OR rituximab CD20 antibody [tiab] OR belimumab [tiab] OR benlysta [tiab] OR vedolizumab [tiab] OR "vedolizumab" [Supplementary Concept] OR entyvio [tiab] OR "Janus Kinase Inhibitors"[Mesh] OR janus kinase inhibitor* [tiab] OR JAK-inhibitor* [tiab] OR JAK inhibitor* [tiab] OR tofacitinib [tiab] OR xeljanz [tiab] OR upadacitinib [tiab] OR Rinvoq [tiab] OR peficitinib [tiab] OR smyraf [tiab] OR baricitinib [tiab] OR Olumiant [tiab] OR filgotinib [tiab] OR “Biosimilar Pharmaceuticals"[Mesh] OR biosimilar* [tiab] OR "Antibodies, Monoclonal"[Mesh] OR monoclonal antibod* [tiab] OR biologic* [tiab]) AND ("Arthritis, Rheumatoid"[Mesh] OR “rheumatoid arthritis” [tiab]) AND ("Infections"[Mesh] OR infection* [tiab]))

Cochrane:

ID	Search

#1	(TNF-alpha inhibitor):ti,ab,kw (Word variations have been searched)

#2	(TNF-alpha antagonist):ti,ab,kw (Word variations have been searched)

#3	(Tumor necrosis factor alpha inhibitor):ti,ab,kw (Word variations have been searched)

#4	(Tumor necrosis factor alpha antagonist):ti,ab,kw (Word variations have been searched)

#5	MeSH descriptor: [Infliximab] explode all trees

#6	(infliximab):ti,ab,kw (Word variations have been searched)

#7	(renflexis):ti,ab,kw (Word variations have been searched)

#8	(remicade):ti,ab,kw (Word variations have been searched)

#9	(inflectra):ti,ab,kw (Word variations have been searched)

#10	MeSH descriptor: [Adalimumab] explode all trees

#11	(adalimumab):ti,ab,kw (Word variations have been searched)

#12	(humira):ti,ab,kw (Word variations have been searched)

#13	(amjevita):ti,ab,kw (Word variations have been searched)

#14	(cyltezo):ti,ab,kw (Word variations have been searched)

#15	MeSH descriptor: [Etanercept] explode all trees

#16	(etanercept):ti,ab,kw (Word variations have been searched)

#17	(enbrel):ti,ab,kw (Word variations have been searched)

#18	(erelzi):ti,ab,kw (Word variations have been searched)

#19	MeSH descriptor: [Certolizumab Pegol] explode all trees

#20	(certolizumab pegol):ti,ab,kw (Word variations have been searched)

#21	(certolizumab):ti,ab,kw (Word variations have been searched)

#22	(cimzia):ti,ab,kw (Word variations have been searched)

#23	(golimumab):ti,ab,kw (Word variations have been searched)

#24	(simponi):ti,ab,kw (Word variations have been searched)

#25	(interleukin antagonist):ti,ab,kw (Word variations have been searched)

#26	MeSH descriptor: [Interleukin 1 Receptor Antagonist Protein] explode all trees

#27	(anakinra):ti,ab,kw (Word variations have been searched)

#28	(kineret):ti,ab,kw (Word variations have been searched)

#29	(antril):ti,ab,kw (Word variations have been searched)

#30	(interleukin 1 receptor antagonist):ti,ab,kw (Word variations have been searched)

#31	(canakinumab):ti,ab,kw (Word variations have been searched)

#32	(ilaris):ti,ab,kw (Word variations have been searched)

#33	(dupilumab):ti,ab,kw (Word variations have been searched)

#34	(dupixent):ti,ab,kw (Word variations have been searched)

#35	(secukinumab):ti,ab,kw (Word variations have been searched)

#36	(cosentyx):ti,ab,kw (Word variations have been searched)

#37	(tocilizumab):ti,ab,kw (Word variations have been searched)

#38	(actemra):ti,ab,kw (Word variations have been searched)

#39	MeSH descriptor: [Ustekinumab] explode all trees

#40	(ustekinumab):ti,ab,kw (Word variations have been searched)

#41	(stelara):ti,ab,kw (Word variations have been searched)

#42	(ixekizumab):ti,ab,kw (Word variations have been searched)

#43	(taltz):ti,ab,kw (Word variations have been searched)

#44	(sarilumab):ti,ab,kw (Word variations have been searched)

#45	(kevzara):ti,ab,kw (Word variations have been searched)

#46	MeSH descriptor: [Abatacept] explode all trees

#47	(abatacept):ti,ab,kw (Word variations have been searched)

#48	(belatacept):ti,ab,kw (Word variations have been searched)

#49	(nulojix):ti,ab,kw (Word variations have been searched)

#50	(orencia):ti,ab,kw (Word variations have been searched)

#51	(B-cell inhibitor):ti,ab,kw (Word variations have been searched)

#52	MeSH descriptor: [Rituximab] explode all trees

#53	(rituxan):ti,ab,kw (Word variations have been searched)

#54	(rituximab):ti,ab,kw (Word variations have been searched)

#55	(mabthera):ti,ab,kw (Word variations have been searched)

#56	(rituximab CD20 antibody):ti,ab,kw (Word variations have been searched)

#57	(belimumab):ti,ab,kw (Word variations have been searched)

#58	(benlysta):ti,ab,kw (Word variations have been searched)

#59	(vedolizumab):ti,ab,kw (Word variations have been searched)

#60	(entyvio):ti,ab,kw (Word variations have been searched)

#61	MeSH descriptor: [Janus Kinase Inhibitors] explode all trees

#62	(janus kinase inhibitor):ti,ab,kw (Word variations have been searched)

#63	(JAK-inhibitor):ti,ab,kw (Word variations have been searched)

#64	(tofacitinib):ti,ab,kw (Word variations have been searched)

#65	(xeljanz):ti,ab,kw (Word variations have been searched)

#66	(upadacitinib):ti,ab,kw

#67	(rinvoq):ti,ab,kw

#68	(peficitinib):ti,ab,kw

#69	(smyraf):ti,ab,kw

#70	(baricitinib):ti,ab,kw

#71	(olumiant):ti,ab,kw

#72	(filgotinib):ti,ab,kw

#73	MeSH descriptor: [Biosimilar Pharmaceuticals] explode all trees

#74	(biosimilar):ti,ab,kw (Word variations have been searched)

#75	(biologic):ti,ab,kw (Word variations have been searched)

#76	MeSH descriptor: [Antibodies, Monoclonal] explode all trees

#77	(monoclonal antibodies):ti,ab,kw (Word variations have been searched)

#78	(vaccin*):ti,ab,kw

#79	MeSH descriptor: [Arthritis, Rheumatoid] explode all trees

#80	(rheumatoid arthritis):ti,ab,kw (Word variations have been searched)

#81	juvenile

#82	(infection):ti,ab,kw (Word variations have been searched)

#83	MeSH descriptor: [Infections] explode all trees

#84	{OR #1-#77} NOT #78

#85	#79 OR #80 NOT #81

#86	#82 OR #83

#87	#84 AND #85 AND #86

**Appendix 2: flowchart of study in- and exclusion**

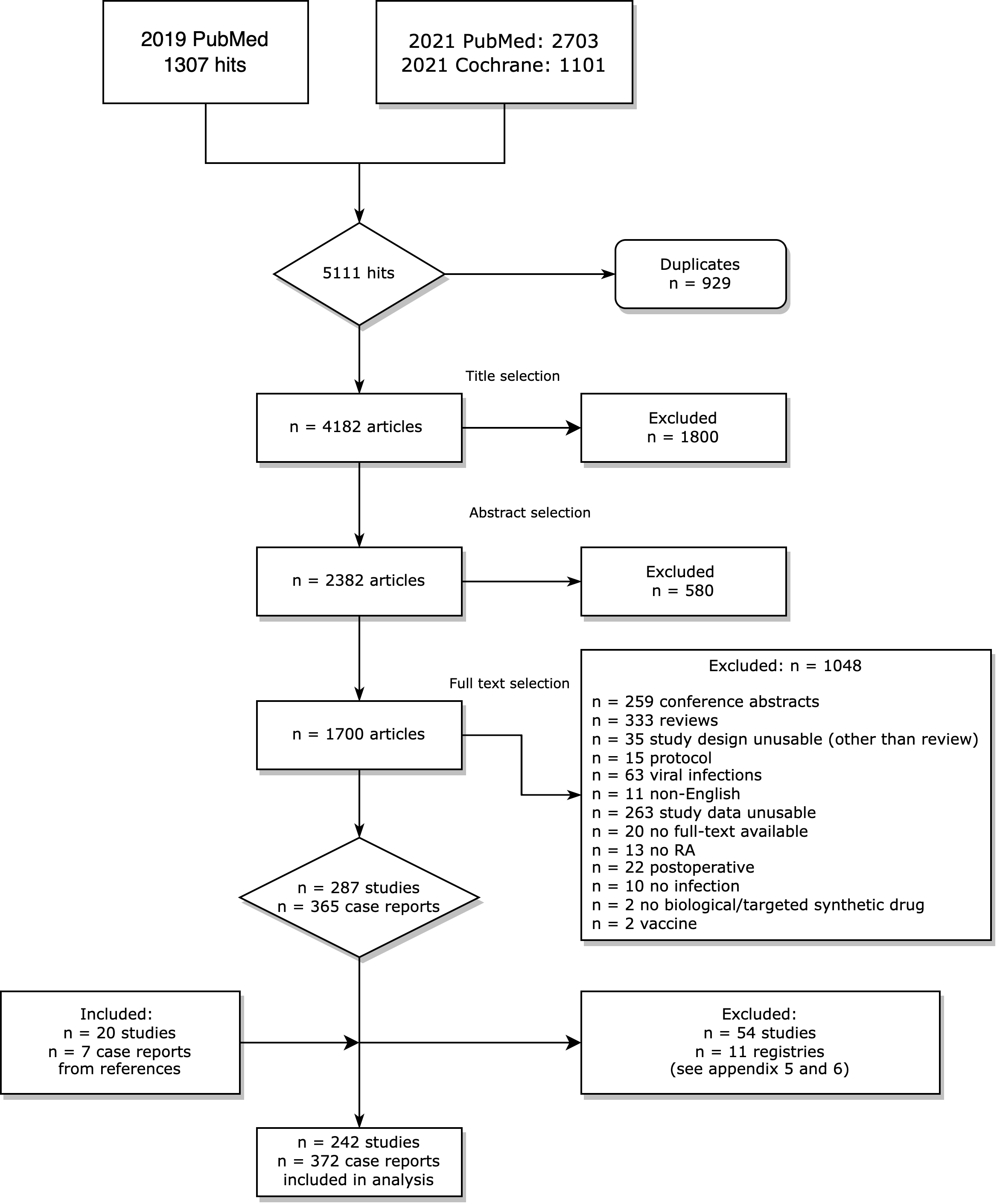

**Appendix 3: database format for the trials database**

In separate excel file

**Appendix 4: database format for the case report database**

In separate excel file

**Appendix 5: Studies making use of the same databases: inclusion, exclusion and reasoning**

Studies making use of the same database were grouped, and decisions were made as to which study (arm) to include in the analyses. Table 1 shows the different groups, decisions and reasoning.

**Table 1. In- and exclusion of studies making use of the same dataset**

| Problem | Solution | PMID | Excluded PMID | Included PMID |
| --- | --- | --- | --- | --- |
| Multiple updates of same prospective cohort with new inclusions and different outcome measurements | We chose the most recent update of the registry concerning TNF-alpha inhibitors. Included were studies that focused on specific infections rather than a total amount of unspecified infections. The study arm on Rituximab from PMDI 28968862 was included as the other studies only used data on TNF-alpha inhibitors. From PMID 20675706 only the serious infection incidence rate was included, since the other studies did not report on that. | 20675706  22532633  21784730  17763441  24140761  28968862 | 17763441  24140761  28968862 TNF-alpha inhibitor study arm | 20675706  22532633  21784730  28968862 Rituximab study arm |
|  | Include studies that analysed biologicals separately, only included from PMID 21791449 the incidence rates of serious infections 21791449 | 16255017  21791449 |  | 16255017  21791449 only incidence rate of Serious infections |
|  | We chose to include the most recent update of the complete registry | 19359261  29329557  31371657  21498482  22422487  24852650  25880658 | 19359261  31371657  21498482  24852650  25880658 study arm on TNF-alpha inhibitors | 29329557  22422487  25880658 study arm on Tocilizumab |
| Registry also used in another study | Use most recent update of registry | 27258623  17324968 | 17324968 Biobadaser study arm | 27258623  17324968 Emecar study arm |
|  | Use most recent update of registry | 30612115  31764977  17261532 | 30612115 Danbio study arm  17261532 | 30612115 Artis study arm  31764977 |
| Same cohort, different biologicals | Include both studies since they focus on different biologicals | 31174592  31777822 |  | 31174592  31777822 |
| Cohorts used in multiple studies with different inclusion periods | Include individual study arms based on most recent update of registry | 17530704  22833479  26315675  28255642  30570833  30679153  30772001 | 17530704  22833479  28255642  30772001  30679153 study arms on Medicare and MarketScan  26315675 study arm on Tocilizumab | 26315675  30570833  30679153 study arms on Tocilizumab and IMS |
| Multiple updates on same prospective cohort | Choose most recent update | 21358439  24470378 | 21358439 | 24470378 |

**Appendix 6: RCTs and extension studies: inclusion, exclusion and reasoning**

RCTs and associated extension studies were grouped according to the common dataset. Per group, the most recent study describing infectious complications from the start of antirheumatic therapy with biological or targeted synthetic treatment was included.

**Table 1. In- and exclusion decisions regarding RCT/OLE clusters**

| Cluster | Source | Include/exclude | Reasoning | PMID | Design |
| --- | --- | --- | --- | --- | --- |
| 1 | Original search | Exclude | Data unsuitable | 23918037 | RCT+OLE |
|  | Original search | Exclude | Data unsuitable | 22596211 | RCT+OLE |
|  | Reference | Exclude | Data unsuitable | 19909548 | RCT |
|  | Original search | Include |  | 18975346 | RCT |
| 2 | Original search | Include |  | 26353833 | RCT+OLE |
|  | Reference | Include |  | 19015207 | RCT |
| 3 | Original search | Exclude | Data unsuitable | 29219638 | RCT+OLE |
|  | Original search | Include |  | 22984173 | RCT |
| 4 | Original search | Exclude | Data unsuitable | 27130908 | RCT+OLE |
|  | Reference | Include |  | 23687260 | RCT |
| 5 | Original search | Exclude | Data unsuitable | 26474452 | RCT+OLE |
|  | Reference | Include |  | 19644849 | RCT |
| 6 | Original search | Exclude | Data unsuitable | 22798265 | RCT+OLE |
|  | Original search | Exclude | Data unsuitable | 17921185 | RCT+OLE |
|  | Original search | Include |  | 16162882 | RCT |
| 7 | Original search | Exclude | Data unsuitable | 28612179 | RCT+OLE |
|  | Reference | Include |  | 27624791 | RCT |
| 8 | Original search | Exclude | Data unsuitable | 29692005 | RCT+OLE |
|  | Reference | Exclude | Data unsuitable | 26909489 | RCT |
| 9 | Original search | Include |  | 26199453 | RCT+OLE |
|  | Reference | Include |  | 12528101 | RCT |
|  | Reference | Exclude | RCT+OLE more recent | 12860493 | RCT |
|  | Original search | Include |  | 14719195 | RCT |
|  | Original search | Include |  | 16308341 | RCT+OLE |
| 10 | Original search | Exclude | Data unsuitable | 27909081 | RCT+OLE |
|  | Original search | Include |  | 19297346 | RCT |
| 11 | Original search | Exclude | Data unsuitable | 25627338 | RCT+OLE |
|  | Reference | Include |  | 19560810 | RCT |
|  | Original search | Exclude | Data unsuitable | 22459542 | RCT+OLE |
| 12 | Original search | Exclude | Data unsuitable | 20444749 | RCT+OLE |
|  | Original search | Include |  | 19066176 | RCT |
|  | Original search | Exclude | Data unsuitable | 23678153 | RCT+OLE |
|  | Original search | Exclude | Data unsuitable | 26669912 | RCT+OLE |
| 13 | Original search | Exclude | Data unsuitable | 23962455 | RCT+OLE |
|  | Reference | Include |  | 23169319 | RCT |
| 14 | Original search | Exclude | Data unsuitable | 24241487 | RCT+OLE |
|  | Original search | Include |  | 16385520 | RCT |
| 15 | Original search | Exclude | Data unsuitable | 24372225 | RCT+OLE |
|  | Original search | Include |  | 24981319 | RCT |
| 16 | Original search | Exclude | Data unsuitable | 31113455 | RCT+OLE |
|  | Reference | Include |  | 29259050 | RCT |
| 17 | Original search | Exclude | Data unsuitable | 25005467 | RCT+OLE |
|  | Original search | Exclude | Data unsuitable | 19273451 | RCT+OLE |
|  | Original search | Include |  | 16052582 | RCT |
| 18 | Original search | Exclude | Data unsuitable | 30922373 | RCT+OLE |
|  | Reference | Include |  | 28584187 | RCT |
| 19 | Original search | Include |  | 31452928 | RCT+OLE |
|  | Reference | Exclude | Data unsuitable | 25733246 | RCT |
| 20 | Original search | Exclude | Data unsuitable | 29657147 | RCT+OLE |
|  | Original search | Exclude | Data unsuitable | 24584926 | RCT+OLE |
|  | Original search | Include |  | 21618201 | RCT |
| 21 | Original search | Include |  | 25834203 | RCT+OLE |
|  | Reference | Exclude | Data unsuitable | 23983039 | RCT |
| 22 | Original search | Exclude | Data unsuitable | 27747585 | RCT+OLE |
|  | Reference | Exclude | Data unsuitable | 23904473 | RCT |
|  | Original search | Include |  | 24942540 | RCT |
| 23 | Original search | Exclude | Data unsuitable | 19019888 | RCT+OLE |
|  | Reference | Include |  | 15188351 | RCT |
| 24 | Original search | Exclude | Data unsuitable | 30299202 | RCT+OLE |
|  | Reference | Include |  | 28622048 | RCT |
| 25 | Original search | Include |  | 24786925 | RCT+OLE |
|  | Original search | Exclude | Data unsuitable | 21893583 | RCT+OLE |
|  | Reference | Exclude | Data unsuitable | 18383390 | RCT+OLE |
|  | Original search | Include |  | 16785475 | RCT |
| 26 | Original search | Exclude | Data unsuitable | 31387417 | RCT+OLE |
|  | Reference | Include |  | 29514803 | RCT |
| 27 | Original search | Exclude | Data unsuitable | 15996057 | RCT+OLE |
|  | Reference | Include |  | 11096165 | RCT |
| 28 | Original search | Exclude | Data unsuitable | 32371430 | RCT+OLE |
|  | Reference | Exclude | Data unsuitable | 31831079 | RCT |
| 29 | Original search | Exclude | Data unsuitable | 25623393 | RCT+OLE |
|  | Original search | Exclude | Data unsuitable | 24001888 | RCT+OLE |
|  | Reference | Include |  | 22661646 | RCT |
| 30 | Original search | Exclude | Data unsuitable | 22089463 | RCT+OLE |
|  | Original search | Include |  | 20131276 | RCT |
| 31 | Original search | Exclude | Data unsuitable | 29361199 | RCT+OLE |
|  | Reference | Include |  | 28213566 | RCT |
|  | Reference | Exclude | Poster |  | RCT |
| 32 | Original search | Include |  | 23027887 | RCT+OLE |
|  | Original search | Include |  | 16947627 | RCT |
| 33 | Original search | Exclude | Data unsuitable | 24429175 | RCT+OLE |
|  | Reference | Include |  | 22730366 | RCT |
| 34 | Original search | Exclude | Data unsuitable | 30997153 | RCT+OLE |
|  | Reference | Include |  | 30053896 | RCT |
| 35 | Original search | Include |  | 24441150 | RCT+OLE |
|  | Original search | Include |  | 23316080 | RCT |
| 36 | Original search | Include |  | 17504821 | RCT+OLE |
|  | Original search | Exclude | Data unsuitable | 17329305 | exclude |
|  | Reference | Exclude | Data unsuitable | 17504821 | exclude |
| 37 | Original search | Exclude | Data unsuitable | 32164762 | RCT+OLE |
|  | Original search | Include |  | 26672064 | RCT |
|  | Original search | Include |  | 31350269 | RCT |
| 38 | Original search | Exclude | Data unsuitable | 18050221 | RCT+OLE |
|  | Original search | Include |  | 15201414 | RCT |
|  | Original search | Exclude | Data unsuitable | 11257159 | RCT |
|  | Original search | Include |  | 16947627 | RCT |
| 39 | Original search | Include |  | 12687534 | RCT |
|  | Original search | Exclude | Data unsuitable | 16396977 | RCT+OLE |
| 40 | Original search | Exclude | Data unsuitable | 29042358 | RCT+OLE |
|  | Original search | Include |  | 26318384 | RCT |
|  | Reference | Exclude | Data unsuitable | 28957563 | RCT+OLE |
| 41 | Original search | Exclude | Data unsuitable | 33164349 | RCT+OLE |
|  | Original search | Include |  | 30590833 | RCT |
| 42 | Original search | Exclude | Data unsuitable | 21821865 | RCT+OLE |
|  | Original search | Include |  | 19124524 | RCT |
| 43 | Original search | Exclude | Data unsuitable | 22596211 | RCT+OLE |
|  | Original search | Include |  | 18975346 | RCT |
| 44 | Original search | Exclude | Data unsuitable | 27334658 | RCT+OLE |
|  | Reference | Include |  | 22873531 | RCT |
| 45 | Original search | Exclude | Data unsuitable | 29439289 | RCT+OLE |
|  | Reference | Include |  | 28950421 | RCT |
| 46 | Original search | Include |  | 24026258 | RCT+OLE |
|  | Original search | Exclude | Data unsuitable | 30666826 | RCT+OLE |
|  | Reference | Include |  | 23348607 | RCT |
|  | Reference | Include |  | 21584942 | RCT |
|  | Reference | Include |  | 25496464 | RCT |
| 47 | Original search | Exclude | Data unsuitable | 26568428 | RCT+OLE |
|  | Reference | Include |  | 22923753 | RCT |
| 48 | Original search | Exclude | Data unsuitable | 29247149 | RCT+OLE |
|  | Original search | Include |  | 24942540 | RCT |
| 49 | Original search | Exclude | Data unsuitable | 31410787 | RCT+OLE |
|  | Original search | Include |  | 28118538 | RCT |
|  | Original search | Include |  | 27748083 | RCT |
| 50 | Original search | Exclude | Data unsuitable | 27006728 | RCT+OLE |
|  | Original search | Include |  | 24356474 | Open label trial |
| 51 | Original search | Include |  | 31287230 | RCT |
|  | Original search | Exclude | Data unsuitable | 33123205 | RCT+OLE |
|  | Reference | Exclude | Data unsuitable | 31362993 | RCT+OLE |
| 52 | Original search | Include |  | 28584187 | RCT |
|  | Original search | Exclude | Data unsuitable | 30922373 | RCT+OLE |
|  | Original search | Exclude | Data unsuitable | 30922373 | RCT+OLE |

*PMID, PubMed ID; RCT, randomised controlled trial; RCT+OLE, randomised controlled trial and open label extension. Source = how the article was found during literature search. Include/exclude = the decision we made to in- or exclude the article in the analysis. Data unsuitable = data in the original article not presented in a way that made inclusion possible, e.g., no mention of infectious adverse events, infectious adverse events not reported per biological class, OLE that did not report adverse events from initial inclusion of patients. Multiple articles from the same group could be included, if they reported infectious adverse events in a different manner (e.g., incidence rate versus the total number of infected patients). Such studies were not included in the same analyses.*

**Appendix 7: composite variables related to infection**

Specific infections that were reported by studies were grouped into composite variables by organ system as specified in table 1.

**Table 1. Infectious adverse event composite variables and individual infections they contain**

| **Composite variable** | **Contains** |
| --- | --- |
| Upper respiratory tract infection | Upper respiratory tract infection |
|  | Nasopharyngitis |
|  | Pharyngitis |
|  | Rhinitis |
|  | Sinusitis |
|  | Bronchitis |
|  | Common cold |
|  | Tonsillitis |
|  | Otitis |
| Lower respiratory tract infection | Lower respiratory tract infection |
|  | Pneumonia |
|  | Empyema |
|  | Viral pneumonia |
|  | Influenza like illness |
|  | Influenza |
|  | PJP |
|  | Pulmonary aspergillosis |
|  | CMV pneumonitis |
|  | Infected bronchiectasis |
| Genitourinary tract infection | Genitourinary tract infection |
|  | Urinary tract infection |
|  | Pyelonephritis |
|  | Epididymitis |
|  | Vaginal infection |
|  | Vaginal candidiasis |
|  | Endometritis |
| Gastrointestinal infection | Gastrointestinal infection |
|  | Gastroenteritis |
|  | Enteritis |
|  | Colitis |
|  | Cholecystitis |
|  | Diverticulitis |
|  | Appendicitis |
|  | Peritonitis |
|  | Gastritis |
|  | Whipple's disease |
|  | Salmonellosis |
| Skin- and soft tissue infections | Skin- and soft tissue infections |
|  | Cellulitis |
|  | Erysipelas |
|  | Abscess |
|  | Wound infection |
|  | Necrotizing fasciitis |
| Bone and joint infections | Bone and joint infections |
|  | Osteomyelitis |
|  | Arthritis |
|  | Bursitis |
|  | Spondylodiscitis |
|  | Prosthetic joint infection |
| Sepsis | Sepsis |
| Mycobacterial infections | TB |
|  | NTM infection |
| Neurological infections | Herpes zoster |
|  | Meningitis |
|  | Encephalitis |
|  | Viral meningitis |
|  | Herpetic encephalitis |
| Fungal infections | Fungal infection |
|  | Mycotic skin infection |
|  | Oral mycosis |
|  | Onychomycosis |
|  | Endocarditis fungal |
|  | Histoplasma infection |
| Viral infections | Viral infection |
|  | Herpes virus infection |
|  | Herpes simplex |
|  | Oral herpes |
|  | Hepatitis A infection |
|  | Hepatitis B infection de novo |
|  | Hepatitis B reactivation |
|  | Hepatitis C infection |
|  | Hepatitis E infection |
|  | Dengue fever |
|  | Varicella infection |
|  | EBV reactivation |
| Other | Dental infection |
|  | Eye infection |
|  | Stomatitis |
|  | Catheter related infection |
|  | Bartonella henselae infection |
|  | Mastitis |
|  | Myositis |
|  | Parasitic infection |
|  | Sialoadenitis |
|  | Borrelia infection |
|  | Hand foot mouth disease |
|  | Lokalised infection |
|  | Extradural abscess |

*PJP, Pneumocystis jirovecii pneumonia; CMV, cytomegalovirus; TB, tuberculosis; NTM, non-tuberculous mycobacteria; EBV, Epstein-Barr virus*

**Appendix 8: Subgroup construction**

We observed a considerable level of heterogeneity between studies included in this review for both the total number of infected patients and the number of seriously infected patients. To create subgroups of relatively homogeneous studies, we grouped studies based on study design and follow-up duration, as explained in Table 1 below.

**Table 1. Theoretical basis for subgroups created in this review**

| **Potential effect modifiers** | **Evaluation** | **Evidence** |
| --- | --- | --- |
| Study design | RCTs comprise a highly selected population, a double-blind design and an intensive follow-up where patients are carefully monitored for every sign of adverse events.  Registries contain real-world data with large study populations; these are secondary data extracted from electronic patient or insurance records. Registries mainly report on serious infections. Patients were not subjected to harsh in- and exclusion criteria like in RCTs, registries therefore contain patients with a higher number of comorbidities than RCTs.  Open-label studies are usually performed in one centre. Patients are subjected to selection before inclusion, the design is not double-blind. | Evans SR. Clinical trial structures. J Exp Stroke Transl Med. 2010 Feb 9;3(1):8-18. doi: 10.6030/1939-067x-3.1.8. PMID: 21423788; PMCID: PMC3059315.    <https://clinicaldevice.typepad.com/cdg_whitepapers/2011/07/registry-studies-why-and-how.html> |
| Follow-up duration | It is likely that the longer a study takes, the more information on ADRs is lost.  The incidence of infections in biological treatment is higher in the first 3-6 months after treatment initiation. | Sakai, R., Komano, Y., Tanaka, M., Nanki, T., Koike, R., … Nagasawa, H. (2012). Time-dependent increased risk for serious infection from continuous use of TNF antagonists during three years in rheumatoid arthritis patients. Arthritis Care & Research, n/a–n/a. doi:10.1002/acr.21666  Strangfeld, A., Eveslage, M., Schneider, M., Bergerhausen, H. J., Klopsch, T., Zink, A., & Listing, J. (2011). Treatment benefit or survival of the fittest: what drives the time-dependent decrease in serious infection rates under TNF inhibition and what does this imply for the individual patient? Annals of the Rheumatic Diseases, 70(11), 1914–1920. doi:10.1136/ard.2011.151043  Ranza, R., de la Vega, M.C., Laurindo, I.M.M. et al. Changing rate of serious infections in biologic-exposed rheumatoid arthritis patients. Data from South American registries BIOBADABRASIL and BIOBADASAR. Clin Rheumatol 38, 2129–2139 (2019). https://doi.org/10.1007/s10067-019-04516-2 |

*ADR – adverse drug reaction; RCT – randomised controlled trial*

As presented in the manuscript, we assessed how heterogeneity metrics changed as we formed subgroups. We first evaluated the relationship between follow-up duration and infection prevalence, by incorporating the follow-up duration as a moderator in a multivariate meta-regression model. I^2^, i.e., the percentage of the total variability in a set of effect sizes due to true heterogeneity, was used as primary criterion to assess the change in the level of heterogeneity. The $\sigma_{1}^{2}$ and $\sigma_{2}^{2}$, being the between-study and between-study-arms variance respectively, were used to support our I^2^ based decision as subgroups were created. Cochran’s Q is a weighted sum of squares that uses the deviation of each study’s observed effect from the summary effect, weighted by the inverse of the study’s variance.

First, we computed the heterogeneity metrics for all studies taken together, then, studies were divided by study design, and finally also by follow-up duration (please see Figure 2 in the manuscript).

All subgroups based on study design – except the registry subgroup (which is based on only three studies) – showed significant heterogeneity. We therefore formed subgroups based on study design and follow-up duration after exploring how the follow-up duration influences the proportion of infected patients (Table 2, Figures 1 & 2). Following the patterns observed in Figures 2 and 3, we defined three subgroups: £18 weeks, 19-38 weeks, and 39-60 weeks. Because of the small number of studies per study design, only RCT studies were further subgrouped according to follow-up duration.

Although the level of heterogeneity was reduced subsequently after subgroups were formed, a moderate to higher level of heterogeneity that needs to be explained or controlled (if possible) remained. For that, we employed a multivariate meta-regression model (see Appendices 10, 19 and 20).

**Table 2: Multivariate meta-regression results using follow-up duration as moderator (RCTs only)**

|  | | $\beta$ (SE) | Test for moderator | $\sigma_{1}^{2}$ | $\sigma_{2}^{2}$ | $I^{2}$ | Cochran's Q test (P) |
| --- | --- | --- | --- | --- | --- | --- | --- |
| Total number of infected patients | Intercept | -0.9426(0.0850) | <0.0001 | 0.2210 | 0.0035 | 87.7992 | $< 0.001$ |
|  | Follow up length  (weeks) | 0.0106 (0.0023) | <0.0001 |  |  |  |  |
| Number of seriously infected patients | Intercept | -3.8412(0.0948) | <0.001 | 0.1322 | 0.0285 | 36.3202 | $0.0218$ |
|  | Follow up  (weeks) | 0.0054(0.0023) | <0.001 |  |  |  |  |

$\beta$*, meta-regression coefficient estimate; SE, Standard error;* $I^{2},$*variation (%) explained by differences in studies;* $\sigma_{1}^{2}$*, variance (between study);* $\sigma_{2}^{2},$ $variance (within study)$*; Cochran's Q test (P), Cochran's Q test p-value.*

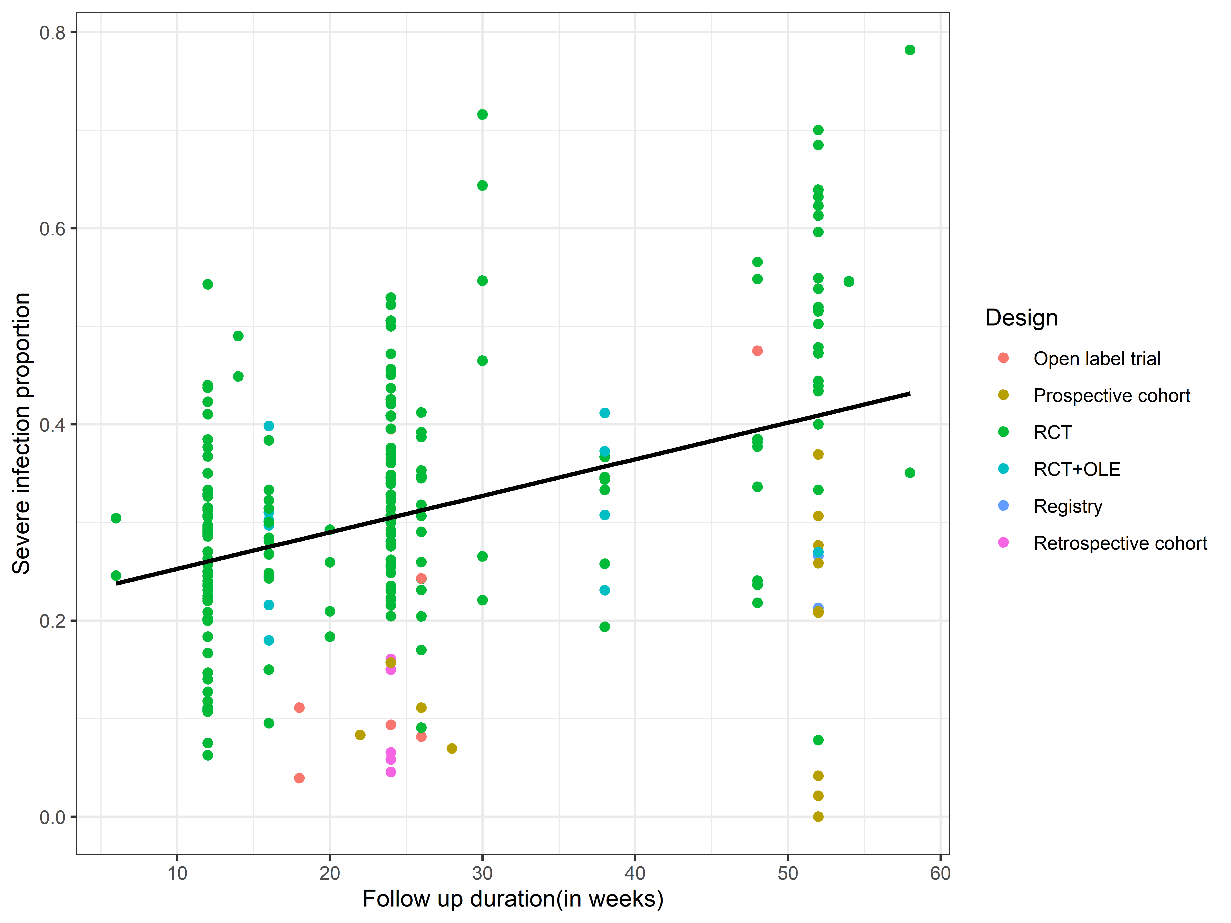

**Figure 1: Infection proportion of total number of infected** **patients vs follow-up duration.**

*RCT, randomised controlled trial; RCT+OLE = randomised controlled trial open-label extension*

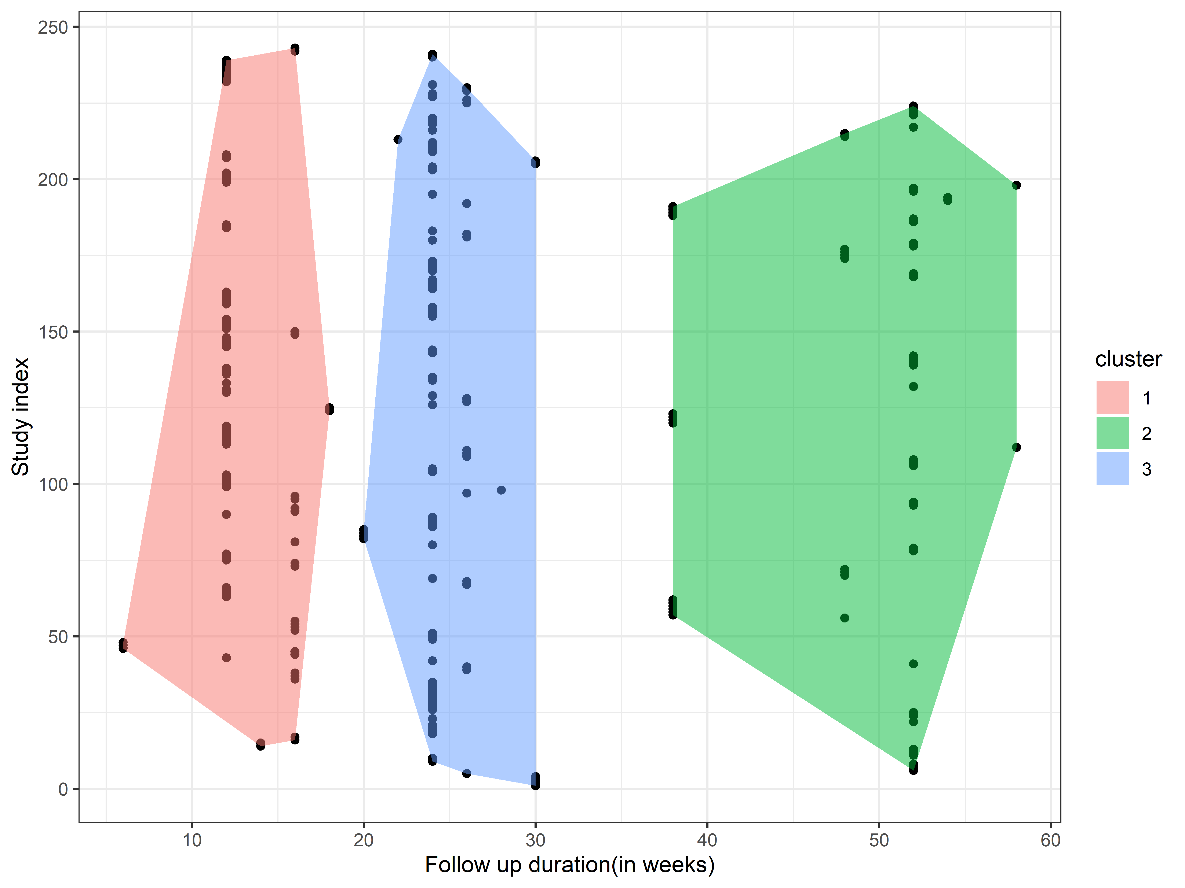

**Figure 2: clusters in the follow-up length of included studies**

**Specifying inputs to the rma.mv() function**

We used the *rma.mv* function of the *metafor package* with the input of the observed effect sizes$(y_{i}$) and the corresponding sampling variances$(v_{i})$. For the proportion of the total(seriously) infected patients, for the $i^{th}$study $\left( i=1,\ldots,n \right)$, the observed effect size was $log[\frac{\hat{p}_{i}}{1-\hat{p}_{i}}]$ and the corresponding sampling variance was $\frac{1}{x_{i}}+\frac{1}{L_{i}-x_{i}}$, where $\hat{p}_{i}$ = the reported proportion of the total(seriously) infected patients, $x_{i}$ = the total number of (seriously) infected patients, and $L_{i}$ is the sample size of the $i^{th}$study, $n$ = the number of studies/study arms included in this review. For the event rates, the observed effect size and the corresponding sampling variance were $\frac{e_{i}}{t_{i}}$ and $\frac{e_{i}}{t_{i}^{2}}$ respectively, where $e_{i}$ = the (total) number of (seriously) infectious events and $t_{i}$ = the total exposure time in patient-year in the $i^{th}$ study. In the *rma.mv* function, we specified the random effect as ~ 1| study_id/study_arm_id where study_id is the unique identifying number for each study and study_arm_id is the unique identifying number for each study arms within the study. This specification of the random effect allows the effect size to vary within and between study arms and by studies.

**Appendix 9: Variable selection**

A multivariate meta-regression model was fitted for the three subgroups of RCT studies (grouped by study design and follow-up duration, see figure 1 in appendix 8). We identified 51 potential moderators that have a theoretical basis (literature supported) and that were also extractable from the studies included in this review. From these, we selected only those features that had at least been reported in 10 studies per subgroup (table 1). The estimated coefficient together with other relevant information for the selected moderators are presented in the result section of the manuscript, and appendix 10.

**Table 1. Variables selected for the multivariate meta-regression model**

| Variable | Reference(s) | Included/excluded | Reasoning |
| --- | --- | --- | --- |
| Age | KOMANO, Y., TANAKA, M., NANKI, T., KOIKE, R., SAKAI, R., … KAMEDA, H. (2011). Incidence and Risk Factors for Serious Infection in Patients with Rheumatoid Arthritis Treated with Tumor Necrosis Factor Inhibitors: A Report from the Registry of Japanese Rheumatoid Arthritis Patients for Longterm Safety. The Journal of Rheumatology, 38(7), 1258–1264. doi:10.3899/jrheum.101009 | Include age | Increasing age increases the risk of serious infection |
| Sex | Bechman, K., Halai, K., Yates, M., Norton, S., Cope, A. P., British Society for Rheumatology Biologics Register for Rheumatoid Arthritis Contributors Group, Hyrich, K. L., & Galloway, J. B. (2021). Nonserious Infections in Patients With Rheumatoid Arthritis: Results From the British Society for Rheumatology Biologics Register for Rheumatoid Arthritis. Arthritis & rheumatology (Hoboken, N.J.), 73(10), 1800–1809. https://doi.org/10.1002/art.41754 | Include sex | Does not appear to influence the risk of serious infection, but does influence the risk of non-serious infection (female sex) |
| Chronic pulmonary diseases: COPD, Asthma, ILD | KOMANO, Y., TANAKA, M., NANKI, T., KOIKE, R., SAKAI, R., … KAMEDA, H. (2011). Incidence and Risk Factors for Serious Infection in Patients with Rheumatoid Arthritis Treated with Tumor Necrosis Factor Inhibitors: A Report from the Registry of Japanese Rheumatoid Arthritis Patients for Longterm Safety. The Journal of Rheumatology, 38(7), 1258–1264. doi:10.3899/jrheum.101009 | Include COPD, Asthma, ILD | Presence of chronic pulmonary diseases increases the risk of serious infection, particularly pneumonia |
| Advanced disease stage of RA (steinbroker stage III or IV) | KOMANO, Y., TANAKA, M., NANKI, T., KOIKE, R., SAKAI, R., … KAMEDA, H. (2011). Incidence and Risk Factors for Serious Infection in Patients with Rheumatoid Arthritis Treated with Tumor Necrosis Factor Inhibitors: A Report from the Registry of Japanese Rheumatoid Arthritis Patients for Longterm Safety. The Journal of Rheumatology, 38(7), 1258–1264. doi:10.3899/jrheum.101009 | Include Steinbroker stage | Advanced disease stage of RA increases the risk of serious infection |
| Dosage of MTX, particularly > 8 mg/week | Sakai, R., Komano, Y., Tanaka, M., Nanki, T., Koike, R., … Nagasawa, H. (2012). Time-dependent increased risk for serious infection from continuous use of TNF antagonists during three years in rheumatoid arthritis patients. Arthritis Care & Research, n/a–n/a. doi:10.1002/acr.21666 | Include MTX dosage | Higher MTX dosage increases the risk of infection |
| Biological class/target/name | Singh, J. A., Wells, G. A., Christensen, R., Tanjong Ghogomu, E., Maxwell, L. J., MacDonald, J. K., … Buchbinder, R. (2011). Adverse effects of biologics: a network meta-analysis and Cochrane overview. Cochrane Database of Systematic Reviews. doi:10.1002/14651858.cd008794.pub2  10.1002/14651858.CD008794.pub2  Strangfeld, A., Eveslage, M., Schneider, M., Bergerhausen, H. J., Klopsch, T., Zink, A., & Listing, J. (2011). Treatment benefit or survival of the fittest: what drives the time-dependent decrease in serious infection rates under TNF inhibition and what does this imply for the individual patient? Annals of the Rheumatic Diseases, 70(11), 1914–1920. doi:10.1136/ard.2011.151043  Bechman, K., Halai, K., Yates, M., Norton, S., Cope, A. P., British Society for Rheumatology Biologics Register for Rheumatoid Arthritis Contributors Group, Hyrich, K. L., & Galloway, J. B. (2021). Nonserious Infections in Patients With Rheumatoid Arthritis: Results From the British Society for Rheumatology Biologics Register for Rheumatoid Arthritis. Arthritis & rheumatology (Hoboken, N.J.), 73(10), 1800–1809. https://doi.org/10.1002/art.41754 | Include biological class, target and individual drugs | Biological class/target/name may increase or decrease the risk of contracting a serious infection. TNF-alpha inhibitors seem more likely to cause serious infections than other biological classes. Etanercept seems less likely to cause serious infection, and infliximab/adalimumab/certolizumab/tocilizumab/anakinra seem more likely to do so  Also influences risk of nonserious infections |
| Disease activity   - Mean DAS28-CRP - Mean CDAI - HAQ - TJC28 - VAS pain score   Functional status FFbH | Sakai, R., Komano, Y., Tanaka, M., Nanki, T., Koike, R., … Nagasawa, H. (2012). Time-dependent increased risk for serious infection from continuous use of TNF antagonists during three years in rheumatoid arthritis patients. Arthritis Care & Research, n/a–n/a. doi:10.1002/acr.21666  Au, K., Reed, G., Curtis, J. R., Kremer, J. M., Greenberg, J. D., … Strand, V. (2011). High disease activity is associated with an increased risk of infection in patients with rheumatoid arthritis. Annals of the Rheumatic Diseases, 70(5), 785–791. doi:10.1136/ard.2010.128637  Van Dartel, S. A. A., Fransen, J., Kievit, W., Dutmer, E. A. J., Brus, H. L. M., Houtman, N. M., … van Riel, P. L. C. M. (2013). Predictors for the 5-year risk of serious infections in patients with rheumatoid arthritis treated with anti-tumour necrosis factor therapy: a cohort study in the Dutch Rheumatoid Arthritis Monitoring (DREAM) registry. Rheumatology, 52(6), 1052–1057. doi:10.1093/rheumatology/kes413  10.1093/rheumatology/kes413  Bechman, K., Halai, K., Yates, M., Norton, S., Cope, A. P., British Society for Rheumatology Biologics Register for Rheumatoid Arthritis Contributors Group, Hyrich, K. L., & Galloway, J. B. (2021). Nonserious Infections in Patients With Rheumatoid Arthritis: Results From the British Society for Rheumatology Biologics Register for Rheumatoid Arthritis. Arthritis & rheumatology (Hoboken, N.J.), 73(10), 1800–1809. https://doi.org/10.1002/art.41754 | Include DAS-28 CRP, CDAI, HAQ, TJC28, VAS pain score separately  No FFbH included in database | Higher disease activity may lead to higher infection risk, however, data are conflicting  Increasing functional status is associated with decreased infection risk  DAS28 and HAQ-DI also influence risk of nonserious infection |
| DAS-28 ESR  TJC68  SJC28  SJC66  Physicians VAS  Patient’s VAS  FACIT  SDAI  mTSS  Morning stiffness duration  SF-36 |  | Include composite variable of DAS28-CRP, DAS28-ESR, CDAI, TJC28, TJC68, SJC28, SJC66, Physicians VAS, Patient’s VAS, Pain VAS, FACIT | DAS-28 CRP and TJC28 are associated with increased infection risk. By extension, DAS-28 ESR and TJC68 could be predictors of infection risk. Swollen joint count and physycians and patients VAS are also important measures of disease activity and should be included separately. We also included a composite disease activity variable |
| Chronic renal disease | Strangfeld, A., Eveslage, M., Schneider, M., Bergerhausen, H. J., Klopsch, T., Zink, A., & Listing, J. (2011). Treatment benefit or survival of the fittest: what drives the time-dependent decrease in serious infection rates under TNF inhibition and what does this imply for the individual patient? Annals of the Rheumatic Diseases, 70(11), 1914–1920. doi:10.1136/ard.2011.151043 | Include  renal disease | Presence of chronic renal disease increases the risk of serious infection |
| History of serious infections | Strangfeld, A., Eveslage, M., Schneider, M., Bergerhausen, H. J., Klopsch, T., Zink, A., & Listing, J. (2011). Treatment benefit or survival of the fittest: what drives the time-dependent decrease in serious infection rates under TNF inhibition and what does this imply for the individual patient? Annals of the Rheumatic Diseases, 70(11), 1914–1920. doi:10.1136/ard.2011.151043 | Include history of serious infections | A history of serious infections increases the risk of serious infections during the study period |
| Baseline ESR | Favalli, E. G., Desiati, F., Atzeni, F., Sarzi-Puttini, P., Caporali, R., Pallavicini, F. B., … Marchesoni, A. (2009). Serious infections during anti-TNFα treatment in rheumatoid arthritis patients. Autoimmunity Reviews, 8(3), 266–273. doi:10.1016/j.autrev.2008.11.002  10.1016/j.autrev.2008.11.002 | Include baseline ESR | Increasing ESR correlates with increasing risk of serious infection |
| Concomitant use of corticosteroids and dose-dependent risk  Mean dosage of prednisone >10 mg/day (“high-dose corticosteroids”) | Favalli, E. G., Desiati, F., Atzeni, F., Sarzi-Puttini, P., Caporali, R., Pallavicini, F. B., … Marchesoni, A. (2009). Serious infections during anti-TNFα treatment in rheumatoid arthritis patients. Autoimmunity Reviews, 8(3), 266–273. doi:10.1016/j.autrev.2008.11.002  10.1016/j.autrev.2008.11.002  Wolfe, F., Caplan, L., & Michaud, K. (2006). Treatment for rheumatoid arthritis and the risk of hospitalization for pneumonia: Associations with prednisone, disease-modifying antirheumatic drugs, and anti–tumor necrosis factor therapy. Arthritis & Rheumatism, 54(2), 628–634. doi:10.1002/art.21568  Sakai, R., Komano, Y., Tanaka, M., Nanki, T., Koike, R., … Nagasawa, H. (2012). Time-dependent increased risk for serious infection from continuous use of TNF antagonists during three years in rheumatoid arthritis patients. Arthritis Care & Research, n/a–n/a. doi:10.1002/acr.21666  Schneeweiss, S., Setoguchi, S., Weinblatt, M. E., Katz, J. N., Avorn, J., Sax, P. E., … Solomon, D. H. (2007). Anti–tumor necrosis factor α therapy and the risk of serious bacterial infections in elderly patients with rheumatoid arthritis. Arthritis & Rheumatism, 56(6), 1754–1764. doi:10.1002/art.22600  Strangfeld, A., Eveslage, M., Schneider, M., Bergerhausen, H. J., Klopsch, T., Zink, A., & Listing, J. (2011). Treatment benefit or survival of the fittest: what drives the time-dependent decrease in serious infection rates under TNF inhibition and what does this imply for the individual patient? Annals of the Rheumatic Diseases, 70(11), 1914–1920. doi:10.1136/ard.2011.151043  Bechman, K., Halai, K., Yates, M., Norton, S., Cope, A. P., British Society for Rheumatology Biologics Register for Rheumatoid Arthritis Contributors Group, Hyrich, K. L., & Galloway, J. B. (2021). Nonserious Infections in Patients With Rheumatoid Arthritis: Results From the British Society for Rheumatology Biologics Register for Rheumatoid Arthritis. Arthritis & rheumatology (Hoboken, N.J.), 73(10), 1800–1809. https://doi.org/10.1002/art.41754 | Include concomitant corticosteroid  Include corticosteroid dose | Concomitant use of corticosteroids and their increasing dosage both increase the risk of serious infection  Use of corticosteroids increases the risk of non-serious infection |
| Diabetes mellitus | Curtis, J. R., Xie, F., Chen, L., Muntner, P., Grijalva, C. G., Spettell, C., Fernandes, J., McMahan, R. M., Baddley, J. W., Saag, K. G., Beukelman, T., & Delzell, E. (2012). Use of a disease risk score to compare serious infections associated with anti-tumor necrosis factor therapy among high- versus lower-risk rheumatoid arthritis patients. Arthritis care & research, 64(10), 1480–1489. <https://doi.org/10.1002/acr.21805>  Crowson, C. S., Hoganson, D. D., Fitz-Gibbon, P. D., & Matteson, E. L. (2012). Development and validation of a risk score for serious infection in patients with rheumatoid arthritis. Arthritis and rheumatism, 64(9), 2847–2855. https://doi.org/10.1002/art.34530 | Include diabetes mellitus | Presence of diabetes mellitus increases the risk of serious infection |
| Several other comorbidities:   - Alcoholism - Coronary heart disease - Peripheral vascular disease | Crowson, C. S., Hoganson, D. D., Fitz-Gibbon, P. D., & Matteson, E. L. (2012). Development and validation of a risk score for serious infection in patients with rheumatoid arthritis. Arthritis and rheumatism, 64(9), 2847–2855. https://doi.org/10.1002/art.34530 | Include coronary heart disease, include congestive heart failure  Include alcoholism  Include peripheral vascular disease | Presence of these comorbidities increases the risk of serious infection |
| Smoking | Bechman, K., Halai, K., Yates, M., Norton, S., Cope, A. P., … Hyrich, K. L. (2021). *Nonserious Infections in Patients With Rheumatoid Arthritis: Results From the British Society for Rheumatology Biologics Register for Rheumatoid Arthritis. Arthritis & Rheumatology.* doi:10.1002/art.41754 | Include smoking status (current, former, ever) | Smoking seems to be associated with an increased risk of serious infection, but a decreased risk of nonserious infection |
| Extra-articular manifestations of RA, organic brain disease | Doran, M. F., Crowson, C. S., Pond, G. R., O'Fallon, W. M., & Gabriel, S. E. (2002). Predictors of infection in rheumatoid arthritis. Arthritis and rheumatism, 46(9), 2294–2300. https://doi.org/10.1002/art.10529 | Include extra-articular manifestations of RA  Organic brain disease not included in database | Extra-articular manifestations of RA and organic brain disease are associated with increasing risk of serious infection |
| RA duration | Lang, V. R., Englbrecht, M., Rech, J., Nüsslein, H., Manger, K., Schuch, F., … Zwerina, J. (2011). Risk of infections in rheumatoid arthritis patients treated with tocilizumab. Rheumatology, 51(5), 852–857. doi:10.1093/rheumatology/ker223  10.1093/rheumatology/ker223 | Include disease duration | Longer RA duration is associated with increased risk of serious infection |
| Previous exposure to >3 DMARDs | Lang, V. R., Englbrecht, M., Rech, J., Nüsslein, H., Manger, K., Schuch, F., … Zwerina, J. (2011). Risk of infections in rheumatoid arthritis patients treated with tocilizumab. Rheumatology, 51(5), 852–857. doi:10.1093/rheumatology/ker223  10.1093/rheumatology/ker223 | Include no of prior DMARDs 0->3  Include mean number of prior DMARDs | Exposure to 3 or more prior DMARDS is associated with an increased risk of serious infection |
| Concomitant leflunomide use | Lang, V. R., Englbrecht, M., Rech, J., Nüsslein, H., Manger, K., Schuch, F., … Zwerina, J. (2011). Risk of infections in rheumatoid arthritis patients treated with tocilizumab. Rheumatology, 51(5), 852–857. doi:10.1093/rheumatology/ker223  10.1093/rheumatology/ker223  Morel, J., Constantin, A., Baron, G., Dernis, E., Flipo, R. M., Rist, S., … Sibilia, J. (2017). Risk factors of serious infections in patients with rheumatoid arthritis treated with tocilizumab in the French Registry REGATE. Rheumatology, 56(10), 1746–1754. doi:10.1093/rheumatology/kex238  10.1093/rheumatology/kex238 | Include percentage of patients on concomitant leflunomide | Concomitant leflunomide use is associated with an increased risk of serious infections in tocilizumab use |
| Negative anti-CCP | Morel, J., Constantin, A., Baron, G., Dernis, E., Flipo, R. M., Rist, S., … Sibilia, J. (2017). Risk factors of serious infections in patients with rheumatoid arthritis treated with tocilizumab in the French Registry REGATE. Rheumatology, 56(10), 1746–1754. doi:10.1093/rheumatology/kex238  10.1093/rheumatology/kex238 | Include percentage of patients with positive anti-CCP | Negative anti-CCP at baseline is associated with an increased risk of serious infection in tocilizumab use |
| Baseline B-cell count, IgM, IgG, neutrophils |  | Include | Decreased B-cell and immunoglobulin counts at baseline are associated with increased risk of serious infection. |
| Prior bDMARD use | Salmon JH, Gottenberg JE, Ravaud P on behalf of all the investigators of the ORA registry and the French Society of Rheumatology, et alPredictive risk factors of serious infections in patients with rheumatoid arthritis treated with abatacept in common practice: results from the Orencia and Rheumatoid Arthritis (ORA) registryAnnals of the Rheumatic Diseases 2016;75:1108-1113. | Include | A lower number of prior bDMARDs seems to increase the risk of serious infection |
| Mean number of prior bDMARs | Salmon JH, Gottenberg JE, Ravaud P on behalf of all the investigators of the ORA registry and the French Society of Rheumatology, et alPredictive risk factors of serious infections in patients with rheumatoid arthritis treated with abatacept in common practice: results from the Orencia and Rheumatoid Arthritis (ORA) registryAnnals of the Rheumatic Diseases 2016;75:1108-1113. | Include | A lower number of prior bDMARDs seems to increase the risk of serious infection |
| Number of prior bDMARDs 0->3 | Salmon JH, Gottenberg JE, Ravaud P on behalf of all the investigators of the ORA registry and the French Society of Rheumatology, et alPredictive risk factors of serious infections in patients with rheumatoid arthritis treated with abatacept in common practice: results from the Orencia and Rheumatoid Arthritis (ORA) registryAnnals of the Rheumatic Diseases 2016;75:1108-1113. | Include | A lower number of prior bDMARDs seems to increase the risk of serious infection |

*Anti-CCP, anti-cyclic citrullinated protein antibodies; bDMARD, biological DMARD; CDAI, clinical disease activity index; COPD, chronic obstructive pulmonary disease; CRP, c-reactive protein; ILD, interstitial lung disease; DAS28-CRP, disease activity score in 28 joints using CRP; DAS28-ESR, disease activity score in 28 joints using ESR; DMARD, disease modifying anti-rheumatic drug; ESR, erythrocyte sedimentation rate; FACIT, functional assessment of chronic illness therapy-fatigue; FFbH, Funktionsfragebogen Hannover; HAQ-DI, health assessment questionnaire-disability index; mTSS, modified total Sharp score; MTX, methotrexate; RA, rheumatoid arthritis; SDAI, simplified disease activity index; SF-36, 36-item short form survey; SJC-28/66, swollen joint count in 28 or 66 joints; TJC,-28/68 tender joint count in 28 or 68 joints; VAS, visual analogue scale. Included/excluded: the decision whether the variable was included in the analysis, based on previously published literature. Included variables were analysed if they were reported in at least 10 studies.*

**Appendix 10: Multivariate meta-regression**

This section presents the result of the multivariate meta-regression analysis for the total number of infected patients and the number of seriously infected patients for studies grouped by study design.

**Table 1. Multivariate meta-regression results for the total number of infected patients; RCT studies**

| Moderators | N | $\boldsymbol{\beta}\mathbf{(}\boldsymbol{SE}\mathbf{)}$ | 95%CI | Cochran's Q test (P) | Test for moderator (P) | $\boldsymbol{\sigma}_{\mathbf{1}}^{\mathbf{2}}$ | $\boldsymbol{\sigma}_{\mathbf{2}}^{\mathbf{2}}$ | $\mathbf{I}^{\mathbf{2}}$ |
| --- | --- | --- | --- | --- | --- | --- | --- | --- |
| Biological target (reference: TNF alpha inhibitor) | 203 |  |  | <0.001 | **0.048** | 0.26 | 0.002 | 89.41 |
| Constant |  | -0.62(0.07) | [-0.77, -0.48] |  |  |  |  |  |
| B-cell inhibitor |  | 0.05(0.16) | [-0.26, 0.36] |  |  |  |  |  |
| Rituximab |  | -0.13(0.25) | [-0.61, 0.36] |  |  |  |  |  |
| BTK-inhibitor |  | -0.91(0.28) | [-1.46, -0.36] |  |  |  |  |  |
| IL-17 inhibitor |  | -0.48(0.39) | [-1.24, 0.28] |  |  |  |  |  |
| IL-6 inhibitor |  | 0.08(0.13) | [-0.17, 0.33] |  |  |  |  |  |
| JAK1/2 inhibitor |  | 0.17(0.13) | [-0.09, 0.43] |  |  |  |  |  |
| JAK1/3inhibitor |  | -0.43(0.32) | [-1.07, 0.21] |  |  |  |  |  |
| JAK1 inhibitor |  | 0.11(0.13) | [-0.15, 0.38] |  |  |  |  |  |
| Pan-JAKinhibitor |  | -0.43(0.54) | [-1.49, 0.63] |  |  |  |  |  |
| T-cell inhibitor |  | 0.09(0.12) | [-0.15, 0.33] |  |  |  |  |  |
| Biological class (reference: TNF alpha inhibitor) |  |  |  | <0.001 | **0.0310** | 0.27 | 0.002 | 89.60 |
| Constant |  | -0.65( 0.07) | [-0.79, -0.51] |  |  |  |  |  |
| Interleukin antagonists |  | 0.06(0.12) | [-0.18, 0.30] |  |  |  |  |  |
| JAK-inhibitor |  | 0.10( 0.09) | [-0.08, 0.28] |  |  |  |  |  |
| Other |  | -0.90( 0.28) | [-1.45, -0.35] |  |  |  |  |  |
| B-cell inhibitor |  | 0.02( 0.14) | [-0.25, 0.30] |  |  |  |  |  |
| T-cell inhibitor |  | 0.09(0.12) | [-0.14, 0.33] |  |  |  |  |  |
| Follow up duration (weeks) | 211 | 0.01(0.002) | [0.01, 0.02] | <0.001 | **<.0001** | 0.22 | 0.004 | 89.28 |
| Age (mean) | 192 | -0.005(0.01) | [-0.03, 0.02] | <0.001 | 0.72 | 0.26 | 0.004 | 89.42 |
| MTX dose (mg/week) (mean) | 101 | 0.02(0.01) | [-0.01, 0.05] | <0.001 | 0.16 | 0.22 | 0.002 | 88.59 |
| DAS28-CRP (mean) | 131 | -0.0001(0.09) | [-0.17, 0.17] | <0.001 | 1.0 | 0.17 | 0.002 | 83.64 |
| CDAI (mean) | 51 | -0.01(0.01) | [-0.03, 0.02] | <0.001 | 0.53 | 0.11 | 0.003 | 73.24 |
| HAQ-DI (mean) | 175 | -0.01(0.01) | [-0.02, 0.01] | <0.001 | 0.53 | 0.24 | 0.004 | 88.93 |
| TJC-28 (mean) | 38 | -0.05(0.03) | [-0.11, 0.02] | <0.001 | 0.15 | 0.14 | 0.01 | 78.82 |
| Pain VAS (mean) | 107 | 0.01(0.01) | [-0.01, 0.03] | <0.001 | 0.47 | 0.20 | 0.01 | 87.98 |
| Exclusion serious/recurrent infection | 208 | -0.05(0.10) | [-0.25, 0.16] | <0.001 | 0.65 | 0.27 | 0.004 | 89.27 |
| ESR (mm/h) (mean) | 85 | -0.004(0.011) | [-0.03, 0.02] | <0.001 | 0.68 | 0.25 | 0.01 | 89.38 |
| Corticosteroid use (percent) | 135 | -0.002(0.003) | [-0.01, 0.004] | <0.001 | 0.56 | 0.23 | 0.004 | 90.15 |
| Corticosteroid dose (mg/day) (mean) | 40 | -0.004(0.07) | [-0.13, 0.13] | <0.001 | 0.95 | 0.19 | 0 | 88.49 |
| RA duration (years) (mean) | 173 | -0.005(0.01) | [-0.03, 0.02] | <0.001 | 0.74 | 0.20 | 0.003 | 87.85 |
| 3 or more prior DMARDs (percent) | 15 | 0.0003(0.01) | [-0.03, 0.03] | <0.001 | 0.98 | 0.30 | 0.02 | 93.56 |
| Number of prior DMARDs (mean) | 26 | 0.14(0.17) | [-0.19, 0.46] | <0.001 | 0.41 | 0.20 | 0.02 | 89.62 |
| Number of prior DMARDs (SD) | 19 | 0.1(0.39) | [-0.67, 0.87] | <0.001 | 0.80 | 0.22 | 0.02 | 91.56 |
| Leflunomide use (percent) | 15 | 0.09(0.04) | [0.02, 0.17] | <0.001 | **0.01** | 0.03 | 0.02 | 81.26 |
| Anti-CCP positive (percent) | 121 | 0.003(0.003) | [-0.002, 0.01] | <0.001 | 0.26 | 0.24 | 0.003 | 86.73 |
| Previous bDMARD use (percent) | 140 | 0(0.0013) | [-0.002, 0.002] | <0.001 | 0.99 | 0.21 | 0 | 87.05 |
| 1 previous bDMARD (percent) | 24 | -0.012(0.004) | [-0.02, -0.003] | <0.001 | **0.01** | 0.06 | 0 | 65.93 |
| 2 previous bDMARD (percent) | 24 | 0.01(0.011) | [-0.011, 0.03] | <0.001 | 0.36 | 0.10 | 0 | 76.28 |
| 3 or more previous bDMARD (percent) | 15 | -0.002(0.015) | [-0.03, 0.03] | <0.001 | 0.92 | 0.11 | 0 | 79.57 |
| Disease activity composite variable | 194 | -0.15(0.17) | [-0.48, 0.18] | <0.001 | 0.37 | 0.25 | 0.003 | 89.27 |
| Exclusion criteria composite variable | 211 | 0.012(0.03) | [-0.05, 0.07] | <0.001 | 0.71 | 0.27 | 0.004 | 89.91 |

*Anti-CCP, anti-citrullinated protein antibodies; BTK, Bruton tyrosine kinase; CDAI = clinical disease activity index; CI, confidence interval; CRP, c-reactive protein; csDMARD, conventional synthetic disease modifying anti rheumatic drug; DAS28-CRP, disease activity score using c-reactive protein; ESR, erythrocyte sedimentation rate; GM-CSF, granulocyte-macrophage colony-stimulating factor; IL, interleukin; JAK, Janus kinase; MTX, methotrexate; n, number of study arms; RA, rheumatoid arthritis; RCT, randomised controlled trial; Reference, a category to which the coefficient estimates for the categorical predictors are compared to; SE, standard error; TNF-alpha, tumour necrosis factor alpha; VAS, visual analogue scale. Statistically significant moderators in bold.*

**Table 2. Multivariate meta-regression results for the total number of infected patients; prospective cohort studies**

| Moderators | n | $\boldsymbol{\beta}\mathbf{(}\boldsymbol{SE}\mathbf{)}$ | 95%CI | Cochran's Q test (P) | Test for moderator (P) | $\boldsymbol{\sigma}_{\mathbf{1}}^{\mathbf{2}}$ | $\boldsymbol{\sigma}_{\mathbf{2}}^{\mathbf{2}}$ | $\mathbf{I}^{\mathbf{2}}$ |
| --- | --- | --- | --- | --- | --- | --- | --- | --- |
| Biological class (reference: TNF-alpha inhibitor) | 15 |  |  | <0.001 | 0.07 | 0.92 | 0 | 98.92 |
| Constant |  | -1.43(0.29) | [-2.00,-0.85] |  |  |  |  |  |
| Interleukin antagonist |  | -0.26(0.14) | [-0.53, 0.02] |  |  |  |  |  |
| Biological target (reference: TNF-alpha inhibitor) | 7 |  |  | 0.16 | 0.07 | 0.92 | 0.0 | 98.92 |
| Constant |  | 1.43 (0.29 ) | [-0.26, 0.14] |  |  |  |  |  |
| IL-6 inhibitor |  | -0.26 (0.14) | [-0.53, 0.02] |  |  |  |  |  |
| Followup duration | 15 | 0.001(0.001) | [-0.001, 0.002] | <0.001 | 0.27 | 0.85 | 0.03 | 98.87 |
| Age (mean) | 17 | -0.06(0.06) | [-0.19,0.06] | <0.001 | 0.32 | 0.53 | 0.02 | 98.49 |
| Corticosteroid use (percent) | 16 | -0.016(0.01) | [-0.03,0.001] | <0.001 | 0.07 | 0.98 | 0 | 99.0 |
| RA duration (mean) (years) | 15 | -0.031(0.07) | [-0.17,0.11] | <0.001 | 0.67 | 0.58 | 0.02 | 98.12 |
| Previous bDMARD use (percent) | 14 | -0.002(0.01) | [-0.02,-0.01] | <0.001 | 0.72 | 0.54 | 0.01 | 98.71 |
| Disease activity composite variable | 15 | -7.73(2.33) | [-12.30,-3.16] | <0.001 | **0.001** | 0.33 | 0.02 | 97.82 |
| Exclusion criteria composite variable | 19 | -0.15(0.49) | [-1.11, 0.81] | <0.001 | 0.76 | 0.87 | 0.01 | 98.96 |

*bDMARD, biological synthetic disease modifying anti rheumatic drug; CI, confidence interval; RA, rheumatoid arthritis; Reference, a category to which the coefficient estimates for the categorical predictors are compared to; SE, standard error; TNF-alpha, tumour necrosis factor alpha. Statistically significant moderators in bold.*

**Table 3. Multivariate meta-regression results for the total number of infected patients; RCT+OLE studies**

| Moderators | n | $\boldsymbol{\beta}\mathbf{(}\boldsymbol{SE}\mathbf{)}$ | 95%CI | Cochran's Q test(P) | Test for moderator(P) | $\boldsymbol{\sigma}_{\mathbf{1}}^{\mathbf{2}}$ | $\boldsymbol{\sigma}_{\mathbf{2}}^{\mathbf{2}}$ | $\mathbf{I}^{\mathbf{2}}$ |
| --- | --- | --- | --- | --- | --- | --- | --- | --- |
| Biological class(reference: TNF-alpha inhibitor) |  |  |  | <0.0001 | 0.32 | 0.66 | 0.01 | 94.15 |
| Constant |  | -0.29(0.48) | [-1.24, 0.66] |  |  |  |  |  |
| Interleukin antagonist |  | -0.75(0.76) | [-2.24, 0.74] |  |  |  |  |  |
| Biological target (reference: TNF-alpha inhibitor) |  |  |  | <0.0001 | 0.32 | 0.81 | 0.11 | 94.15 |
| Constant |  | -0.29(0.48) | [-1.24, 0.66] |  |  |  |  |  |
| IL-17 inhibitor |  | -0.75(0.76) | [-2.24, 0.74] |  |  |  |  |  |
| Follow up duration |  | 0.002(0.0004) | [0.001, 0.003] | 0.0275 | **<0.001** | 0.08 | 0.01 | 67.67 |
| Age (mean) | 12 | -0.003(0.10) | [-0,19, 0.18] | <0.0001 | 0.97 | 0.69 | 0.02 | 94.41 |
| HAQ-DI (mean) | 12 | 1.94(1.69) | [-1.37, 5.24] | <0.001 | 0.25 | 0.54 | 0.03 | 93.19 |
| Pain VAS (mean) | 11 | 0.001(0.07) | [-0.13, 0.14] | <0.001 | 0.99 | 0.827 | 0.02 | 95.47 |
| Exclusion serious/recurrent infection | 12 | -0.75(0.76) | [-2.24, 0.74] | <0.001 | 0.32 | 0.66 | 0.012 | 94.15 |
| Disease activity composite variable | 12 | -6.17(14.55) | [-34.70, 22.35] | <0.001 | 0.67 | 0.67 | 0.021 | 94.28 |
| Exclusion criteria composite variable | 12 | -0.14(0.17) | [-0.48, 0.19] | <0.001 | 0.40 | 0.71 | 0.01 | 94.53 |

*Anti-CCP, anti-citrullinated protein antibodies; HAQ-DI, health assessment questionnaire-disability index; IL, interleukin; n, number of study arms; RA, rheumatoid arthritis; RCT, randomised controlled trial; RCT+OLE, randomised controlled trial and open label extension; Reference, a category to which the coefficient estimates for the categorical predictors are compared to; SE, standard error; TNF-alpha, tumour necrosis factor alpha; VAS, visual analogue scale. Statistically significant moderators in bold.*

**Table 4. Multivariate meta-regression results for seriously infected patients; RCT studies**

| Moderators | n | $\boldsymbol{\beta}\mathbf{(}\boldsymbol{SE}\mathbf{)}$ | 95%CI | Cochran's Q test (P) | Test for moderator (P) | $\boldsymbol{\sigma}_{\mathbf{1}}^{\mathbf{2}}$ | $\boldsymbol{\sigma}_{\mathbf{2}}^{\mathbf{2}}$ | $\mathbf{I}^{\mathbf{2}}$ |
| --- | --- | --- | --- | --- | --- | --- | --- | --- |
| Biological class(reference: TNF- alpha inhibitor) | 250 |  |  | 0.001 | 0.08 | 0.13 | 0.03 | 39.97 |
| Constant |  | -3.65(0.09) | [-3.82, -3.47] |  |  |  |  |  |
| B-cell inhibitor |  | 0.22(0.19) | [-0.15, 0.58] |  |  |  |  |  |
| Interleukin antagonist |  | 0.15(0.15) | [-0.14, 0.43] |  |  |  |  |  |
| JAK-inhibitor |  | -0.21(0.14) | [ -0.48, 0.07] |  |  |  |  |  |
| Other |  | -0.55(0.44) | [-1.42, 0.32] |  |  |  |  |  |
| T-cell inhibitor |  | -0.2(0.19) | [-0.61, 0.15] |  |  |  |  |  |
| Biological target (reference: TNF-alpha inhibitor) | 245 |  |  | <0.001 | 0.08 | 0.14 | 0.03 | 39.57 |
| Constant |  | -3.66(0.09) | [-3.84, 3.48] |  |  |  |  |  |
| B-cell inhibitor |  | 0.24(0.24) | [-0.23, 0.70] |  |  |  |  |  |
| Rituximab |  | 0.19(0.28) | [-0.35, 0.74] |  |  |  |  |  |
| BTK-inhibitor |  | -0.51(0.64) | [-1.77, 0.75] |  |  |  |  |  |
| GM-CSF-inhibitor |  | -0.56(0.62) | [-1.77, 0.65] |  |  |  |  |  |
| IL-1 inhibitor |  | -0.03(0.37) | [-0.76, 0.70] |  |  |  |  |  |
| IL-17 inhibitor |  | -0.23(0.46) | [-1.14, 0.67] |  |  |  |  |  |
| IL-6 inhibitor |  | 0.21(0.16) | [-0.10, 0.52] |  |  |  |  |  |
| JAK1/2 inhibitor |  | -0.09(0.24) | [ -0.55, 0.38] |  |  |  |  |  |
| JAK1/3 inhibitor |  | -0.15(0.21) | [-0.57, 0.27] |  |  |  |  |  |
| JAK1 inhibitor |  | -0.31(0.25) | [-0.82, 0.19] |  |  |  |  |  |
| Pan-JAK inhibitor |  | -0.36(0.34) | [-1.03, 0.30] |  |  |  |  |  |
| T-cell inhibitor |  | -0.23(0.20) | [-0.62, 0.16] |  |  |  |  |  |
| Followup duration (weeks) | 253 | 0.005(0.002) | [0.001, 0.01] | <0.001 | **0.02** | 0.13 | 0.03 | 39.47 |
| Age (mean) | 233 | 0.01(0.02) | [-0.03, 0.05] | <0.001 | 0.51 | 0.18 | 0.004 | 38.68 |
| MTX dose (mg/week) (mean) | 115 | -0.031(0.03) | [-0.08, 0.02] | 0.0001 | 0.25 | 0.15 | 0.05 | 44.40 |
| DAS28-CRP (mean) | 147 | -0.04(0.11) | [-0.25, 0.18] | 0.95 | 0.74 | 0.06 | 0 | 13.9 |
| CDAI (mean) | 69 | -0.03(0.02) | [-0.07, 0.02] | 0.67 | 0.21 | 0.11 | 0 | 20.15 |
| HAQ-DI (mean) | 208 | -0.011(0.011) | [-0.03, 0.01] | 0.002 | 0.31 | 0.15 | 0.003 | 35.88 |
| TJC-28 (mean) | 50 | 0.02(0.05) | [-0.08, 0.11] | 0.88 | 0.72 | 0.02 | 0.02 | 7.61 |
| Pain VAS (mean) | 151 | 0.003(0.01) | [-0.02, 0.03] | 0.04 | 0.82 | 0.16 | 0 | 34.17 |
| Exclusion serious/recurrent infection | 253 | 0.05(0.11) | [-0.26, 0.17] | <0.001 | 0.68 | 0.15 | 0.02 | 38.96 |
| ESR (mm/h) (mean) | 121 | 0.04(0.011) | [0.02, 0.06] | <0.001 | **0.001** | 0.16 | 0.003 | 39.23 |
| Corticosteroids (percent) | 148 | 0.01(0.005) | [-0.001, 0.02] | 0.001 | 0.07 | 0.13 | 0.03 | 41.42 |
| Corticosteroids dose (mg/day) (mean) | 44 | 0.15(0.14) | [-0.14, 0.43] | <0.001 | 0.31 | 0.41 | 0.01 | 66.01 |
| RA duration (years) (mean) | 213 | -0.01(0.02) | [-0.04, 0.03] | <0.001 | 0.68 | 0.17 | 0.01 | 39.12 |
| 3 or more prior DMARDs (percent) | 16 | -0.01(0.01) | [-0.03, 0.02] | 0.63 | 0.64 | 0.13 | 0 | 37.15 |
| Number of prior DMARDs (mean) | 38 | 0.11(0.14) | [-0.17, 0.38] | 0.005 | 0.45 | 0.06 | 0.09 | 42.84 |
| Number of prior DMARDs (SD) | 26 | 0.28(0.32) | [-0.36, 0.91] | 0.03 | 0.39 | 0.09 | 0.03 | 40.44 |
| Leflunomide use (percent) | 15 | 0.04(0.05) | [-0.06, 0.14] | 0.58 | 0.44 | 0 | 0 | 0 |
| Anti-CCP positive (percent) | 128 | 0.01(0.01) | [0.002, 0.02] | 0.72 | **0.02** | 0.07 | 0.01 | 17.53 |
| Previous bDMARD use (percent) | 162 | 0.0001(0.002) | [-0.003, 0.003] | 0.01 | 0.94 | 0.14 | 0.001 | 33.19 |
| 1 previous bDMARD (percent) | 29 | -0.01(0.01) | [-0.02, 0.004] | 0.92 | 0.22 | 0 | 0 | 0 |
| 2 previous bDMARD (percent) | 29 | 0.01(0.01) | [-0.02, 0.004] | 0.87 | 0.66 | 0.03 | 0 | 7.48 |
| 3 or more previous bDMARD (percent) | 22 | -0.01(0.01) | [-0.03, 0.02] | 0.83 | 0.67 | 0 | 0 | 0 |
| Disease activity composite variable | 233 | -0.22(0.22) | [-0.65, 0.21] | 0.0001 | 0.31 | 0.18 | 0 | 39.29 |
| Exclusion criteria composite variable | 253 | 0.01(0.03) | [-0.05, 0.06] | <0.001 | 0.83 | 0.15 | 0.02 | 38.54 |

*Anti-CCP, anti-citrullinated protein antibodies; bDMARD, biological synthetic disease modifying anti rheumatic drug; BTK, Bruton tyrosine kinase; CDAI = clinical disease activity index; CI, confidence interval; CRP, c-reactive protein; csDMARD, conventional synthetic disease modifying anti rheumatic drug; DAS28-CRP, disease activity score using c-reactive protein; ESR, erythrocyte sedimentation rate; GM-CSF, granulocyte-macrophage colony-stimulating factor; IL, interleukin; JAK, Janus kinase; MTX, methotrexate; n, number of study arms; RA, rheumatoid arthritis; RCT, randomised controlled trial; Reference, a category to which the coefficient estimates for the categorical predictors are compared to; SE, standard error; TNF-alpha, tumour necrosis factor alpha; VAS, visual analogue scale. Statistically significant moderators in bold.*

**Table 5. Multivariate meta-regression results for seriously infected patients; prospective cohort studies**

| Moderators | n | $\boldsymbol{\beta}\mathbf{(}\boldsymbol{SE}\mathbf{)}$ | 95%CI | Cochran's Q test (P) | Test for moderator (P) | $\boldsymbol{\sigma}_{\mathbf{1}}^{\mathbf{2}}$ | $\boldsymbol{\sigma}_{\mathbf{2}}^{\mathbf{2}}$ | $\mathbf{I}^{\mathbf{2}}$ |
| --- | --- | --- | --- | --- | --- | --- | --- | --- |
| Biological class (reference: TNF-alpha inhibitor) | 15 |  |  | <0.0001 | 0.36 | 0.26 | 0.01 | 88.94 |
| Intercept |  | -2.97(0.21) | [-3.38, -2.55] |  |  |  |  |  |
| Interleukin antagonist |  | -0.26(0.28) | [-0.81, 0.29] |  |  |  |  |  |
| Biological target (reference: TNF-alpha inhibitor) | 15 |  |  | <0.0001 | 0.36 | 0.26 | 0.01 | 88.94 |
| Intercept |  | -2.97(0.21) | [-3.38, -2.55] |  |  |  |  |  |
| IL-6 inhibitor |  | -0.26(0.28) | [-0.81, 0.29] |  |  |  |  |  |
| Follow-up duration (weeks) | 15 | 0.0003(0.0004) | [-0.0004, 0.001] | < 0.0001 | 0.39 | 0.24 | 0.02 | 88.42 |
| Age (mean) | 16 | 0.04(0.05) | [-0.06, 0.15] | < 0.0001 | 0.43 | 0.27 | 0.03 | 89.66 |
| Exclusion history of serious infection | 18 | 1.05(0.85) | [-0.62, 2.71] | < 0.0001 | 0.22 | 0.20 | 0.05 | 86.37 |
| ESR (mm/h) (mean) | 10 | 0.025(0.01) | [0.01, 0.05] | 0.0871 | **0.014** | 0 | 0.06 | 50.02 |
| Corticosteroid use (percent) | 12 | -0.01(0.02) | [-0.04, 0.03] | < 0.0001 | 0.69 | 0.25 | 0.06 | 91.29 |
| RA duration (years) (mean) | 15 | 0.12(0.06) | [-0.004, 0.25] | < 0.0001 | 0.06 | 0.29 | 0 | 87.75 |
| Previous bDMARD use (percent) | 12 | 0.002(0.005) | [-0.01, 0.012] | < 0.0001 | 0.77 | 0.11 | 0.16 | 88.15 |
| Disease activity composite variable | 15 | 2.63(3.24) | [-3.73, 8.98] | < 0.0001 | 0.42 | 0.28 | 0.05 | 90.45 |
| Exclusion criteria composite variable | 18 | 0.14(0.13) | [-0.11, 0.40] | < 0.001 | 0.28 | 0.21 | 0.05 | 86.87 |

*Anti-CCP, anti-citrullinated protein antibodies; bDMARD, biological synthetic disease modifying anti rheumatic drug; IL, interleukin; JAK, Janus kinase; MTX, methotrexate; n, number of study arms; RA, rheumatoid arthritis; RCT, randomised controlled trial; Reference, a category to which the coefficient estimates for the categorical predictors are compared to; SE, standard error; TNF-alpha, tumour necrosis factor alpha. Statistically significant moderators in bold.*

**Table 6. Multivariate meta-regression results for seriously infected patients; registry studies**

| Moderators | n | $\boldsymbol{\beta}\mathbf{(}\boldsymbol{SE}\mathbf{)}$ | 95%CI | Cochran's Q test (P) | Test for moderator (P) | $\boldsymbol{\sigma}_{\mathbf{1}}^{\mathbf{2}}$ | $\boldsymbol{\sigma}_{\mathbf{2}}^{\mathbf{2}}$ | $\mathbf{I}^{\mathbf{2}}$ |
| --- | --- | --- | --- | --- | --- | --- | --- | --- |
| Biological class(reference: TNF-alpha inhibitor) | 34 |  |  | < 0.0001 | 0.24 | 0.12 | 0.27 | 98.51 |
| Constant |  | -2.63(0.17) | [-2.95, -2.30] |  |  |  |  |  |
| B-cell inhibitor |  | 0.31(0.28) | [-0.23, 0.86] |  |  |  |  |  |
| T-cell inhibitor |  | -0.22(0.27) | [-0.74, 0.30] |  |  |  |  |  |
| Biological class (reference: TNF-alpha inhibitor) | 34 |  |  | < 0.001 | 0.24 | 0.12 | 0.27 | 98.51 |
| Constant |  | -2.63(0.17) | [-2.95, -2.30] |  |  |  |  |  |
| B-cell inhibitor |  | 0.31(0.28) | [-0.23, 0.86] |  |  |  |  |  |
| T-cell inhibitor |  | -0.22(0.27) | [-0.74, 0.30] |  |  |  |  |  |
| Follow-up duration (weeks) | 34 | -0.0001(0.0004) | [-0.001, 0.001] | <0.001 | 0.83 | 0.16 | 0.27 | 98.64 |
| Age (mean) | 31 | 0.02(0.05) | [-0.07, 0.11] | <0.001 | 0.63 | 0.18 | 0.29 | 98.74 |
| COPD (percent) | 20 | 0.003(0.03) | [-0.06, 0.06] | <0.001 | 0.91 | 0.20 | 0.35 | 99.12 |
| HAQ-DI (mean) | 11 | 0.76(0.43) | [-0.08, 1.61] | <0.001 | 0.08 | 0.30 | 0.01 | 98.31 |
| Renal disease (patients) | 18 | -0.0004(0.001) | [-0.003, 0.002] | <0.001 | 0.70 | 0.10 | 0.40 | 98.91 |
| Renal disease (percent) | 18 | 0.21(0.13) | [-0.03, 0.47] | <0.001 | 0.09 | 0 | 0.40 | 98.67 |
| Previous severe infection (patients) | 19 | 0.0004(0.001) | [-0.001, 0.002] | <0.001 | 0.51 | 0.04 | 0.40 | 98.84 |
| Previous severe infection (percent) | 19 | 0.047(0.04) | [-0.03, 0.13] | <0.001 | 0.25 | 0.08 | 0.35 | 98.82 |
| Exclusion serious/recurrent infection | 37 | -0.08(0.53) | [-1.11, 0.95] | <0.001 | 0.88 | 0.15 | 0.26 | 98.51 |
| Corticosteroid use (percent) | 35 | 0.005(0.01) | [-0.01, 0.02] | <0.001 | 0.54 | 0.16 | 0.27 | 98.57 |
| Diabetes mellitus (patients) | 32 | -0.0003(0.0002) | [-0.001, 0.0001] | <0.001 | 0.13 | 0.10 | 0.28 | 98.54 |
| Diabetes mellitus (percent) | 32 | -0.02(0.02) | [-0.06, 0.02] | <0.001 | 0.29 | 0.12 | 0.28 | 98.61 |
| CHF (percent) | 15 | 0.02(0.03) | [-0.04, 0.08] | <0.001 | 0.45 | 0.13 | 0.44 | 99.08 |
| Smoking. Ever (percent) | 12 | 0.001(0.002) | [-0.003, 0.005] | <0.001 | 0.56 | 0.25 | 0.02 | 98.24 |
| Smoking. current (percent) | 10 | 0(0.03) | [-0.06, 0.06] | <0.001 | 1 | 0.38 | 0.02 | 98.72 |
| Previous bDMARD use (percent) | 18 | 0.01(0.01) | [-0.004, 0.02] | <0.001 | 0.21 | 0.05 | 0.58 | 99.17 |
| Disease activity composite variable | 18 | 2.55(1.38) | [-0.15, 5.24] | <0.001 | 0.06 | 0.18 | 0.01 | 96.40 |
| Exclusion criteria composite variable | 37 | -0.09(0.17) | [-0.42, 0.23] | <0.001 | 0.58 | 0.14 | 0.26 | 98.48 |

*Anti-CCP, anti-citrullinated protein antibodies; bDMARD, biological synthetic disease modifying anti rheumatic drug; CHF, congestive heart failure; CI, confidence interval; COPD, chronic obstructive pulmonary disease; HAQ-DI, health assessment questionnaire-disability index; IL, interleukin; n, number of study arms; RA, rheumatoid arthritis; Reference, a category to which the coefficient estimates for the categorical predictors are compared to; SE, standard error; TNF-alpha, tumour necrosis factor alpha; VAS, visual analogue scale. Statistically significant moderators in bold*

**Appendix 11: Creation of composite disease severity score**

The following individual disease severity scores, when available, were used to create a composite variable:

Disease Activity Score using CRP or ESR (DAS28-CRP/ESR)

Clinical Disease Activity Index for RA (CDAI)

Health Assessment Questionnaire – Disability Index (HAQ-DI)

Swollen Joint Count in 66 joints (SJC66)

Tender Joint Count in 68 Joints (TJC68)

Patient’s Global Assessment (Visual Analogue Scale 0-100)

Physicians Global Assessment (Visual Analogue Scale 0-100)

Pain score (Visual Analogue Scale 0-100)

Functional Assessment of Chronic Illness Therapy – Fatigue (FACIT-fatigue)

The composite variable ‘disease severity score’ was created by scaling each separate disease activity index and taking the mean of all these scaled values together.

**Appendix 12: Creation of exclusion criteria composite variable**

Studies could exclude patients from participating based on certain criteria. Exclusion criteria often entailed a history of serious infection or significant comorbidities, which influence infection risk. We therefore found it appropriate to include information on exclusion criteria in the analyses. To that end, we extracted exclusion criteria per study arm, and found studies use a combination of different exclusion criteria. Since the combination of exclusion criteria was not the same across studies, we decided to create a composite variable of exclusion criteria. This composite variable is the sum of the number of exclusion criteria that was used in the study.

**Table 1. Exclusion criteria and frequency of use across studies**

| **Exclusion criterion** | **Frequency (study arms)** | **Percentage (study arms)** |
| --- | --- | --- |
| Significant comorbidity | 118 | 23,0 |
| History of malignancy | 111 | 21,7 |
| Previous serious infection or recurrent infections | 188 | 36,7 |
| Other inflammatory disease | 128 | 25,0 |
| TB | 149 | 29,1 |
| Chronic viral infection | 61 | 11,9 |
| Leukopenia | 77 | 15,0 |
| Lab abnormalities | 88 | 17,2 |
| High dose corticosteroids | 75 | 14,6 |
| Prior bDMARD failure | 42 | 8,2 |
| Recent or future surgery | 8 | 1,6 |
| Organ transplantation | 6 | 1,2 |
| Disability | 4 | 0,8 |
| Immunodeficiency | 4 | 0,8 |

*bDMARD, biological disease modifying anti rheumatic drug; TB, tuberculosis. Studies could use multiple exclusion criteria mentioned in this table.*

**Appendix 13: characteristics of included registries and trials**

Characteristics of studies expressed as the number of included studies and patients they contain per study design and geographical location. We also registered information on the mode of infectious adverse event reporting and follow up duration. The last table describes demographic characteristics of patients.

**Table 1. Characteristics of included studies**

|  | **N (studies)** | **%** | **patients** | **%*** |
| --- | --- | --- | --- | --- |
| **total** | **242** | **100.0** | **293431** | **100.0** |
| RCT | 150 | 62.0 | 52352 | 17.8 |
| Registry | 34 | 14.0 | 172892 | 58.9 |
| Prospective cohort | 26 | 10.7 | 42399 | 14.4 |
| Retrospective cohort | 7 | 2.9 | 4798 | 1.6 |
| RCT+OLE | 13 | 5.4 | 16569 | 5.6 |
| Open label studies | 12 | 5.0 | 4421 | 1.5 |
| North America | 122 | 50.4 | 149308 | 50.9 |
| Latin America | 58 | 24.0 | 27457 | 9.4 |
| Europe | 152 | 62.8 | 124421 | 42.4 |
| East-Asia | 82 | 33.9 | 70562 | 24.0 |
| South-Asia | 16 | 6.6 | 5363 | 1.8 |
| Africa | 8 | 3.3 | 2676 | 0.9 |
| Middle East | 8 | 3.3 | 2050 | 0.7 |
| Oceania | 16 | 6.6 | 15540 | 5.3 |
| **Reporting** |  |  |  |  |
| Total number of infected patients | 129 | 53.3 | 58789 | 20.0 |
| Total number of seriously infected patients | 157 | 64.9 | 168042 | 57.3 |
| Total number of infectious events | 25 | 10.3 | 13937 | 4.7 |
| Total number of serious infectious events | 38 | 15.7 | 90204 | 30.7 |
| Incidence rate total number of infected pateitns | 18 | 7.4 | 22647 | 7.7 |
| Incidence rate total number of seriously infected patients | 48 | 19.8 | 155217 | 52.9 |
| Event rate total number of infections | 6 | 2.5 | 2753 | 0.9 |
| Event rate total number of serious infections | 13 | 5.4 | 26344 | 9.0 |
| individual infections | 166 | 68.6 | 97082 | 33.1 |
| individual infectious events | 29 | 12.0 | 90443 | 30.8 |
|  | median | IQR |  |  |
| Follow-up duration (weeks) | 24 | 16-48 |  |  |
| Follow-up duration (Patient years) | 747 | 303-2534 |  |  |

*RCT, randomised controlled trial; RCT+OLE, randomised controlled trial and open label extension*

**Table 2. Number of included studies, study arms and patients across different biological classes**

|  | TNF alpha | % of total | Interleukin antagonists | % of total | IL-6 | % of Interleukin antagonists | B-cell inhibitors | % of total | RTX | % of B-cell inhibitor | T-cell inhibitors | % of total | JAK-inhibitors | % of total | Tofacitinib | % of JAK-inhibitor |
| --- | --- | --- | --- | --- | --- | --- | --- | --- | --- | --- | --- | --- | --- | --- | --- | --- |
| studies | 117 | 48,3 | 57 | 23,6 | 46 | 80,7 | 34 | 14,0 | 23 | 67,6 | 26 | 10,7 | 29 | 12,0 | 10 | 34,5 |
| study arms | 205 | 40,0 | 115 | 22,5 | 84 | 73,0 | 59 | 11,5 | 26 | 44,1 | 35 | 6,8 | 70 | 13,7 | 24 | 34,3 |
| patients | 170826 | 58,2 | 49182 | 16,8 | 45379 | 92,3 | 22685 | 7,7 | 19149 | 84,4 | 40098 | 13,7 | 9243 | 3,1 | 3845 | 41,6 |

*IL, interleukin; JAK, Janus kinase; TNF, tumour necrosis factor*

**Table 3. Characteristics of included patients**

|  | N | %* |
| --- | --- | --- |
| **Total** | **293431** | **100** |
| Female | 231893 | 79.0 |
| Male | 59597 | 20.3 |
| RF positive | 58953 | 20.1 |
| Anti-CCP positive | 22911 | 7.8 |
| MTX | 146859 | 50.0 |
| Corticosteroid use | 151837 | 51.7 |
| Previous biological use | 77385 | 26.4 |
|  | N | %** |
| total | 43790 | 100 |
| Caucasian | 39064 | 89.2 |
| Black | 1107 | 2.5 |
| Hispanic | 506 | 1.2 |
| Asian | 1468 | 3.4 |
| Native American | 191 | 0.4 |
| Other | 1454 | 3.3 |
|  | Weighted mean |  |
| Age | 56.6 |  |
| Weight | 60.2 |  |
| RA duration | 9.31 |  |
| RF titre | 276 |  |
| Anti-CCP titre | 464 |  |
| CRP | 26.2 |  |
| ESR | 45.1 |  |

*Anti-CCP, anti-citrullinated protein antibodies; CRP, c-reactive protein; ESR, erythrocyte sedimentation rate; MTX, methotrexate; n, number of patients; RA, rheumatoid arthritis; RF, rheumatoid factor.*

**percentage of the total number of patients, since not all studies reported on all characteristics*

***percentage of the number of patients for whom race was reported*

**Appendix 14: geographical location of trials and case reports**

We extracted information on the geographical location where included studies were conducted. Case reports and registry studies usually originated in a single country, RCTs and open label (extension) studies, on the other hand, were performed in multiple countries across different continents. For case reports, we included individual countries. For the other studies, we included the continents where the study took place. One study could therefore account for multiple continents on the map.

**Figure 1A. Geographical location of case reports**

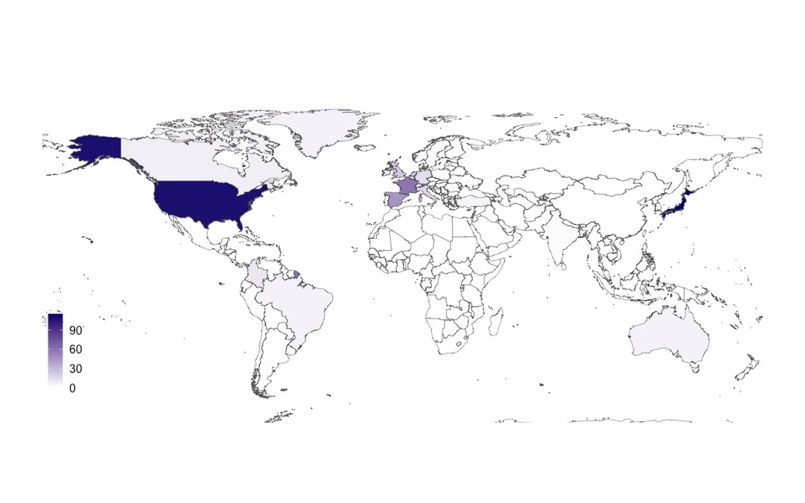

**Figure 1B. Geographical location of studies**

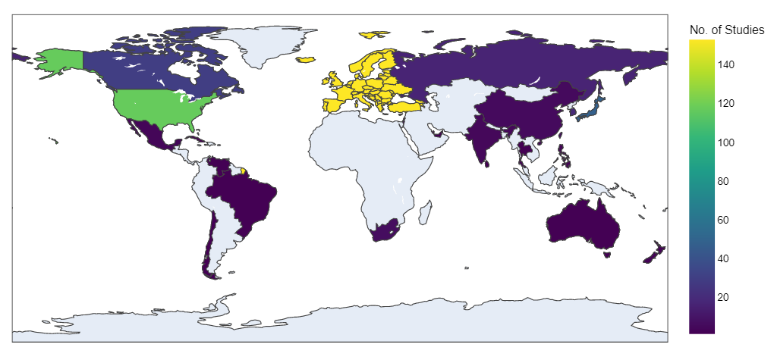

**Appendix 15: Multivariate meta-regression for the total number of (seriously) infected patients in RCT studies with a follow-up duration of 18 weeks or less and between 19-38 weeks**

**Table 1. Multivariate meta-regression of RCT studies with a follow-up duration of 18 weeks or less and between 19-38 weeks**

| Total number of infected patients: follow-up duration of  18 weeks or less | | | | | | | Total number of infected patients: follow-up duration of 19-38 weeks | | | | | |
| --- | --- | --- | --- | --- | --- | --- | --- | --- | --- | --- | --- | --- |
| Moderators | n | $\beta$ | 95%CI | Test for moderator(P) | Cochran's Q test(P) | $I^{2}$ | n | $\beta$ | 95%CI | Test for moderator(P) | Cochran's test(P) |  |
| Biological target | 75 |  |  | 0.10 | <0.001 | 67.58 | 92 |  |  | **<0.001** | 0.51 | 84.69 |
| (reference: TNF-alpha inhibitor) |  |  |  |  |  |  |  |  |  |  |  |  |
| Constant |  | -0.86 | [-1.09, -0.63] |  |  |  |  | -0.71 | [ -0.87, -0.55] |  |  |  |
| Anti-LT-alpha |  | -0.27 | [-0.88, 0.33] |  |  |  |  | - | - |  |  |  |
| B-cell inhibitor |  | 0.22 | [-0.43, 0.87] |  |  |  |  | -0.18 | [ -0.52, 0.15] |  |  |  |
| Rituximab |  |  |  |  |  |  |  | -0.18 | [-0.65, 0.30] |  |  |  |
| BTK-inhibitor |  | -0.91 | [-1.43, -0.38] |  |  |  |  | - | - |  |  |  |
| IL-17 inhibitor |  | -0.19 | [-0.98, 0.61] |  |  |  |  | -0.46 | [-1.26, 0.35] |  |  |  |
| IL-20 inhibitor |  | -0.39 | [-1.42, 0.64] |  |  |  |  | - | - |  |  |  |
| IL-6 inhibitor |  | -0.02 | [-0.57, 0.52] |  |  |  |  | 0.04 | [-0.21, 0.29] |  |  |  |
| IL-1 inhibitor |  |  |  |  |  |  |  | -0.002 | [-0.69, 0.68] |  |  |  |
| JAK1/2 inhibitor |  | -0.33 | [-1.23, 0.56] |  |  |  |  | 0.30 | [-0.17, 0.77] |  |  |  |
| JAK1/3 inhibitor |  | -0.19 | [-0.71, 0.33] |  |  |  |  | - | - |  |  |  |
| JAK1 inhibitor |  | 0.01 | [-0.44, 0.46] |  |  |  |  | 0.21 | [-0.10, 0.52] |  |  |  |
| Pan-JAK inhibitor |  | -0.19 | [-1.01, 0.62] |  |  |  |  | - | - |  |  |  |
| T-cell inhibitor |  | - | - |  |  |  |  | 0.12 | [-0.33, 0.58] |  |  |  |
| Age (mean) | 68 | 0.04 | [-0.01, 0.09] | 0.11 | <0.001 | 56.17 | 84 | 0.01 | [-0.02, 0.05] | 0.45 | <0.001 | 78.32 |
| MTX dose (mg/week, mean) | 41 | -0.03 | [-0.08, 0.02] | 0.23 | 0.002 | 54.22 | 41 | 0.03 | [-0.003, 0.06] | 0.08 | <0.001 | 83.36 |
| DAS28-CRP (mean) | 48 | -0.20 | [-0.58, 0.19] | 0.31 | <0.001 | 52.87 | 65 | -0.01 | [-0.15, 0.13] | 0.88 | <0.001 | 75.50 |
| CDAI (mean) | 22 | -0.04 | [-0.09, 0.01] | 0.09 | 0.24 | 43.78 | 22 | -0.01 | [-0.03, 0.02] | 0.68 | <0.001 | 77.54 |
| HAQ-DI (mean) | 55 | -0.21 | [-0.61, 0.19] | 0.31 | <0.001 | 61.65 | 83 | -0.003 | [-0.02 ,0.01] | 0.60 | <0.001 | 80.95 |
| TJC-28 (mean) | 19 | 0.01 | [-0.07, 0.09] | 0.79 | 0.42 | 29.14 | 17 | -0.03 | [-0.10, 0.03] | 0.32 | <0.001 | 76.06 |
| Pain (mean VAS) | 36 | 0.01 | [-0.02, 0.03] | 0.60 | 0.03 | 47.20 | 50 | -0.004 | [-0.03, 0.022] | 0.76 | <0.001 | 84.04 |
| Exclusion serious infection | 77 | 0.09 | [-0.21, 0.39] | 0.55 | <0.001 | 65.60 | 89 | 0.09 | [-0.124, 0.30] | 0.41 | <0.001 | 85.01 |
| ESR (mm/h, mean) | 24 | -0.02 | [-0.06, 0.02] | 0.41 | <0.001 | 67.25 | 37 | -0.006 | [-0.03, 0.02] | 0.61 | <0.001 | 82.53 |
| Corticosteroids (percent) | 48 | 0.01 | [0.002, 0.02] | **0.01** | 0.15 | 3.71 | 62 | -0.005 | [-0.01, 0.00] | 0.20 | <0.001 | 88.43 |
| Corticosteroid dose (mg/day, mean) | 24 | -0.05 | [-0.17, 0.06] | 0.35 | 0.61 | 25.07 | 10 | 0.10 | [-0.15, 0.35] | 0.42 | 0.002 | 72.46 |
| RA duration (years, mean) | 54 | 0.02 | [-0.04, 0.08] | 0.46 | 0.01 | 44.07 | 81 | 0.04 | [0.01, 0.07] | **0.01** | <0.001 | 79.89 |
| Number of prior csDMARDs (mean) | - | - | - | - | - | - | 12 | 0.81 | [0.40, 1.23] | **<0.001** | <0.001 | 83.81 |
| Number of prior csDMARDS (SD) | - | - | - | - | - | - | 12 | 0.68 | [-0.54, 1.9] | 0.27 | <0.001 | 92.65 |
| Leflunomide (percent) | - | - | - | - | - | - | 10 | 0.09 | [0.003, 0.18] | 0.04 | <0.001 | 87.60 |
| Anti-CCP positive (percent) | 43 | 0.002 | [-0.004, 0.01] | 0.54 | 0.001 | 58.49 | 50 | -0.001 | [-0.01, 0.01] | 0.93 | <0.001 | 67.48 |
| Previous biological use (percent) | 46 | 0.002 | [-0.001, 0.004] | 0.21 | 0.24 | 27.74 | 66 | 0.001 | [-0.001, 0.004] | 0.35 | <0.001 | 79.13 |
| 1 previous biological (percent) | - | - | - | - | - | - | 16 | -0.02 | [-0.03, -0.01] | <0.001 | 0.02 | 48.57 |
| 2 previous biologicals (percent) | - | - | - | - | - | - | 16 | 0.01 | [-0.02, 0.04] | 0.37 | <0.001 | 82.04 |
| Disease activity composite variable (mean) | 67 | -0.63 | [-211, 0.85] | 0.41 | <0.001 | 63.90 | 88 | -0.09 | [-0.33, 0.14] | 0.44 | <0.001 | 81.70 |
| Exclusion criteria composite variable | 77 | 0.06 | [-0.04, 0.15] | 0.24 | <0.001 | 65.25 | 92 | 0.001 | [-0.06, 0.06] | 0.97 | <0.001 | 84.65 |
| Total number of seriously infected patients: follow-up duration of  18 weeks or less | | | | | | | **Total number of seriously infected patients: follow-up duration of 19-38 weeks** | | | | | |
| Moderators | n | $\beta$ | 95%CI | Test for moderator(P) | Cochran's Q $I^{2}$ test(P) | | n | $\beta$ | 95%CI | Test for moderator(P) | Cochran's Q $I^{2}$  test(P) | |
| Biological target (reference: TNF-alpha inhibitor) | 91 |  |  | **0.03** | 0.99 | 0 | 105 |  |  | 0.3127 | 0.01 | 39.74 |
| Constant |  | -3.57 | [-3.86, -3.27] |  |  |  |  | -3.77 | [-4.01, -3.54] |  |  |  |
| BTK-inhibitor |  | -0.7219 | [-1.75, 0.31] |  |  |  |  | - | - |  |  |  |
| GM-CSF inhibitor |  | -0.6557 | [-1.75, 0.44] |  |  |  |  | - | - |  |  |  |
| IL-17 inhibitor |  | -0.02 | [-0.80, 0.77] |  |  |  |  | -0.77 | [-2.38, 0.84] |  |  |  |
| IL-6 inhibitor |  | 0.2532 | [-0.39, 0.90] |  |  |  |  | 0.15 | [-0.24, 0.54] |  |  |  |
| IL-1 inhibitor |  | 0.1793 | [-0.72, 1.08] |  |  |  |  | - | - |  |  |  |
| JAK1/2 inhibitor |  | -0.3263 | [-1.75, 1.10] |  |  |  |  | -0.09 | [-0.73, 0.56] |  |  |  |
| JAK1/3 inhibitor |  | -0.3437 | [-1.01, 0.32] |  |  |  |  | - | - |  |  |  |
| JAK1 inhibitor |  | -0.4253 | [-0.99, 0.14] |  |  |  |  | - | - |  |  |  |
| Pan-JAK inhibitor |  | -0.9455 | [-1.70, -0.19] |  |  |  |  | - | - |  |  |  |
| B-cell inhibitor |  | 0.9679 | [0.14, 1.79] |  |  |  |  | -0.69 | [-1.53, 0.16] |  |  |  |
| T-cell inhibitor |  | - | - |  |  |  |  | -0.23 | [-0.83, 0.37] |  |  |  |
| Rituximab |  | - | - |  |  |  |  | 0.40 | [-0.21, 1.01] |  |  |  |
| Age (mean) | 85 | 0.085 | [-0.003, 0.17] | 0.06 | 0.99 | 13.62 | 94 | -0.0002 | [-0.07, 0.07] | **0.004** | 0.99 | 40.87 |
| MTX dose (mg/week, mean) | 41 | -0.0774 | [-0.14, -0.02] | **0.01** | 0.99 | 0.42 | 54 | 0.041 | [-0,05, 0,13] | **0.001** | 0.36 | 51.83 |
| DAS28-CRP (mean) | 58 | -0.3723 | [-1.13, 0.39] | 0.34 | 0.99 | 3.50 | 64 | 0.08 | [-0.16, 0.33] | 0.515 | 0.50 | 18.09 |
| CDAI (mean) | 30 | -0.0999 | [-0.18, -0.02] | **0.02** | 0.99 | 1.88 | 30 | 0.009 | [-0.04, 0.06] | 0.26 | 0.75 | 28.46 |
| HAQ-DI (mean) | 71 | 0.1243 | [-0.84, 1.09] | 0.80 | 0.98 | 18.55 | 92 | -0.01 | [-0.03, 0.01] | 0.063 | 0.47 | 30.18 |
| TJC-28 (mean) | 24 | -0.0066 | [-0.18, 0.16] | 0.94 | 0.94 | 0 | 25 | 0.03 | [-0.08, 0.14] | 0.903 | 0.61 | 0 |
| Pain (mean VAS) | 60 | -0.0166 | [-0.07, 0.03] | 0.50 | 0.99 | 8.98 | 65 | 0.02 | [-0.02, 0.06] | **<0.001** | 0.30 | 52.28 |
| Exclusion serious infection | 91 | 0.0257 | [-0.46, 0.51] | 0.92 | 0.95 | 16.41 | 105 | 0.25 | [-0.08, 0.57] | **0.002** | 0.13 | 40.56 |
| ESR (mm/h, mean) | 35 | 0.0197 | [-0.07, 0.10] | 0.65 | 0.81 | 23.93 | 57 | 0.04 | [0.01, 0.07] | 0.16 | 0.01 | 24.0 |
| Corticosteroids (percent) | 51 | 0.0291 | [0.01, 0.04] | **<0.001** | 0.99 | 0 | 67 | 0.01 | [-0.01, 0.02] | **0.01** | 0.34 | 37.88 |
| Corticosteroid dose (mg/day, mean) | 22 | -0.1375 | [-0.55, 0.28] | 0.52 | 0.48 | 25.49 | 14 | 0.35 | [-0.06, 0.76] | **0.003** | 0.09 | 73.084 |
| RA duration (years, mean) | 70 | 0.0313 | [-0.08, 0.14] | 0.58 | 0.97 | 19.63 | 92 | 0.03 | [-0.02, 0.09] | **0.01** | 0.21 | 36.69 |
| Anti-CCP positive (percent) | 44 | 0.0086 | [-0.01, 0.03] | 0.33 | 0.93 | 10.52 | 48 | 0.01 | [-0.01, 0.03] | 0.81 | 0.44 | 10.75 |
| Previous bDMARD use (percent) | 55 | 0.0042 | [-0.003, 0.01] | 0.24 | 0.89 | 14.56 | 72 | 0.002 | [-0.001,0.005] | 0.65 | 0.18 | 9.60 |
| 1 previous bDMARD (percent) | 13 | 0.0137 | [-0.01, 0.04] | 0.21 | 0.99 | 0 | 15 | -0.02 | [-0,03,-0.003] | 0.93 | 0.01 | 0 |
| 2 previous bDMARD (percent) | 13 | 0.0333 | [-0.03, 0.10] | 0.33 | 0.98 | 0 | 15 | 0.001 | [-0,034, 0.04] | 0.48 | 0.95 | 13.47 |
| 3 or more previous bDMARD (percent) | 10 | 0.0265 | [-0.03, 0.08] | 0.36 | 0.98 | 0 | 11 | -0.01 | [-0,04, 0.02] | 0.65 | 0.66 | 0 |
| Disease activity composite variable (mean) | 83 | -1.2384 | [-3.99, 1.51] | 0.38 | 0.96 | 17.17 | 102 | -0.12 | [-0.55, 0.30] | **0.01** | 0.57 | 36.89 |
| Exclusion criteria composite variable | 91 | -0.0103 | [-0.16, 0.14] | 0.89 | 0.95 | 15.94 | 108 | 0.01 | [-0.06, 0.08] | **0.001** | 0.78 | 41.69 |
| Number of prior DMARDs (mean)  - |  | - | - | - | - | - | 21 | 0.27 | [0.02, 0.52] | 0.55 | 0.04 | 0 |
| Number of prior DMARDs (SD)  - |  | - | - | - | - | - | 19 | 0.48 | [-0.10, 1.07] | 0.65 | 0.10 | 8.41 |
| Leflunomide use (percent)  - |  | - | - | - | - | - | 10 | 0.07 | [-0.05, 0.19] | 0.51 | 0.29 | 0 |

*Anti-CCP, anti-citrullinated protein antibodies; bDMARD, biological disease modifying anti rheumatic drug; BTK, Burton tyrosine kinase; CDAI, clinical disease activity index; CI, 95% confidence interval; CRP, c-reactive protein; csDMARD, conventional synthetic disease modifying anti rheumatic drug; DAS28-CRP, disease activity score using c-reactive protein; ESR, erythrocyte sedimentation rate; GM-CSF, granulocyte-macrophage colony-stimulating factor; IL, interleukin; JAK, Janus kinase; MTX, methotrexate; n, number of study arms; RA, rheumatoid arthritis; Reference, a category to which the coefficient estimates for the categorical predictors are compared to; TNF-alpha, tumour-necrosis factor alpha; VAS, visual analogue scale.*

*Statistically significant moderators in bold.*

**Appendix 16: Multivariate meta regression of the total number of (seriously) infected patients in RCT’s with a follow-up duration of 38-60 weeks**

**Table 1 presents the results of the multivariate –meta-regression for the total number of infected patients in RCTs with a follow-up duration of 39-60 weeks.**

**Table 1: Multivariate meta-regression results for the total number of infected patients in RCT studies with a follow-up duration of 39-60 weeks**

| Moderators | n | $\boldsymbol{\beta(SE)}$ | 95%CI | Test for moderator (P) | Cochran's Q test (P) | $\boldsymbol{I}^{\boldsymbol{2}}$ |
| --- | --- | --- | --- | --- | --- | --- |
| Biological target (reference: TNF-alpha inhibitor) | 26 |  |  | 0.97 | <0.001 | 96.80 |
| TNF-alpha inhibitor (reference) |  | -0.03(0.27) | [-0.56, 0.51] |  |  |  |
| B- cell inhibitor |  | 0.18(0.62) | [-1.04, 1.39] |  |  |  |
| IL-6 inhibitor |  | 0.12(0.49) | [-0.83, 1.08] |  |  |  |
| T-cell inhibitor |  | -0.04(0.13) | [-0.30, 0.22] |  |  |  |
| Age (mean) | 34 | -0.03(0.03) | [-0.09, 0.03] | 0.26 | <0.001 | 95.33 |
| MTX dose (mg/week, mean) | 19 | -0.01(0.08) | [-0.16, 0.14] | 0.92 | <0.001 | 93.58 |
| Biological class | 36 | 0.07(0.50) | [-0.90, 1.05] | 0.88 | <0.001 | 96.19 |
| Biological target | 36 | -0.59(0.82) | [-2.20, 1.02] | 0.47 | <0.001 | 95.97 |
| Biological name | 36 | -0.06(0.17) | [-0.39, 0.27] | 0.71 | <0.001 | 96.96 |
| DAS28-CRP (mean) | 18 | -0.27(0.40) | [ -1.05, 0.52] | 0.50 | <0.001 | 86.92 |
| HAQ-DI (mean) | 32 | -0.38(0.55) | [-1.46, 0.70] | 0.49 | <0.001 | 95.70 |
| Pain (mean VAS) | 16 | -0.06(0.03) | [-0.12, 0.01] | 0.09 | <0.001 | 91.09 |
| Exclusion serious infection | 36 | 0.11(0.34) | [-0.55, 0.77] | 0.75 | <0.001 | 95.80 |
| ESR (mm/h, mean) | 19 | 0.02(0.02) | [-0.02, 0.06] | 0.35 | <0.001 | 89.80 |
| Corticosteroids (percent) | 25 | 0.001(0.01) | [-0.01, 0.01] | 0.95 | <0.001 | 90.10 |
| RA duration (years, mean) | 32 | -0.03(0.03) | [-0.08, 0.03] | 0.34 | <0.001 | 88.93 |
| Anti-CCP positive (percent) | 28 | -0.002(0.013) | [-0.03, 0.023] | 0.89 | <0.001 | 90.79 |
| Previous biological use (percent) | 22 | -0.003(0.01) | [-0.01, 0.01] | 0.57 | <0.001 | 95.87 |
| Disease activity composite variable (mean) | 34 | -1.32(2.37) | [-5.97, 3.33] | 0.58 | <0.001 | 95.94 |
| Exclusion criteria composite variable | 36 | 0.10(0.13) | [-0.16, 0.3583] | 0.45 | <0.001 | 95.69 |

*Anti-CCP, anti-citrullinated protein antibodies; BTK, Burton tyrosine kinase; CDAI = clinical disease activity index; CI, confidence interval; CRP, c-reactive protein; csDMARD, conventional synthetic disease modifying anti rheumatic drug; DAS28-CRP, disease activity score using c-reactive protein; ESR, erythrocyte sedimentation rate; GM-CSF, granulocyte-macrophage colony-stimulating factor; IL, interleukin; JAK, Janus kinase; MTX, methotrexate; n, number of study arms; RA, rheumatoid arthritis; RCT, randomised controlled trial; Reference, a category to which the coefficient estimates for the categorical predictors are compared to; SE, standard error; TNF-alpha, tumour necrosis factor alpha; VAS, visual analogue scale. Statistically significant moderators in bold.*

**The following table presents the results of seriously infected patients in multivariate-meta regression for RCT 39-60 weeks.**

**Table 2. Multivariate meta-regression results for RCT studies with follow up duration between 39 and 60 weeks (seriously infected patients)**

| Moderators | k | $\boldsymbol{\beta}\mathbf{(}\boldsymbol{SE}\mathbf{)}$ | 95%CI | Test for moderator (P) | Cochran's Q test (P) | $\boldsymbol{\sigma}_{\mathbf{1}}^{\mathbf{2}}$ | $\boldsymbol{\sigma}_{\mathbf{2}}^{\mathbf{2}}$ | $\mathbf{I}^{\mathbf{2}}$ |
| --- | --- | --- | --- | --- | --- | --- | --- | --- |
| Biological target (reference: TNF-alpha inhibitor) | 36 |  |  | 0.35 | 0.003 | 0.09 | 0.07 | 54.75 |
| Contact |  | -3.3748(0.1832) | [-3.73, -3.02] |  |  |  |  |  |
| B-cell inhibitor |  | 0.26(0.35) | [-0.42, 0.95] |  |  |  |  |  |
| IL-6 inhibitor |  | 0.29(0.30) | [-0.29, 0.88] |  |  |  |  |  |
| JAK1/3 inhibitor |  | 0.01(0.35) | [-0.67, 0.69] |  |  |  |  |  |
| T-cell inhibitor |  | -0.34(0.28) | [-0.90 0.21] |  |  |  |  |  |
| Age (mean) | 45 | 0.004(0.04) | [-0.06, 0.08] | **0.002** | 0.91 | 0.05 | 0.08 | 46.69 |
| MTX dose (mg/week) (mean) | 20 | -0.04(0.06) | [-0.17, 0.08] | **0.002** | 0.47 | 0.03 | 0.17 | 56.82 |
| HAQ-DI (mean) | 40 | -0.22(0.44) | [-1.08, 0.64] | **0.003** | 0.62 | 0.04 | 0.08 | 45.41 |
| Pain VAS (mean) | 21 | -0.04(0.02) | [-0.08, 0.01] | 0.74 | 0.10 | 0.003 | 0 | 2.204 |
| Exclusion of serious and recurrent infection | 45 | -0.06(0.18) | [-0.42, 0.2874] | **0.003** | 0.72 | 0.04 | 0.08 | 45.65 |
| ESR (mm/h) (mean) | 22 | 0.03(0.02) | [-0.0101, 0.07] | **<0.001** | 0.15 | 0.12 | 0.05 | 61.40 |
| Corticosteroid use (percent) | 30 | -.002(0.01) | [-0.02, 0.01] | **0.002** | 0.83 | 0.07 | 0.06 | 50.46 |
| RA duration (years) (mean) | 43 | -0.02(0.02) | [-0.06, 0.03] | **0.002** | 0.48 | 0.04 | 0.09 | 46.89 |
| Anti-CCP positive (percent) | 33 | 0.01(0.01) | [-0.01, 0.04] | 0.33 | 0.27 | 0.03 | 0.03 | 23.36 |
| Previous bDMARD use (percent) | 28 | -0.005(0.006) | [-0.02, 0.01] | **0.002** | 0.36 | 0.10 | 0.07 | 52.80 |
| Disease activity composite variable (mean) | 43 | -0.82(1.45) | [-3.66, 2.02] | **0.001** | 0.57 | 0.04 | 0.09 | 47.63 |
| Exclusion criteria composite variable | 45 | 0.16(0.07) | [0.01, 0.30] | **0.01** | 0.03 | 0.04 | 0.07 | 41.69 |

*Anti-CCP, anti-citrullinated protein antibodies; bDMARD, biological synthetic disease modifying anti rheumatic drug; CI, confidence interval; CRP, c-reactive protein; ESR, erythrocyte sedimentation rate; HAQ-DI, health assessment questionnaire-disability index; IL, interleukin; JAK, Janus kinase; MTX, methotrexate; n, number of study arms; RA, rheumatoid arthritis; RCT, randomised controlled trial; Reference, a category to which the coefficient estimates for the categorical predictors are compared to; SE, standard error; TNF-alpha, tumour necrosis factor alpha; VAS, visual analogue scale. Statistically significant moderators in bold.*

**Appendix 17: additional forest plots**

**Below the forest plots are shown that were created to separately assess inconsistencies for the total number of infected patients and the number of seriously infected patients by study design.**

**Prospective cohorts**

**(total number of infected patients)**

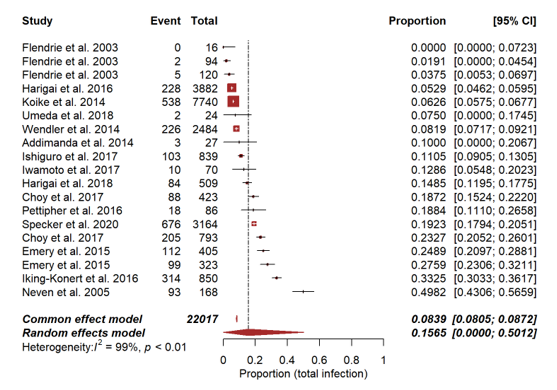

**(number of seriously infected patients)**

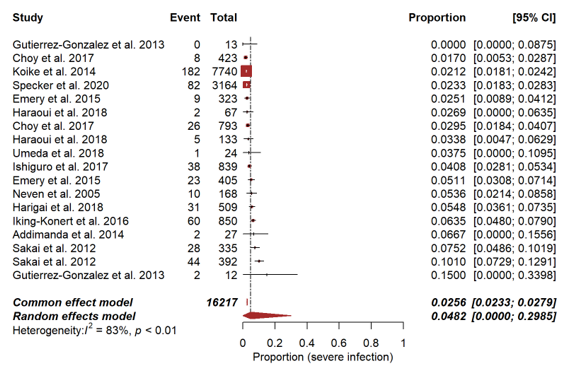

**Registry studies**

**(total number of infected patients)**

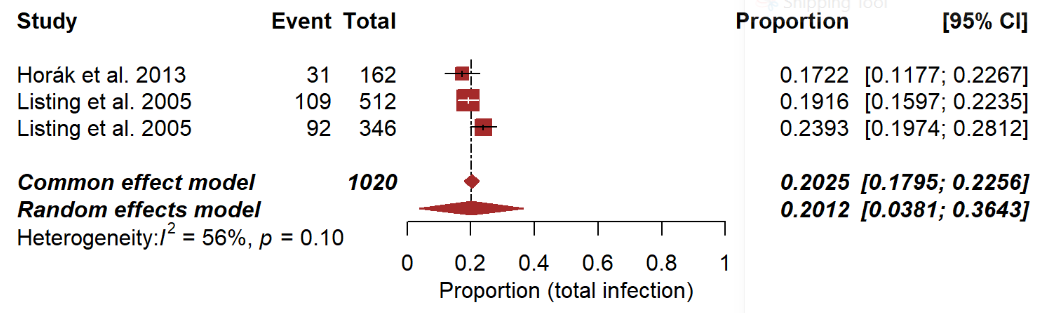

**(number of seriously infected patients)**

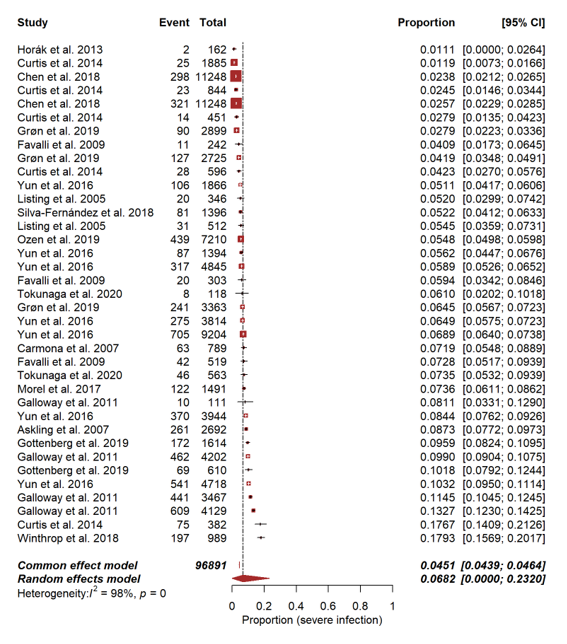

**RCT + OLE**

**(number of infected patients)**

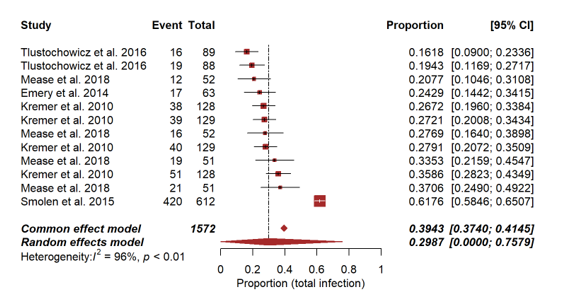

**(number of seriously infected patients)**

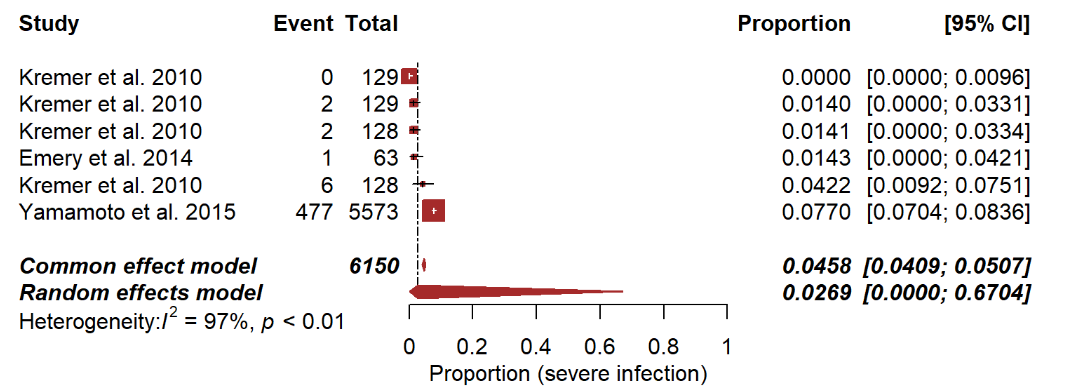

**Retrospective cohorts**

**(total number of infected patients)**

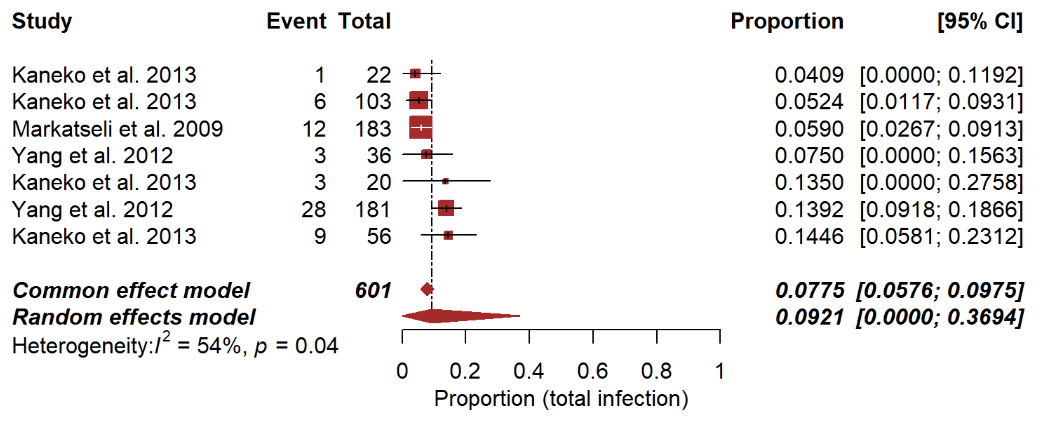

**(number of seriously infected patients)**

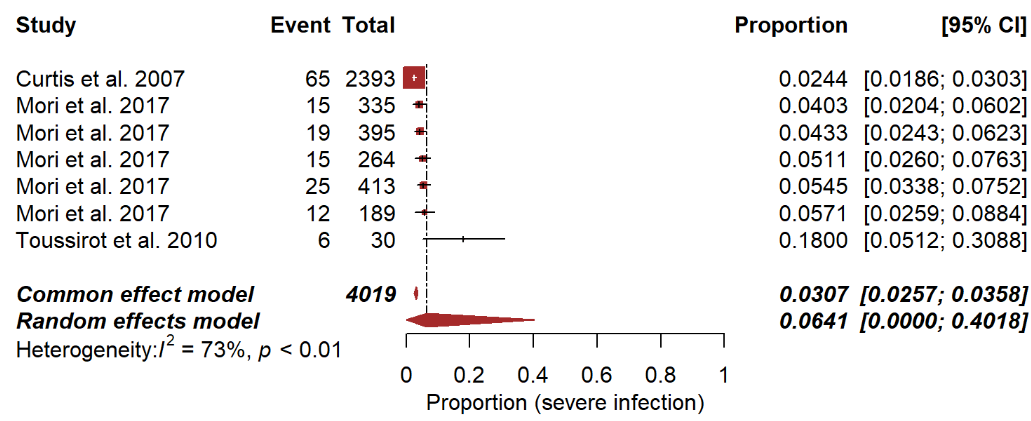

**Open Label Trials**

**(total number of infected patients)**

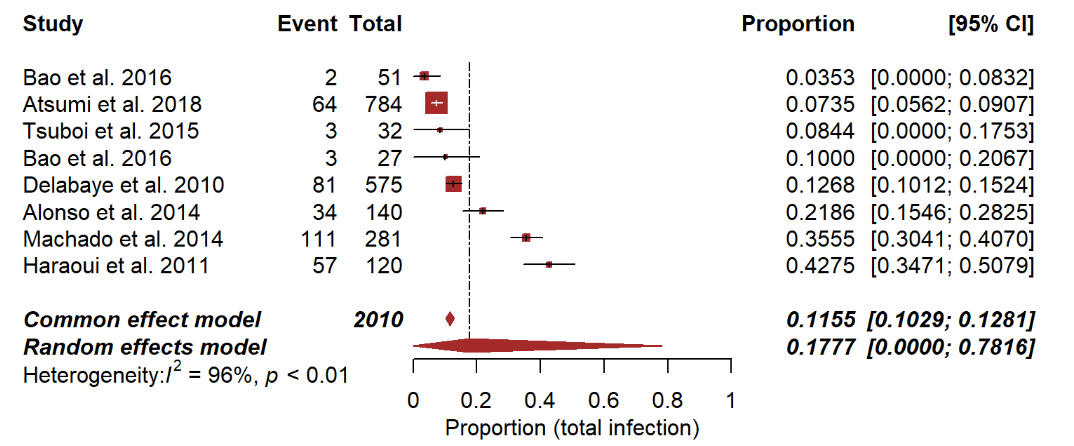

**(number of seriously infected patients)**

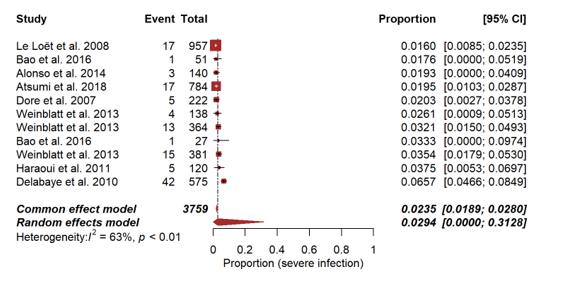

**Appendix 18: Characteristics of case reports**

In appendix 13, we present characteristics of patients from case reports at time of infection. This includes patient demographic characteristics, comorbidities, biological and csDMARD use, and also case characteristics such as outcome, details about treatment, hospitalization and what happened to the bDMARD after infection. Geographical location of case reports was also extracted.

**Table 1. Case report patient characteristics**

|  | **N** | **Percent** | **mean** | **SD** |
| --- | --- | --- | --- | --- |
| Female | 336 | 66.8 |  |  |
| Male | 142 | 28.2 |  |  |
| Unknown sex | 25 | 5.0 |  |  |
| Age |  |  | 61.02 | 11.7 |
| RA duration (years) |  |  | 12.3 | 9.2 |
| Any comorbidity | 155 | 30.8 |  |  |
| No comorbidity | 40 | 8.0 |  |  |
| Comorbidity unspecified | 298 | 59.2 |  |  |
| Corticosteroids use | 210 | 41.8 |  |  |
| No orticosteroid use | 188 | 37.4 |  |  |
| Corticosteroid use unspecified | 105 | 20.9 |  |  |
| csDMARD use | 285 | 56.7 |  |  |
| No csDMARD use | 107 | 21.3 |  |  |
| csDMARD use unspecified | 111 | 22.1 |  |  |
| **csDMARD** |  |  |  |  |
| Azathioprine | 15 | 4.7 |  |  |
| Bucillamine | 1 | 0.3 |  |  |
| Cyclophosphamide | 2 | 0.6 |  |  |
| Dapsone | 1 | 0.3 |  |  |
| Hydroxychloroquine | 13 | 4.0 |  |  |
| Iguratimod | 1 | 0.3 |  |  |
| Leflunomide | 23 | 7.1 |  |  |
| Methotrexate | 246 | 76.4 |  |  |
| Sulfasalazine | 14 | 4.3 |  |  |
| Tacrolimus | 6 | 1.9 |  |  |
| **Biological class** |  |  |  |  |
| B-cell inhibitor | 19 | 3.8 |  |  |
| Interleukin antagonist | 53 | 10.5 |  |  |
| JAK-inhibitor | 3 | 0.6 |  |  |
| T-cell inhibitor | 17 | 3.4 |  |  |
| TNF-alpha inhibitor | 411 | 81.7 |  |  |
| **Biological name** |  |  |  |  |
| Abatacept (T-cell) | 17 | 3.4 |  |  |
| Adalimumab | 102 | 20.3 |  |  |
| Anakinra (IL-1) | 5 | 1.0 |  |  |
| Certolizumab | 7 | 1.4 |  |  |
| Etanercept | 119 | 23.7 |  |  |
| Golimumab | 6 | 1.2 |  |  |
| Infliximab | 173 | 34.4 |  |  |
| Rituximab (B-cell) | 19 | 3.8 |  |  |
| Sarilumab (IL-6) | 1 | 0.2 |  |  |
| Tocilizumab (IL-6) | 48 | 9.5 |  |  |
| Tofacitinib (JAK-inhibitor) | 3 | 0.6 |  |  |
| <NA> | 3 | 0.6 |  |  |

*csDMARD, conventional synthetic disease modifying anti rheumatic drug; IL, interleukin; JAK, Janus kinase; n, number of patients; NA, not available; SD, standard deviation; RA, rheumatoid arthritis*

**Table 2. Case report patient comorbidities**

| **comorbidity** | **N** | **Percent** |
| --- | --- | --- |
| Alzheimer's disease | 1 | 0.4 |
| Amyloidosis | 3 | 1.1 |
| Anaemia | 1 | 0.4 |
| Ankylosing spondilitis | 2 | 0.7 |
| Aorta valve replacement | 1 | 0.4 |
| Aortic regurgitation | 1 | 0.4 |
| Arteriosclerosis obliterans | 1 | 0.4 |
| Asthma | 7 | 2.6 |
| Atrial fibrillation | 9 | 3.3 |
| Autoimmune hepatitis | 1 | 0.4 |
| Barrett's oesophagus | 1 | 0.4 |
| Bone sarcoma | 1 | 0.4 |
| Breast cancer | 2 | 0.7 |
| Brucellosis | 1 | 0.4 |
| Cerebellar ataxia | 1 | 0.4 |
| Cervical arthrodesis | 2 | 0.7 |
| Cervical cancer | 1 | 0.4 |
| Chronic kidney disease | 2 | 0.7 |
| Chronic stable thrombocytopenia | 1 | 0.4 |
| CML | 1 | 0.4 |
| Coccidioidomycosis (past infection) | 2 | 0.7 |
| Colectomy | 1 | 0.4 |
| COPD | 11 | 4.0 |
| Corneal transplant | 1 | 0.4 |
| Coronary artery disease | 2 | 0.7 |
| Crohn's disease | 2 | 0.7 |
| Degenerative spondylosis | 1 | 0.4 |
| Depression | 2 | 0.7 |
| Diabetes mellitus | 36 | 13.2 |
| Diverticulitis | 1 | 0.4 |
| Dyslipidaemia | 1 | 0.4 |
| Endovascular prosthesis | 1 | 0.4 |
| Fibromyalgia | 1 | 0.4 |
| Gallstones | 1 | 0.4 |
| Gastric cancer | 1 | 0.4 |
| Gastric ulcer | 4 | 1.5 |
| Gastroesophageal reflux disease | 4 | 1.5 |
| Hashimoto hypothyroidism | 2 | 0.7 |
| Hepatitis | 2 | 0.7 |
| Hepatitis B | 1 | 0.4 |
| Hiatal hernia | 1 | 0.4 |
| Hip prosthesis | 7 | 2.6 |
| HTLV-1 | 1 | 0.4 |
| Hyper IgG4-syndrome | 1 | 0.4 |
| Hyperlipidaemia | 9 | 3.3 |
| Hypertension | 40 | 14.7 |
| Hypertensive cardiomyopathy | 1 | 0.4 |
| Hypertrophic cardiomyopathy | 1 | 0.4 |
| Hypothyroidism | 7 | 2.6 |
| Interstitial lung disease | 1 | 0.4 |
| Intracerebral haemorrhage | 1 | 0.4 |
| Ischaemic heart disease | 9 | 3.3 |
| Leishmaniasis (past infection) | 1 | 0.4 |
| Lichen planus | 1 | 0.4 |
| LPD | 1 | 0.4 |
| Lumbar fusion | 2 | 0.7 |
| Lumbar spinal canal stenosis | 1 | 0.4 |
| Lung fibrosis | 1 | 0.4 |
| Myasthenia gravis | 2 | 0.7 |
| Mycobacterium xenopi septic arthritis | 1 | 0.4 |
| OSAS | 2 | 0.7 |
| Osteopenia | 2 | 0.7 |
| Osteoporosis | 4 | 1.5 |
| PJP | 1 | 0.4 |
| Prostatic adenoma resection | 1 | 0.4 |
| Psoriasis | 2 | 0.7 |
| Pulmonary embolism recurrent | 1 | 0.4 |
| Pulmonary micronodules | 1 | 0.4 |
| Pulmonary NTM infection | 2 | 0.7 |
| RA lung involvement | 1 | 0.4 |
| Recent dental examination | 1 | 0.4 |
| Recent gastroenteritis | 1 | 0.4 |
| Recurrent UTIs | 1 | 0.4 |
| Recurrent uveitis | 1 | 0.4 |
| Shoulder prosthesis | 1 | 0.4 |
| Sjögren syndrome | 4 | 1.5 |
| Skin ulcer | 1 | 0.4 |
| Spinal stenosis | 1 | 0.4 |
| Splenectomy | 2 | 0.7 |
| Spondylodesis | 1 | 0.4 |
| Squamous cell carcinoma of tongue | 1 | 0.4 |
| Systemic sclerosis | 2 | 0.7 |
| Thyroid carcinoma | 1 | 0.4 |
| Total hip replacement | 2 | 0.7 |
| Total knee replacement | 18 | 6.6 |
| Tuberculosis | 1 | 0.4 |
| Tuberculosis, pulmonary | 9 | 3.3 |
| Ulcerative colitis | 1 | 0.4 |
| Vasculitis | 1 | 0.4 |

*CML, chronic myeloid leukaemia; COPD, chronic obstructive pulmonary disease; csDMARD, conventional synthetic disease modifying anti rheumatic drug; HTLV-1, human T lymphotropic virus type 1; JAK, Janus kinase; LPD, lymphoproliferative disease; n, number of patients; NTM, non-tuberculous mycobacteria; OSAS, obstructive sleep apnaea syndrome; PJP, Pneumocystis jirovecii pneumonia; RA, rheumatoid arthritis; TNF, tumour necrosis factor; UTI, urinary tract infection. Patients could have more than one comorbidity.*

**Table 3. Case characteristics**

| **Hospitalization** | **N** | **Percent** | **mean** | **SD** |
| --- | --- | --- | --- | --- |
| Yes | 185 | 36.78 |  |  |
| No | 9 | 61.43 |  |  |
| Not specified | 309 | 1.79 |  |  |
| Hospitalization duration (n=59) |  |  | 27.05 | 31.68 |
| **Treatment** |  |  |  |  |
| Targeted therapy | 183 | 36.38 |  |  |
| Discontinuation immunosuppression | 16 | 3.18 |  |  |
| Both | 243 | 48.31 |  |  |
| Neither | 4 | 3.18 |  |  |
| **Case outcome** |  |  |  |  |
| Chronic illness | 3 | 0.6 |  |  |
| Death | 53 | 10.54 |  |  |
| Progression | 6 | 1.19 |  |  |
| Resolution | 391 | 77.73 |  |  |
| Stabilization | 1 | 0.2 |  |  |
| Not specified | 49 | 9.74 |  |  |
| **bDMARD outcome** |  |  |  |  |
| Continued | 16 | 3.18 |  |  |
| Continued lower dose | 1 | 0.2 |  |  |
| Discontinued | 148 | 29.42 |  |  |
| Reinitiation | 43 | 8355 |  |  |
| Reinitiation different biological | 17 | 3.38 |  |  |
| Not specified | 278 | 55.27 |  |  |
| **Relapse** |  |  |  |  |
| Yes | 14 | 2.78 |  |  |
| No | 94 | 18.69 |  |  |
| Not specified | 395 | 78.53 |  |  |
| Follow up duration (n = 82) |  |  | 11.74 | 9.31 |

*bDMARD, biological disease modifying anti rheumatic drug; n, number of patients; SD, standard deviation*

**Table 4. Case report countries**

| **Country** | **N** |
| --- | --- |
| USA | 83 |
| Japan | 77 |
| France | 39 |
| Spain | 28 |
| UK | 20 |
| Italy | 17 |
| Netherlands | 14 |
| Germany | 12 |
| Greece | 11 |
| Ireland | 7 |
| Brazil | 6 |
| Australia | 5 |
| Canada | 5 |
| Turkey | 5 |
| Belgium | 4 |
| Colombia | 4 |
| Switzerland | 4 |
| Austria | 3 |
| Denmark | 3 |
| Romania | 3 |
| South Korea | 3 |
| Croatia | 2 |
| India | 2 |
| Israel | 2 |
| Taiwan | 2 |
| Not available | 2 |
| Argentina | 1 |
| Burkina Faso | 1 |
| Lebanon | 1 |
| Malaysia | 1 |
| Mexico | 1 |
| Poland | 1 |
| Portugal | 1 |
| Puerto Rico | 1 |
| Sweden | 1 |

*N = number of case reports*

**Appendix 19: Identified micro-organisms in the case reports**

We extracted information on the causative micro-organisms in each case. One patient could be infected by multiple micro-organisms.

**Table 1. All identified micro-organisms in all case reports per biological (class)**

| **Micro-organism** | **TNF-alpha inhibitors** | **Tocilizumab**  **(IL-6)** | **Anakinra**  **(IL-1)** | **Abatacept** | **Rituximab** | **JAK-inhibitors** |
| --- | --- | --- | --- | --- | --- | --- |
| Acinetobacter baumannii | 1 |  |  |  |  |  |
| Actinobacillus ureae | 1 |  |  |  |  |  |
| Actinomyces spp | 1 |  |  |  |  |  |
| Adenovirus | 1 |  |  |  |  |  |
| Aspergillus spp | 10 | 1 |  | 1 |  |  |
| Babesia microtii | 1 |  |  |  | 1 |  |
| Bacteroides spp | 1 | 1 |  |  |  |  |
| Bartonella henselae | 1 | 2 |  | 1 |  |  |
| Borrelia Afzelii | 1 |  |  |  |  |  |
| Brachiola algerae | 1 |  |  |  |  |  |
| Brucella spp | 2 |  |  |  |  |  |
| Campylobacter fetus |  |  |  |  | 2 |  |
| Candida spp | 5 |  |  | 1 |  |  |
| Candidatus Neoehrlichia mikurensis |  |  |  |  | 1 |  |
| Capnocytophaga cynodegmi | 1 |  |  |  |  |  |
| Cladophialophora bantiana |  |  |  |  |  | 1 |
| Coagulase-negative staphylococci |  | 1 |  |  |  |  |
| Coccidioides spp | 11 |  |  |  |  |  |
| Coxiella burneti | 1 |  |  |  | 1 |  |
| Cryptococcus supp | 12 |  |  | 1 | 1 |  |
| Cutibacterium acnes | 1 |  |  |  |  |  |
| Cytomegalovirus | 2 | 1 |  | 1 | 1 |  |
| Echinococcus multilocularis | 1 |  |  |  | 1 |  |
| Enterobacter aerogenes |  | 1 |  |  |  |  |
| Enterobacter cloacae | 1 |  |  |  |  |  |
| Epstein-Barr virus | 3 | 1 |  | 3 |  |  |
| Escherichia coli | 2 | 2 | 1 |  |  |  |
| Finegoldia magna | 1 |  |  |  |  |  |
| Francisella tularensis | 1 |  |  |  |  |  |
| Gram-positive cocci |  |  |  | 1 |  |  |
| Gram-positive streptococci | 1 | 1 | 1 |  |  |  |
| Haemophilus influenzae |  | 2 |  |  |  |  |
| HHV-6 | 1 |  |  |  |  |  |
| Hepatitis E virus |  | 2 |  | 1 | 1 |  |
| Histoplasma capsulatum | 20 | 1 |  | 1 |  |  |
| HSV1 | 1 |  |  |  |  |  |
| HSV2 | 1 |  |  |  |  |  |
| Influenza virus |  | 1 |  |  |  |  |
| JC virus | 1 |  |  |  |  |  |
| Klebsiella pneumoniae |  | 1 |  |  |  |  |
| Kocuria kristinae | 1 |  |  |  |  |  |
| Legionella pneumophila | 17 | 1 |  |  |  |  |
| Leishmania spp | 19 |  | 1 |  | 1 |  |
| Listeria monocytogenes | 23 | 1 |  |  |  |  |
| Measles morbillivirus | 1 |  |  |  |  |  |
| Molluscum contagiosum virus | 1 |  |  |  |  |  |
| Moraxella catarrhalis | 1 |  |  |  |  |  |
| Mucor spp | 3 |  |  |  |  |  |
| Mycobacterium abscessus | 4 | 1 |  |  |  |  |
| Mycobacterium avium | 10 | 2 |  |  |  |  |
| Mycobacterium avium-intracellulare | 1 | 1 |  | 1 |  |  |
| Mycobacterium bovis | 3 |  |  |  |  |  |
| Mycobacterium chelonae | 7 |  |  |  |  |  |
| Mycobacterium fortuitum | 2 |  |  |  |  |  |
| Mycobacterium haemophilum | 3 |  |  |  |  |  |
| Mycobacterium heckeshornense | 2 |  |  |  |  |  |
| Mycobacterium intracellulare | 4 |  |  |  |  |  |
| Mycobacterium kansasii | 1 |  |  |  |  |  |
| Mycobacterium leprae | 4 | 1 |  |  |  |  |
| Mycobacterium mageritense | 1 |  |  |  |  |  |
| Mycobacterium marinum | 5 |  |  |  |  |  |
| Mycobacterium mucogenicum | 2 |  |  |  |  |  |
| Mycobacterium szulgai | 1 |  |  |  |  |  |
| Mycobacterium tuberculosis | 81 |  | 2 |  | 2 |  |
| Mycobacterium xenopi | 2 |  |  |  |  |  |
| Neisseria meningitidis | 1 |  |  |  |  |  |
| Nocardia spp | 3 | 1 |  | 1 | 1 |  |
| Paracoccidioides spp | 1 |  |  |  |  |  |
| Parvimonas micra | 1 |  |  |  |  |  |
| Parvovirus B19 |  | 1 |  |  | 1 |  |
| Pasteurella multocida | 1 | 1 |  |  | 1 |  |
| Pentatrichomonas hominis | 1 |  |  |  |  |  |
| Peptostreptococcus spp | 1 |  |  |  |  |  |
| Phaeoacremonium spp | 1 |  |  |  |  |  |
| Plasmodium falciparum | 1 |  |  |  |  |  |
| Pneumocystis jirovecii | 24 |  |  | 1 | 1 | 2 |
| Polymicrobial | 3 |  |  |  |  |  |
| Pseudomonas spp | 2 | 1 |  |  |  |  |
| Rhodococcus erythropolis |  |  |  |  | 1 |  |
| Rickettsia rickettsi | 1 |  |  |  |  |  |
| Roseomonas mucosa | 1 |  |  |  |  |  |
| Rothia dentocariosa | 1 |  |  |  |  |  |
| Salmonella spp | 28 |  |  |  |  |  |
| Sarcoptes scabiei | 1 | 1 |  |  |  |  |
| Scedosporium apiospermum | 1 |  |  |  |  |  |
| Serratia marcescens |  | 1 |  |  |  |  |
| Staphylococcus aureus | 15 | 8 | 2 |  | 1 |  |
| Stenotrophomonas maltophilia | 1 | 2 |  |  |  |  |
| Streptobacillus monilliformis |  | 1 |  |  |  |  |
| Streptococcus agalactiae | 1 |  |  |  |  |  |
| Streptococcus constellatus | 2 |  |  |  |  |  |
| Streptococcus pneumoniae | 6 | 2 |  |  |  |  |
| Streptococcus pyogenes | 5 | 3 |  | 1 |  |  |
| Streptococcus viridans | 2 |  |  |  |  |  |
| Strongyloides stercoralis | 3 |  |  |  |  |  |
| Toxoplasma gondii | 7 |  |  |  |  |  |
| Treponema pallidum | 1 |  |  |  |  |  |
| Tropheryme whipplei | 1 |  |  |  |  |  |
| Varicella zoster virus | 3 | 1 |  | 1 |  |  |
| West Nile virus | 12 |  |  |  | 1 |  |

*HHV-6, human herpes virus 6; HSV 1/2, herpes simplex virus type 1/2; IL, interleukin; JAK, Janus kinase; JC virus, John Cunningham virus; TNF, tumour necrosis factor*

**Appendix 20: figures related to the case reports**

Per case, information on the anatomical location of the infection was obtained. Reported causative micro-organisms were subdivided into viral, parasitic, mycobacterial, intracellular bacterial, and other bacterial pathogens.

**Figure 1. Number of cases with an affected organ, for different organ systems, in case reports**

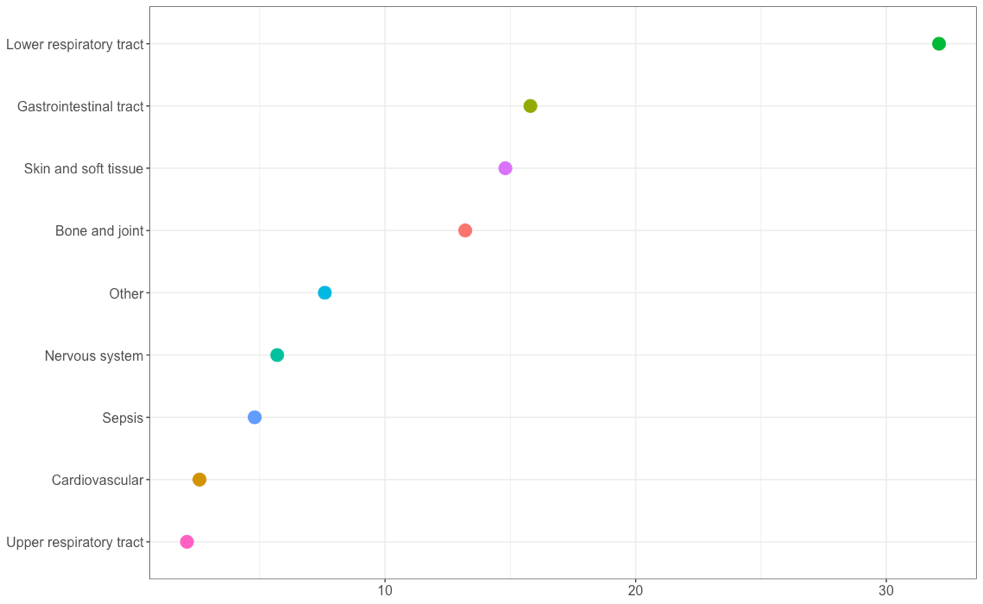

**Figure 2. Percentage of different types of infections per biological**

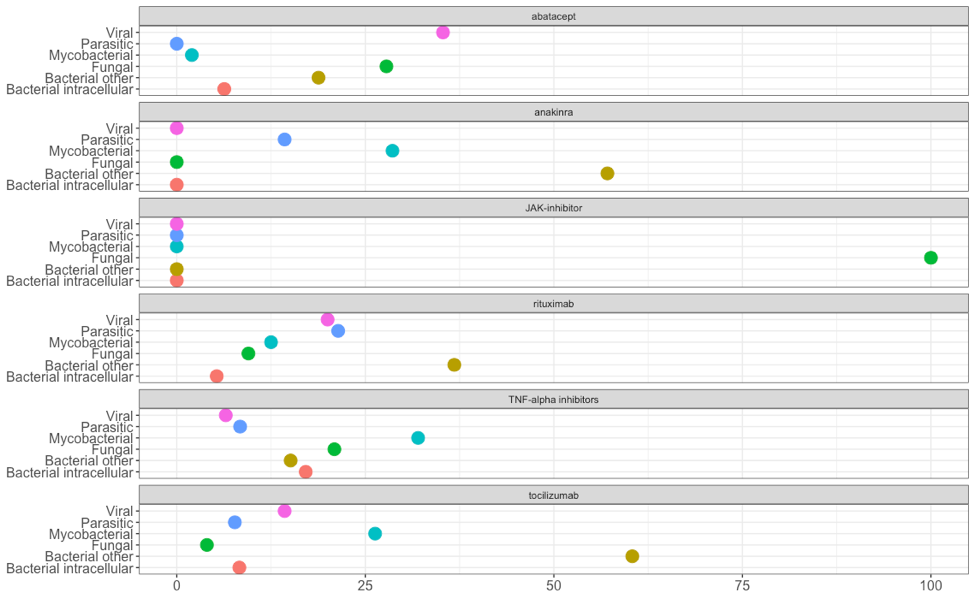
